# Structural brain alterations associated with suicidal thoughts and behaviors in young people: results across 21 international studies from the ENIGMA Suicidal Thoughts and Behaviours consortium

**DOI:** 10.1101/2021.09.27.21264068

**Authors:** Laura S. van Velzen, Maria R. Dauvermann, Lejla Colic, Luca M. Villa, Hannah S. Savage, Yara J. Toenders, Alyssa H. Zhu, Joanna K. Bright, Adrián I. Campos, Lauren Salminen, Sonia Ambrogi, Rosa Ayesa-Arriola, Nerisa Banaj, Zeynep Başgöze, Jochen Bauer, Karina Blair, Robert James Blair, Katharina Brosch, Yuqi Cheng, Romain Colle, Colm G. Connolly, Emmanuelle Corruble, Baptiste Couvy-Duchesne, Benedicto Crespo-Facorro, Kathryn R. Cullen, Udo Dannlowski, Christopher G. Davey, Katharina Dohm, Janice M. Fullerton, Ali Saffet Gonul, Ian H. Gotlib, Dominik Grotegerd, Tim Hahn, Ben J. Harrison, Mengxin He, Ian B. Hickie, Tiffany C. Ho, Frank Iorfino, Andreas Jansen, Fabrice Jollant, Tilo Kircher, Bonnie Klimes-Dougan, Melissa Klug, Elisabeth J. Leehr, Elizabeth T.C. Lippard, Katie A. McLaughlin, Susanne Meinert, Adam Bryant Miller, Philip B. Mitchell, Benson Mwangi, Igor Nenadić, Amar Ojha, Bronwyn J. Overs, Julia-Katharina Pfarr, Fabrizio Piras, Kai G. Ringwald, Gloria Roberts, Georg Romer, Marsal Sanches, Margaret A. Sheridan, Jair C Soares, Gianfranco Spalletta, Frederike Stein, Giana I. Teresi, Diana Tordesillas-Gutiérrez, Aslihan Uyar-Demir, Nic J.A. van der Wee, Steven J. van der Werff, Robert R.J.M. Vermeiren, Alexandra Winter, Mon-Ju Wu, Tony T. Yang, Paul M. Thompson, Miguel E. Rentería, Neda Jahanshad, Hilary P. Blumberg, Anne-Laura van Harmelen, Lianne Schmaal

**Affiliations:** Orygen, Parkville, VIC, Australia; Centre for Youth Mental Health, University of Melbourne, Melbourne, VIC, Australia; Department of Psychiatry, University of Cambridge, Cambridge, UK; Department of Forensic and Neurodevelopmental Science; Institute of Psychiatry, Psychology, and Neuroscience; King’s College London, London, UK; McGovern Institute for Brain Research, Massachusetts Institute of Technology, Cambridge, MA, USA; Department of Psychiatry, Yale School of Medicine, New Haven, CT, USA; Department of Psychiatry and Psychotherapy, University Hospital Jena, Jena, Germany; Center for Intervention and Research on adaptive and maladaptive brain circuits underlying mental health (CIRC), Jena, Germany; Department of Psychiatry, University of Oxford, Oxford, UK; Melbourne Neuropsychiatry Centre, Department of Psychiatry, The University of Melbourne, Victoria, Australia; Imaging Genetics Center, Mark and Mary Stevens Neuroimaging and Informatics Institute, Keck School of Medicine, University of Southern California, Marina del Rey, CA, USA; Social Genetic & Developmental Psychiatry Centre, Institute of Psychiatry, Psychology & Neuroscience, King’s College London, London, UK; Department of Genetics & Computational Biology, QIMR Berghofer Medical Research Institute, Brisbane, QLD, Australia; School of Biomedical Sciences, Faculty of Medicine, University of Queensland, Brisbane, QLD, Australia; Laboratory of Neuropsychiatry, IRCCS Santa Lucia Foundation, Rome, Italy; Department of Psychiatry, Marqués de Valdecilla University Hospital, IDIVAL, School of Medicine, University of Cantabria, Santander, Spain; Centro Investigación Biomédica en Red de Salud Mental (CIBERSAM), Sevilla, Spain; Department of Psychiatry and Behavioural Sciences, University of Minnesota Medical School, USA; University Clinic for Radiology, University of Münster, Münster, Germany; Center for Neurobehavioral Research, Boys Town National Research Hospital, Boys Town, NE, USA; Department of Psychiatry and Psychotherapy, Marburg University, Marburg, Germany; CMBB, Marburg, Germany; Department of Psychiatry, First Affiliated Hospital of Kunming Medical College, China; Yunnan Province Clinical Research Center for Psychiatry, China; MOODS Team, CESP, INSERM, Faculté de Médecine, Univ Paris-Saclay, Le Kremlin Bicêtre, 94275, France; Service Hospitalo-Universitaire de Psychiatrie de Bicêtre, Hôpitaux Universitaires Paris-Saclay, Assistance Publique-Hôpitaux de Paris, Hôpital de Bicêtre, Le Kremlin Bicêtre, F-94275, France; Department of Biomedical Sciences, Florida State University, Tallahassee, FL, USA; Paris Brain Institute (ICM), Inserm (U1127), CNRS (UMR 7225), Sorbonne University, Inria Paris (Aramis project-team), Paris, France; Institute for Molecular Bioscience, The University of Queensland, St Lucia, QLD, Australia; Virgen del Rocío University Hospital, IBiS, University of Sevilla, Sevilla, Spain; Institute for Translational Psychiatry, University of Münster, Münster, Germany; Department of Psychiatry, The University of Melbourne, Victoria, Australia; Neuroscience Research Australia, Randwick, NSW, Australia; School of Medical Sciences, University of New South Wales, Kensington, NSW, Australia; SoCAT Lab, Department of Psychiatry, School of Medicine, Ege University, Izmir, Turkey; Department of Psychology, Stanford University, Stanford, CA, USA; Brain and Mind Centre, University of Sydney, Camperdown, Australia; Department of Psychiatry and Behavioral Sciences, University of California San Francisco, San Francisco, CA, USA; Weill Institute for Neurosciences, University of California San Francisco, San Francisco, CA, USA; Core-Facility Brainimaging, Faculty of Medicine, University of Marburg; Université de Paris & GHU Paris Psychiatrie et Neurosciences, Paris, France; McGill University, Department of psychiatry, Montréal, Canada; Academic Hospital (CHU) Nîmes, France; University of Minnesota, Department of Psychology, Minneapolis, MN, USA; Department of Psychiatry and Behavioral Sciences, Dell Medical School, University of Texas at Austin, USA; Institute of Early Life Adversity Research, Dell Medical School, University of Texas at Austin, USA; Waggoner Center for Alcohol and Addiction Research, University of Texas at Austin, USA; Mulva Clinic for Neuroscience, Dell Medical School, University of Texas at Austin, USA; Department of Psychology, Harvard University, USA; Institute for Translational Neuroscience, University of Münster, Münster, Germany; Mental Health Risk and Resilience Research Program, RTI International, Research Triangle Park, NC; Department of Psychology and Neuroscience, University of North Carolina at Chapel Hill, USA; School of Psychiatry, University of New South Wales, Kensington, NSW, Australia; Center Of Excellence On Mood Disorders, The University of Texas - Health Science Center at Houston; Louis A. Faillace, MD, Department of Psychiatry and Behavioral Sciences at McGovern Medical School, The University of Texas - Health Science Center at Houston; Center for Neuroscience, University of Pittsburgh, Pittsburgh, PA; Center for the Neural Basis of Cognition, University of Pittsburgh, Pittsburgh, PA; Department of Child & Adolescent Psychiatry, Psychosomatics and Psychotherapy, University Hospital Münster; Department of Psychiatry and Behavioral Sciences, Baylor College of Medicine, Houston, Texas, USA; Department of Psychology, University of Pittsburgh, Pittsburgh, PA, USA; Department of Radiology. Marqués de Valdecilla University Hospital, Valdecilla Biomedical Research Institute IDIVAL, Spain; Department of Psychiatry, Leiden University Medical Center, Leiden, the Netherlands; Leiden Institute for Brain and Cognition. Leiden University. Leiden, the Netherlands; Leids Universitair Behandel- en Expertise Centrum, Leiden, the Netherlands; Child and Adolescent Psychiatry Leiden University Medical Center, Leiden, the Netherlands; Youz: child and adolescent psychiatry, Netherlands; Department of Psychiatry and Behavioral Sciences, Division of Child and Adolescent Psychiatry, Weill Institute for Neurosciences, UCSF, USA; Global Brain Health Institute, University of California, San Francisco, San Francisco, CA, USA; Department of Radiology and Biomedical Imaging, Yale School of Medicine, USA; Child Study Center, Yale School of Medicine, USA; Social Security and Resilience Programme, Education and Child Studies, Leiden University, Leiden, the Netherlands

## Abstract

**Objective:** Identifying brain differences associated with suicidal thoughts and behaviors (STBs) in young people is critical to understanding their development and generating effective approaches to early intervention and prevention. The ENIGMA Suicidal Thoughts and Behaviours (ENIGMA-STB) consortium analyzed neuroimaging data harmonized across sites to examine brain morphology associated with STBs in youth.

**Methods:** First, we examined associations among regional brain structure and STBs, which were assessed in six samples of youth with mood disorders, using the Columbia Suicide Severity Rating Scale (C-SSRS; *N*=577). Second, we combined this sample with a larger sample (total 21 sites) in which STBs were assessed using various instruments. MRI metrics were compared among healthy controls without STBs (HC; *N*=688), clinical controls without STBs (CC; *N*=648), and young people with psychiatric diagnoses and current suicidal ideation (*N*=406). In separate analyses, MRI metrics were compared among HCs (*N*=335), CCs (*N*=768), and suicide attempters (*N*=254).

**Results:** In the homogeneous C-SSRS sample, surface area of the frontal pole was lower in young people with mood disorders and history of actual suicide attempts (*N*=163) than those without (*N*=394; FDR-*p*<.001; Cohen’s *d*=.334). When expanding to more clinically heterogeneous samples, we also found lower surface area of the frontal pole in those with a history of suicide attempts (Cohen’s *d*=.22).

**Conclusions:** Lower frontal pole surface area may represent a vulnerability for a suicide attempt; however, more research is needed to understand the nature of its relationship to suicide risk.

## Introduction

Suicide is a leading cause of death worldwide, with around 800,000 deaths by suicide occurring annually (1). Suicidal thoughts and behaviors (STBs) typically emerge during adolescence (2). Suicide is the second leading cause of death for young people aged between 15 and 29 (1). It has been estimated that between 11 and 29% of adolescents report suicidal ideation (suicidal thoughts), and 2-10% of adolescents attempted suicide in the past year (3). Unfortunately, the number of suicide attempts among children and adolescents has continued to increase sharply despite national and international prevention efforts (4).

To improve targeting of prevention and intervention efforts and thereby reduce the number of deaths by suicide in this age group, we must increase our understanding of the mechanisms underlying both suicidal thoughts and suicidal behaviors (including suicide attempts) in young people. Neuroimaging, including magnetic resonance imaging (MRI), is a useful tool with which to identify biological risk markers for STBs *in vivo*. Many neuroimaging studies have been published examining the neural substrates of STBs in the past 20 years, but few have focused on STBs in youth (for a review, see (5)). Although several of these studies support lower regional brain volumes, particularly in ventral and dorsal prefrontal and also in temporal regions (6–9) in suicide attempters with mood disorders, negative findings have also been reported (10, 11). Structural brain alterations related to suicidal ideation in young people have inconsistently been reported in the striatum and temporal lobes (12–14).

In addition to the small number of studies focusing on youth, neuroimaging studies investigating associations between structural brain measures and STBs have also been limited by small sample sizes (5). There are multiple limitations associated with small sample sizes. First, small sample sizes decrease power (i.e., the probability of identifying true effects), increase the probability of false-negative effects, and inflate the effect size estimate when an actual effect is observed (15). Second, there may be small yet clinically significant associations between STBs and brain structure. To reliably identify these effects, larger samples are needed. Another significant limitation of previous work is that clinical controls are often not included, making it difficult to understand if alterations are specific to STBs or reflect mental health disorders in general (5).

To address these limitations, the suicide project within the ENIGMA Major Depressive Disorder (ENIGMA-MDD) consortium pooled data from 18 different studies worldwide to examine associations between brain morphology and suicide attempt in MDD patients (16, 17). While prior structural imaging studies of STBs primarily assessed regional volumes, the ENIGMA studies also examined cortical surface area and thickness, which is essential as unique genetic and environmental factors are implicated in the development of these cortical features. Findings showed a lower volume of the thalamus and pallidum and a smaller surface area of the inferior parietal lobe in adults with MDD and a history of suicide attempts (*N*=679) compared to MDD individuals without a history of suicide attempt (*N*=5,484). However, these studies did not examine MRI correlates of suicidal ideation. In addition, studies within ENIGMA-MDD are limited to individuals with MDD, while STBs are transdiagnostic phenomena, and the extent of neurobiological mechanisms underlying STBs that are common to or may differ across psychiatric disorders is unknown. Finally, these previous studies did not examine structural brain alterations in children and adolescents.

Therefore, we established the transdiagnostic ENIGMA Suicidal Thoughts and Behaviours (ENIGMA-STB) consortium, which allows investigation of neural correlates of STBs across a range of psychiatric conditions, leveraging many samples worldwide. This large dataset enables assessment of structural brain alterations that are common across groups (e.g., groups with a variety of psychiatric conditions including mood disorders, anxiety disorders, post-traumatic stress disorder [PTSD], addiction, and obsessive-compulsive disorder [OCD]), but also alterations that are specific to subgroups, such as males or females. For this ENIGMA-STB study, we focused specifically on young persons, as there is limited information concerning the mechanisms underlying STBs in youth, even though suicide is a leading cause of death in this population. We first pooled data from six studies that were more homogeneous, as all used the Columbia Suicide Severity Rating Scale (C-SSRS) (providing data on the intensity of suicidal ideation and suicide attempts defined specifically as “actual attempts,” i.e., with at least some intent to die) to assess STBs. This sample aimed to investigate differences in structural MRI measures between young persons with a lifetime history of an actual suicide attempt compared to those without and associations with the intensity of current suicidal ideation. To examine how well the observed associations generalize to different STB measures and across psychiatric conditions, we conducted an additional large-scale mega-analysis combining the C-SSRS data with data from additional studies that included young people with different psychiatric conditions and that employed different STB instruments (total of 21 studies). In these analyses in the larger sample, we aimed to identify structural brain alterations in young persons with (1) a lifetime history of a suicide attempt; and (2) current (in the past week, two weeks, or month) suicidal ideation (but no history of attempt), compared to healthy controls (HC) and clinical controls (CC; individuals who have a psychiatric disorder but no current ideation or lifetime attempt). Based on previous findings in adolescents, we predicted that a lifetime history of suicide attempts would be associated with structural alterations in the prefrontal cortex (6, 8, 9), temporal cortex, and caudate (12, 13).

## Methods

### Samples

This mega-analysis pooled data from 21 international studies from ten countries to examine the association between STBs and brain structure in young people ages 8-25 years. Demographic characteristics of the research participants at different sites are presented in Tables 1 and 2 and Figures 1 and 2, while the inclusion/exclusion criteria for the different studies are presented in Table S1. All sites obtained ethics approval from their local institutional review boards and ethics committees. All participants who were 18 years old and over provided written informed consent, and those aged under age 18 years provided written informed assent in addition to written informed consent from a parent/guardian at the local recruitment institution.

**Table 1.** Descriptive statistics for studies included in the ideation analysis. Presented here are age (median, minimum, maximum) and sex for the three groups (HC: healthy controls, CC: clinical controls, Ideation: group with current suicidal ideation) for the different sites included in the analysis on suicidal ideation.

| Site | Age<br>HC<br>(years) | Age<br>CC<br>(years) | Age<br>ideatio<br>n | %<br>female<br>HC | %<br>female<br>CC | %<br>female<br>ideatio<br>n | Total<br>N HC | Total<br>N CC | Total<br>N<br>Ideatio<br>n |
| --- | --- | --- | --- | --- | --- | --- | --- | --- | --- |
| Boystown<br>(USA) | 17.0<br>(14-18) | 16.0<br>(12-19) | 16.0<br>(12-18) | 22.2 | 30.7 | 63.0 | 9 | 114 | 27 |
| DEP-ARREST-<br>CLIN - MOODS<br>(France) | 21.0<br>(20-25) |  | 22.0<br>(18-25) | 54.5 |  | 70.0 | 11 | 0 | 10 |
| EPISCA<br>(Netherlands) | 14.0<br>(13-19) | 16.0<br>(13-20) | 16.0<br>(12-20) | 86.2 | 83.3 | 87.5 | 29 | 12 | 24 |
| FOR2107-Marburg<br>(Germany) | 23.0<br>(18-25) | 24.0<br>(18-25) | 23.0<br>(18-25) | 67.2 | 84.4 | 49.1 | 125 | 32 | 57 |
| FOR2107-Münster<br>(Germany) | 23.0<br>(18-25) | 22.0<br>(18-25) | 23.0<br>(19-25) | 71.7 | 77.3 | 50.0 | 106 | 22 | 26 |
| Houston BD<br>(USA) | 14.0<br>(8-25) | 14.0<br>(8-25) | 14.0<br>(10-24) | 54.3 | 31.6 | 66.7 | 81 | 57 | 9 |
| MDD Cohort<br>(China) | 23.0<br>(22-24) | 22.0<br>(18-25) | 21.0<br>(16-25) | 80.0 | 44.4 | 66.7 | 5 | 9 | 12 |
| Melbourne -<br>YODA<br>(Australia) |  | 18.0<br>(15-24) | 19.0<br>(15-25) |  | 56.1 | 53.3 | 0 | 41 | 45 |
| MR-IMPACT<br>(UK) |  | 15.0<br>(12-17) | 15.0<br>(11-17) |  | 73.2 | 64.5 | 0 | 41 | 31 |
| Muenster<br>Neuroimaging<br>Cohort<br>(Germany) | 22.0<br>(17-25) | 23.0<br>(16-25) | 22.0<br>(17-25) | 47.5 | 57.9 | 50.0 | 80 | 19 | 40 |
| SOCAT<br>(Turkey) | 23.0<br>(17-25) |  | 23.0<br>(19-25) | 100.0 |  | 95.0 | 37 | 0 | 20 |
| Stanford TAD<br>(USA) |  | 17.0<br>(15-18) | 16.0<br>(14-18) |  | 70.0 | 85.0 | 0 | 20 | 20 |
| Sydney Brain<br>and Mind<br>Centre<br>(Australia) | 23.0<br>(18-25) | 19.0<br>(12-25) | 20.0<br>(14-25) | 60.0 | 61.4 | 85.7 | 25 | 83 | 21 |
| UCSF<br>(USA) | 15.0<br>(13-17) | 16.0<br>(13-18) | 14.5<br>(13-17) | 48.8 | 66.7 | 41.7 | 82 | 45 | 12 |
| University of<br>Minnesota<br>(USA) | 16.0<br>(12-19) | 16.0<br>(13-20) | 16.0<br>(12-19) | 54.5 | 74.3 | 84.0 | 22 | 35 | 25 |
| University of<br>Texas- Austin -<br>Bipolar Seed<br>Program<br>(USA) | 21.0<br>(18-25) | 21.0<br>(18-25) | 20.5<br>(19-25) | 69.2 | 75.0 | 83.3 | 26 | 12 | 6 |
| UWashington/<br>Harvard<br>(USA) | 11.0<br>(8-16) | 10.5<br>(8-16) | 14.5<br>(8-16) | 50.0 | 50.0 | 58.3 | 50 | 24 | 12 |
| Yale School of<br>Medicine<br>(USA) |  | 20.0<br>(13-25) | 17.0<br>(15-23) |  | 55.0 | 77.8 | 0 | 80 | 9 |

**Table 2.**
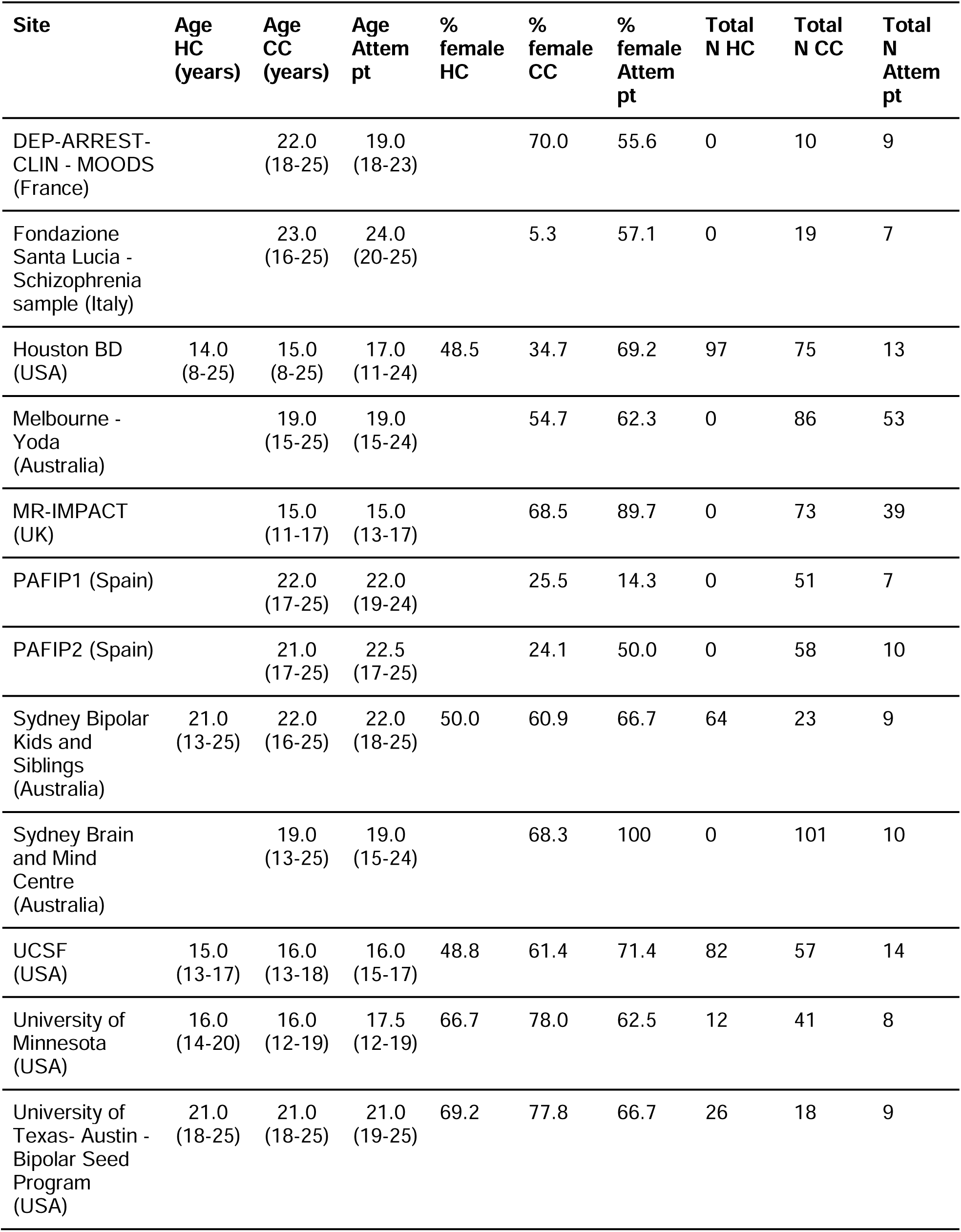
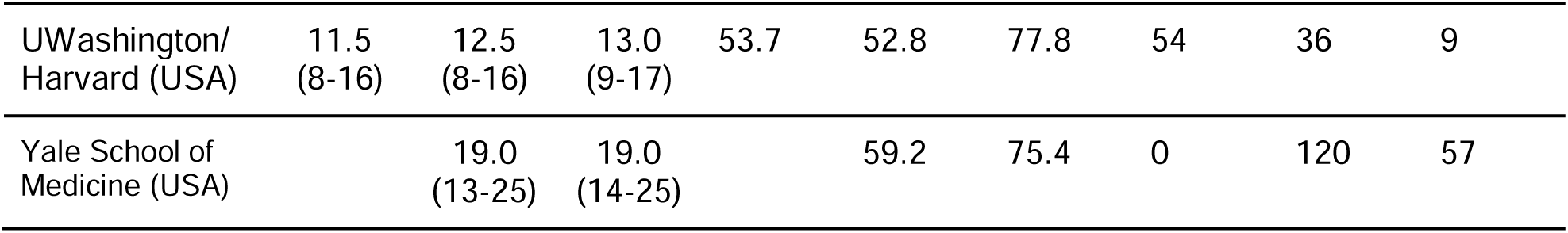
Descriptive statistics for sites included in the attempt analysis. Presented here are age (median, minimum-maximum) and sex for the three groups (HC: healthy controls, CC: clinical controls, attempt: group with lifetime suicide attempt) for the different sites included in the analysis on lifetime history of suicide attempts.

**Figure 1.**
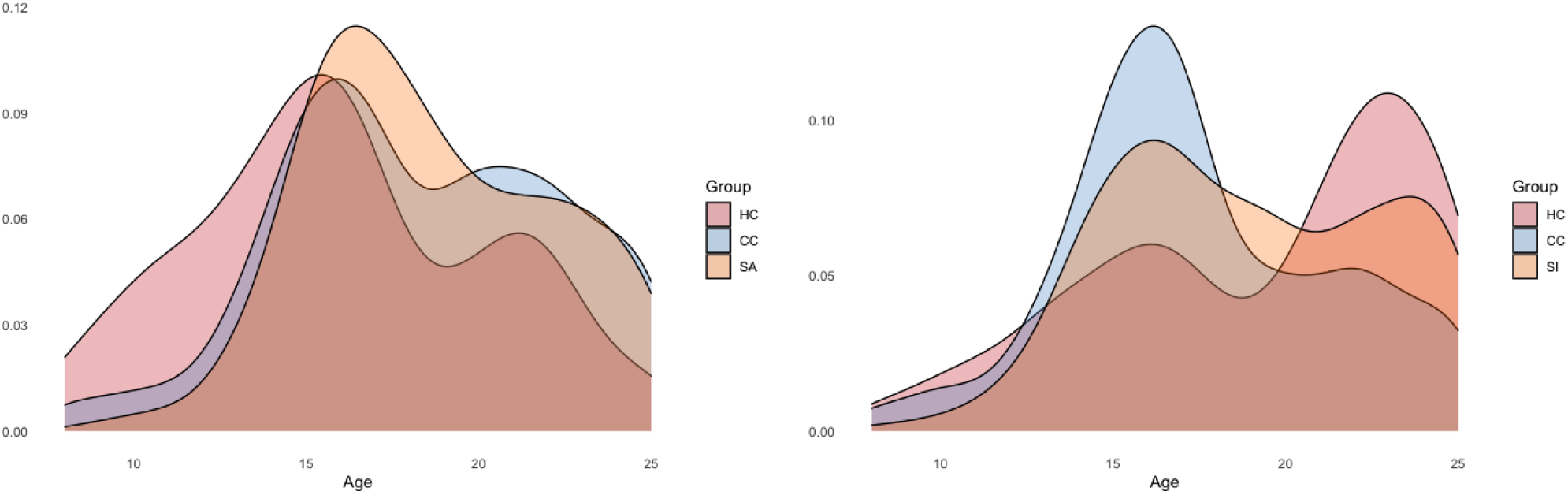
Density plot showing the age distribution per group in the larger ENIGMA-STB sample. In the left panel, the age distribution is shown for the groups in the analysis on lifetime suicide attempt. HC: healthy control group; CC: clinical control group; SA: suicide attempt group. In the right panel, the age distribution is shown for the groups in the analysis on current suicidal ideation. HC: healthy control group; CC: clinical control group; SI: current suicidal ideation group. All analyses are corrected for age, sex, and age-by-sex interaction effects.

**Figure 2.**
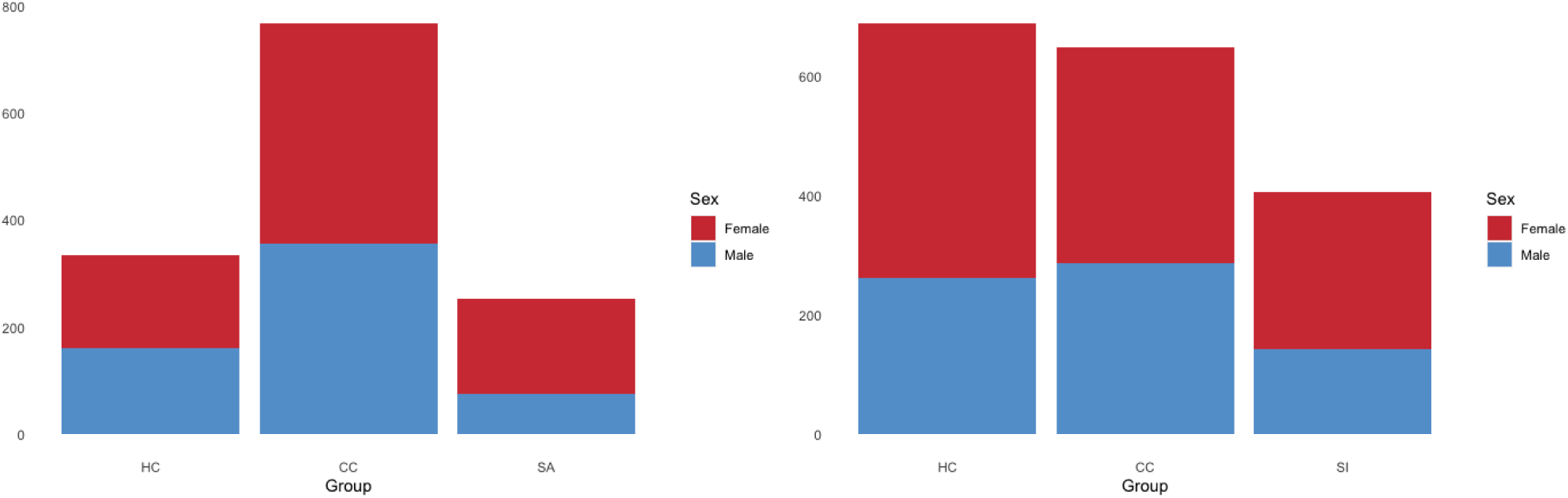
Sex distribution per group. In the left panel, the sex distribution is shown for the groups in the analysis on lifetime suicide attempts in the larger ENIGMA-STB sample. HC: healthy control group; CC: clinical control group; SA: suicide attempt group. In the right panel, the sex distribution is shown for the groups in the analysis on current suicidal ideation. HC: healthy control group; CC: clinical control group; SI: current suicidal ideation group. All analyses are corrected for age, sex, and age-by-sex interaction effects.

### Image processing and harmonization

Structural T1-weighted brain MRI scans were acquired at each site. Information regarding the acquisition parameters, software versions, and scanner characteristics for the different sites is presented in Table S2. The T1-weighted images were analyzed locally using harmonized analysis and quality control protocols for FreeSurfer (18) (http://surfer.nmr.mgh.harvard.edu/), developed by the ENIGMA consortium and freely available (http://enigma.ini.usc.edu/protocols/imaging-protocols/). The ENIGMA FreeSurfer protocol provides tools for quality control of the segmented cortical and subcortical phenotypes. Each site visually inspected the segmentation and excluded regions that were not appropriately segmented. To reduce the number of statistical tests and avoid issues related to left-right flipping that may have occurred at the various sites, we combined regional measures across both hemispheres by taking the mean of the left and right hemisphere regions. We examined the volume of eight subcortical regions and cortical thickness and surface area of 34 regions, defined by the Desikan-Killiany atlas (19). In addition, two global measures were calculated: mean cortical thickness and total surface area across both hemispheres, creating a total of 78 brain measures.

Before the statistical analysis, neuroimaging measures were harmonized across sites using the ComBat algorithm in R (20, 21), with age, sex, and psychiatric diagnosis included as covariates in the model. ComBat uses an empirical Bayes approach to adjust for variability between scanners while still preserving biological variability related to age, sex, and diagnosis. All brain measures included in the statistical analyses were ComBat-corrected. After correction, within-site outliers (measures greater than three standard deviations away from the mean of that region) were excluded from the analysis.

### Statistical analysis

#### Analysis in the Columbia Suicide Severity Rating Scale (C-SSRS) sample

We first examined associations between brain structure and STBs in a subsample of six cohorts, all of which used an instrument designed specifically to assess suicidal ideation and suicide attempt, the C-SSRS (see Table 3). These cohorts included participants with MDD or bipolar disorder (BD) diagnoses (*N*=577, age range 11-25) (healthy control samples were excluded from analyses due to no or limited C-SSRS data). Multiple linear regression analyses were conducted in R, and age, sex and age-by-sex interactions were included as covariates in all analyses. Intracranial volume (ICV) was included as an additional covariate in analyses of subcortical volume and cortical surface area. Because we had estimated and controlled for the contribution of site and scanner using ComBat prior to conducting the analysis, these measures were not included as covariates. In the regression models, the structural brain measures (the average across the left and right hemispheres) were the dependent variables. For suicidal ideation analyses, the continuous C-SSRS measure of recent and lifetime intensity of suicidal ideation were included as predictors. This variable was coded 0-5 (0: no ideation; 1: passive ideation; 2: non-specific active ideation; 3: active ideation with a method, but no plan or intent; 4: active ideation with intent, but no plan; 5: active ideation with a plan and intent). We also examined differences in brain structure between individuals with and without a lifetime history of an actual suicide attempt. Finally, we compared brain morphology between individuals with a lifetime history of suicidal ideation (but no lifetime history of an actual attempt) and those with a lifetime history of an actual attempt. Effect size estimates were calculated using the Cohen’s *d* metric for group comparisons and the standardized beta for associations with the continuous recent or lifetime intensity of suicidal ideation measure.

**Table 3.** Descriptive statistics for studies included in the C-SSRS sample. Presented here are age (median, minimum-maximum) and sex for the six sites in the C-SSRS sample

| Site | Main diagnosis in sample | Age (years) | % Female | Total N |
| --- | --- | --- | --- | --- |
| Melbourne (YODA) | MDD | 19.0<br>range 15-25 | 57.6 | 139 |
| MR-IMPACT | MDD | 15.0<br>range 11-17 | 76.1 | 113 |
| Stanford TAD | MDD | 16.5<br>range 14-18 | 76.2 | 42 |
| Stanford TIGER | MDD | 16.0<br>range 13-18 | 67.6 | 34 |
| UCSF | MDD | 16.0<br>range 13-18 | 63.4 | 71 |
| Yale School of Medicine | MDD + BD | 19.0<br>range: 13-25 | 64.0 | 178 |

MDD: major depressive disorder, BD: bipolar disorder

#### Analysis in the larger ENIGMA-STB sample

We subsequently evaluated whether the significant findings in the C-SSRS sample would be observed in the larger and less homogeneous ENIGMA-STB sample. To this aim, we first restricted the larger ENIGMA-STB sample to individuals with a current or lifetime diagnosis of MDD or BD and healthy controls. In this larger sample, various instruments were used to assess current suicidal ideation and lifetime history of suicide attempts across cohorts. An overview of these instruments is presented in Table S3, and the approach used to harmonize these measures across cohorts is described in Supplemental Note 1. In short, history of lifetime suicidal attempt (yes/no) was determined using either the C-SSRS (22) or the corresponding item from the Kiddie Schedule for Affective Disorders and Schizophrenia (K-SADS) or the Structured Clinical Interview for DSM-IV or DSM-5 (SCID) (23, 24). Current suicidal ideation (in the past week, two weeks or month; yes/no) was determined using the C-SSRS, a diagnostic interview, or items from depression severity rating scale, such as the Beck Depression Inventory (25) or the Hamilton Depression Rating Scale (26).

To investigate whether findings might generalize beyond STBs in mood disorders and to provide additional power to potentially detect any differences not identified in the analyses with the smaller samples, we examined the association between current suicidal ideation and lifetime history of suicide attempt in the overall ENIGMA-STB sample pooled from 21 international cohorts (*N*=2,431). Because only nine sites had information on suicidal ideation and suicide attempt, and previous work in adults has documented differences between the neural correlates of ideation and attempt (27), we conducted separate analyses for suicidal ideation and suicide attempt to optimize the sample size for each analysis. To examine suicidal attempts, we compared three groups: 1) healthy controls, without a current or lifetime psychiatric diagnosis or lifetime history of suicide attempt (‘healthy controls’ *N*=335); 2) ‘clinical controls’, with a current or lifetime psychiatric diagnosis, but no lifetime history of suicide attempt (*N*=768); and 3) ‘clinical attempters’; young people with a current or lifetime psychiatric diagnosis and lifetime history of suicide attempt (*N*=254). To examine current suicidal ideation, we created three groups: 1) HC without a current or lifetime psychiatric diagnosis or lifetime history of suicide attempt or current suicidal ideation (*N*=688); 2) CC with a current or lifetime psychiatric diagnosis but no current suicidal ideation or lifetime history of suicide attempt (*N*=648); and 3) young people with a current or lifetime psychiatric diagnosis and current suicidal ideation, but no lifetime history of suicide attempt (*N*=406). Young people without a psychiatric diagnosis but with a history of lifetime suicide attempts or current ideation were excluded from the analysis, as the sample was too small to analyze separately (*N*=40 and *N*=45, respectively).

Similar to the analyses in the C-SSRS sample, group differences in subcortical volume, cortical thickness, and cortical surface area were compared using multiple linear regression models in R. Because we were specifically interested in differences between individuals with current suicidal ideation or past suicide attempt(s) versus HC or CC, we included a group predictor variable to compare the suicide attempt group to either CC or to HC (in two-group comparisons). In analyses of current suicidal ideation, a group predictor was included to compare the ideation group to either CC or HC. Covariates in the models included age, sex, and age-by-sex interaction. In addition, we corrected for ICV when analyzing subcortical volumes and cortical surface area measures. We calculated effect size estimates using Cohen’s *d* metric.

### Secondary analyses

In addition to evaluating the main effect of current suicidal ideation or a history of suicide attempts, we conducted the above-mentioned analyses in the larger ENIGMA-STB sample separately for males and females, including age and ICV as covariates.

Data on lifetime psychiatric diagnosis were available in a subsample of participants (*N*=655 in the ideation analysis and *N*=842 in the attempt analysis), as some sites only assessed current disorders. Because there were numerous combinations of lifetime diagnoses and comorbidities with low frequencies per combination, we categorized them into six main lifetime diagnosis types: MDD, BD, anxiety disorders, PTSD, OCD, and psychotic disorders. The primary type of diagnosis per group is shown in Figure 3. In secondary analyses, this variable was included as an additional covariate. Given the vast combination of diagnoses and comorbidities and the low number of participants with mental health conditions other than MDD and BD, these analyses were underpowered to perform group-by-diagnosis interaction analyses.

**Figure 3.**
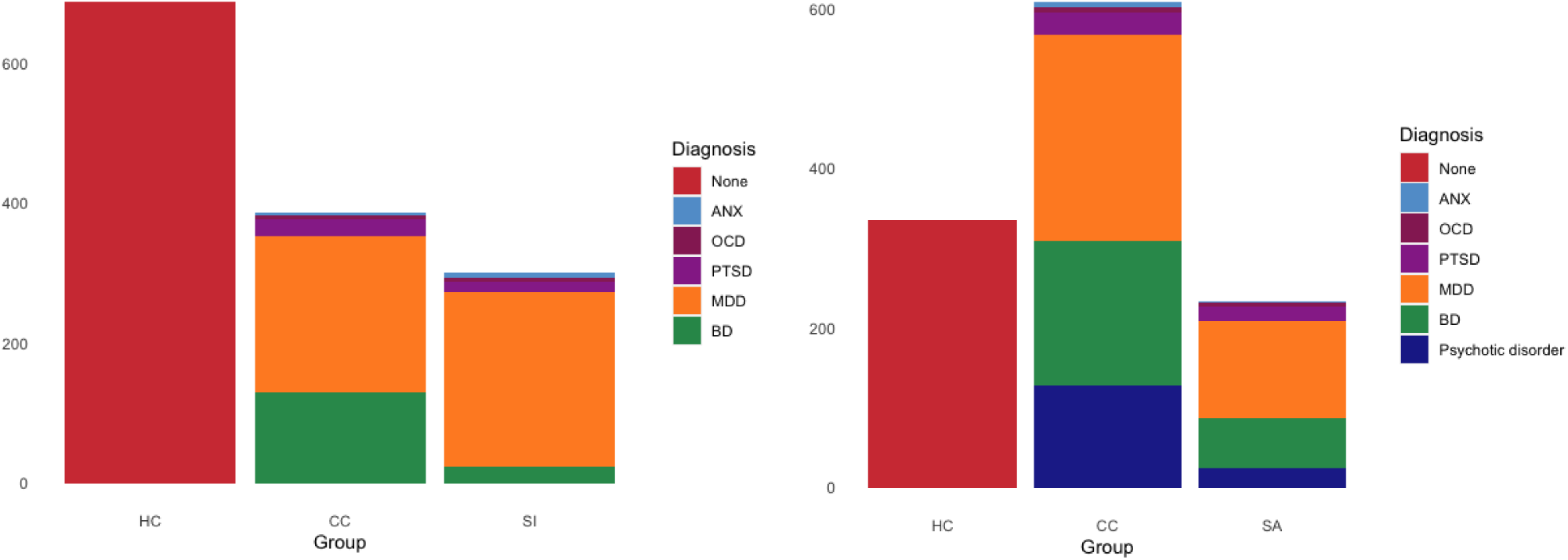
Main type of lifetime diagnosis per group. In the left panel, the main type of diagnosis is shown for the groups in the analysis on current suicidal ideation. HC: healthy control group; CC: clinical control group; SI: current suicidal ideation group. In the right panel, the main type of diagnosis is shown for the groups in the analysis on lifetime suicide attempt. HC: healthy control group; CC: clinical control group; SA: suicide attempt group; ANX: anxiety disorders; MDD: major depressive disorder; BD: bipolar disorder; OCD: obsessive-compulsive disorder; PTSD: post-traumatic stress disorder.

All p-values were corrected for multiple comparisons (for the 78 brain measures) using the Benjamini Hochberg correction in R to ensure an FDR<0.05.

## Results

### Associations with suicidal ideation and attempts in the C-SSRS sample

There were no significant associations between lifetime or recent intensity of ideation and cortical thickness, cortical surface area, and subcortical volume measures (*N*=438 and 510 respectively; Table S4 and S5).

The surface area of the frontal pole was smaller in young people with a lifetime history of a prior suicide attempt (*N*=168) than in those without (N=407; FDR p-value < .001; Cohen’s d: -0.334; Table S6 and Figure 4). Finally, there were no significant differences between those with lifetime ideation (but no history of a prior suicide attempt) (*N*=200) and those with a lifetime history of an attempt (*N*=168; Table S7).

**Figure 4.**
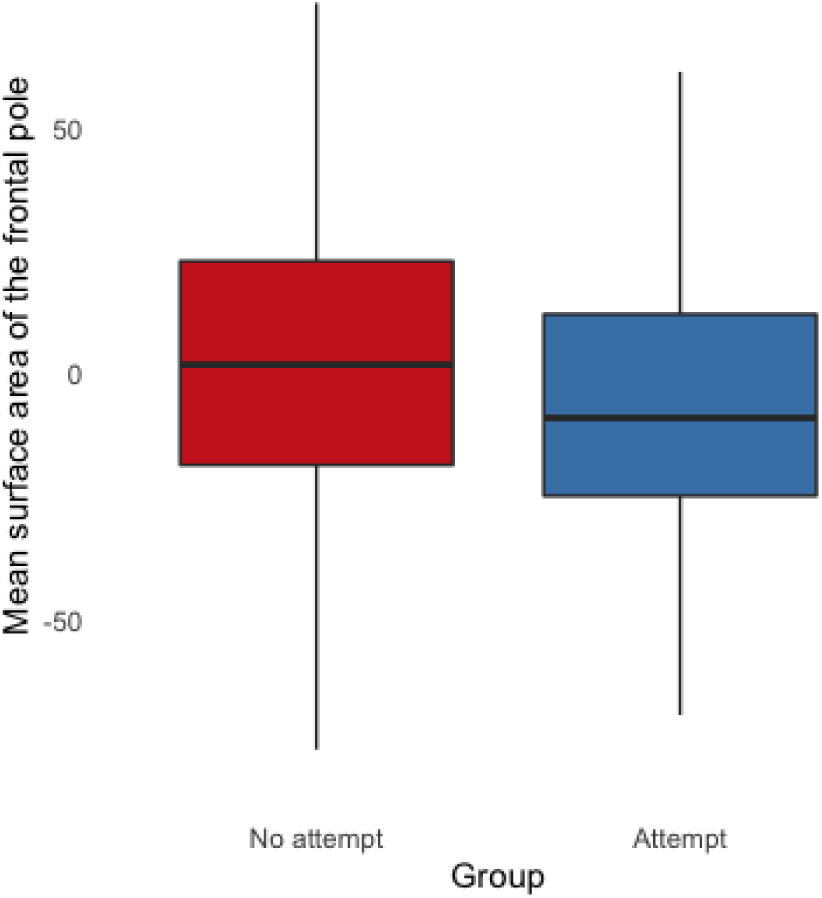
Boxplot showing the mean surface area of the frontal pole in young people without a lifetime history of an actual suicide attempt (in red), and those with a lifetime history of an actual suicide attempt (in blue). Lifetime history of an actual suicide attempt was assessed using the C-SSRS.

### Associations with current suicidal ideation and history of attempt in the larger ENIGMA-STB sample

In the larger ENIGMA-STB sample of participants with a diagnosis of MDD or BD and healthy controls (*N* HC=335, *N* CC=545, *N* attempt=225), surface area of the frontal pole was smaller in the attempt group than in the CC (Cohen’s d = -0.22, uncorrected-*p* = 0.005) and HC (Cohen’s d = -0.18, uncorrected-*p* = 0.035) groups.

#### Current suicidal ideation

In the larger transdiagnostic ENIGMA-STB sample (not restricted to MDD or BD diagnosis) no brain structure measure differed significantly between young people with current suicidal ideation (*N*=406) and HC (*N*=688; Table S8) or CC (*N*=648; Table S9) groups. No differences were observed when additionally adjusting for primary diagnosis (Table S10), nor when conducting separate analyses in males and females (*N* HC=187, *N* CC=153, *N* ideation=116 in males; *N* HC=383, *N* CC=322, *N* ideation=219 in females; Tables S11, S12, S13, and S14).

#### Lifetime history of suicide attempt

In the larger transdiagnostic ENIGMA-STB sample (not restricted to MDD or BD diagnosis), MRI measures also did not differ significantly between the suicide attempt group (*N*=254) and HC (*N*=335; Table S15) or CC (*N*=768; Table S16) groups. No differences were observed when correcting for primary diagnosis (Table S17) or conducting separate sex-stratified analyses (*N* CC=170, *N* attempt=45 in males; *N* HC=174, *N* CC=398, *N* attempt=174 in females; Tables S18, S19, and S20).

## Discussion

In this study we examined the associations between STBs and structural MRI measures in young people in a combined sample from the ENIGMA-STB consortium. In a more homogeneous subsample (*N*=577) including participants from six cohorts assessed with the same well-validated instrument specifically developed to assess STBs (the C-SSRS) and including only young people with MDD or BD (age range 11-25 years), we found a significantly smaller surface area in the frontal pole in those with, compared to without, a lifetime history of suicide attempts. We also found a lower surface area in the frontal pole in suicide attempters compared to CCs in the larger ENIGMA-STB sample restricted to individuals with MDD or BD and healthy controls, but in which different assessments for STBs were used across sites. The effect size in the C-SSRS sample was larger than the effect size in this larger ENIGMA-STB sample (Cohen’s d = -0.334 versus -0.22). The larger effect size for the association between attempt and surface area of the frontal pole in the C-SSRS sample compared to the larger ENIGMA-STB sample could be related to any of several sources of heterogeneity, including the more specific and consistent definition used for suicide attempts in the C-SSRS group and clinical heterogeneity.

The frontal pole is the rostral-most aspect of the prefrontal cortex and plays an essential role in higher-order functions involved in emotion and other behavioral regulation, notably, decision-making and cognitive inhibition, and social cognition processes (e.g., self-referential processes) implicated in STBs (28–31). The development of surface area and cortical thickness are genetically independent and are the result of different neurobiological mechanisms (32); thus, they may represent different features of development and aging. Because cortical surface area is highly heritable (33) and is less affected by environmental factors during development and in later life, than is the cortical thickness (34), alterations in frontal pole surface area may represent a pre-existing vulnerability for suicidal behavior in adolescents. Longitudinal studies are needed to elucidate whether structural alterations, in particular cortical surface area, in this region precede the onset of STBs. A previous longitudinal study, structural alterations in the frontal pole (amongst other frontal regions) was associated with a family history of bipolar disorder, which is also associated with increased risk of developing STBs (35). In another longitudinal study of a sample of 46 adolescents and young adults with mood disorders, decreases in rostral prefrontal volume were found to be associated with future suicide attempts, although thickness and surface area were not studied separately (9). Together with the findings of this study, results suggest that decreases in rostral PFC surface area warrant further study as potential predictors of and targets for the prevention of suicide. Alterations in functional connectivity between the rostral prefrontal cortex from an amygdala seed region have also previously been observed in young people with a history of suicide attempt (6). While future studies are needed to directly assess structural and functional associations, this suggests that the structural abnormalities observed herein may be related to functional dysconnectivity in brain systems that subserve behaviors such as emotional regulation implicated in STBs.

The absence of a significant finding (besides the frontal pole) in the larger, more heterogeneous sample is potentially consistent with prior reports in the literature. A prior ENIGMA-MDD study found significant differences in brain structure in adults with major depressive disorder and a history of suicide attempts (17). However, the sample size of that study was larger (18,925 participants of whom 694 had attempted suicide, compared to 1,357 participants of whom 254 attempted suicide in the current study), so we may still have been underpowered to detect these small effect sizes. In addition, the previous ENIGMA-MDD suicide study focused on adults (versus young people in the current study) and included only people with MDD and HC, whereas here, we included a transdiagnostic sample. Thus, in the larger overall sample of this study, more substantial heterogeneity was introduced by including a variety of mental health conditions. A recent study that examined the association between STBs and brain structure in over 6,000 younger children aged 9-10 years in the Adolescent Brain Cognitive Development (ABCD) study did not reveal significant structural alterations on the frontal cortex in association with STBs (36). This may be related to the fact that the ABCD study is a general population sample with only a few children diagnosed with mood disorders, and STBs were less common and severe. Given the important role of puberty-related developmental processes in STBs, this sample may have been too young to detect these brain alterations (37). Moreover, the rostral frontal cortex is one of the last brain regions to mature. It is still undergoing dynamic changes during childhood, adolescence, and young adulthood (38), which may also interact with other sources of heterogeneity in the sample, such as sex, childhood abuse, family history of suicide attempts, and psychopathology.

Our findings highlight the importance in the study of the neurobiology of STBs of considering sources of heterogeneity in the methods and in the demographic and clinical features of the sample. The study shows the importance of using well-validated and detailed phenotyping of STBs, such as the C-SSRS, when pooling data. Therefore, a strength of this study includes the large sample sizes that allowed the examination of more detailed and homogeneous phenotypes. An additional strength of the study is the use of harmonized protocols for image processing and quality control. We should note the limitations of this study. First, different instruments were used to assess STBs across cohorts for analyses in the larger ENIGMA-STB sample, although we used a detailed process to harmonize measures across studies. Moreover, when multiple instruments were used to assess suicidal ideation (thoughts) or attempts within one cohort, we defined STBs in that sample using instruments that showed strong correlations with the instruments used by other cohorts to assess STBs (39). Future multi-site collaborations would be improved by prospective harmonization in data collection and/or measurement. A second limitation was the cross-sectional study design. Although the findings are consistent with a prior report on future suicide attempts (9), we cannot determine whether brain structure increases the risk for STBs or whether prior attempts affect brain structure. It is important to note that segmentation of the surface area of the frontal pole showed moderate test-retest reliability (40). Finally, while including participants from many international studies, the samples mainly included Caucasian participants from high-income countries.

In conclusion, by harmonizing neuroimaging data from research groups worldwide, we found that a deficit in the surface area of the frontal pole was related to suicide attempts in young people, which we suggest represents a pre-existing vulnerability to suicide attempts. Future studies focusing on the frontal pole may elucidate the functional and structural neurobiological mechanisms through which this region contributes to the development of STBs in young people and is a promising candidate to be a target for the early prevention of suicide.

## Data Availability

NA

## Acknowledgments

This work was supported by the MQ Brighter Futures Award MQBFC/2 (LS, LC, LV, MRD, LvV, ALvH, HB) and the U.S. National Institute of Mental Health under Award Number R01MH117601 (LS, LvV, NJ). LvV received funding through the National Suicide Prevention Research Fund, managed by Suicide Prevention Australia. LS is supported by an NHMRC Career Development Fellowship (1140764). ALvH is funded through the Social Safety and Resilience programme of Leiden University. SA, NB, FP, and GS acknowledge that data collected in IRCCS Santa Lucia Foundation, Rome, Italy was funded by a study funded by the Italian Ministry of Health grant RC17-18-19-20-21/A. ZB, KC, B K-D acknowledge data collected at the University of Minnesota was funded by the National Institute of Mental Health (K23MH090421), the National Alliance for Research on Schizophrenia and Depression, the University of Minnesota Graduate School, the Minnesota Medical Foundation, and the Biotechnology Research Center (P41 RR008079 to the Center for Magnetic Resonance Research), University of Minnesota, and the Deborah E. Powell Center for Women’s Health Seed Grant, University of Minnesota. HB acknowledges data collected at the Yale School of Medicine, New Haven, CT, USA, was funded by: MQ Brighter Futures, R61MH111929RC1MH088366, R01MH070902, R01MH069747, American Foundation for Suicide Prevention, International Bipolar Foundation, Brain and Behavior Research Foundation, For the Love of Travis Foundation and Women’s Health Research at Yale. LC is supported by Interdisziplinäres Zentrum für Klinische Forschung, UKJ. BCD was funded by a CJ Martin Fellowship (NHMRC app 1161356). BCD research leading to these results has received funding from the program “Investissements d’avenir” ANR-10-IAIHU-06. CGD and BJH acknowledge that data collected in Melbourne, Australia, was supported by Australian National Health and Medical Research Council of Australia (NHMRC) Project Grants 1064643 (principal investigator, BJH) and 1024570 (principal investigator, CGD). BJH and CGD were supported by NHMRC Career Development Fellowships (1124472 and 1061757, respectively). UD and TH acknowledge data collected at the FOR2107-Münster was funded by the German Research Foundation (DFG, grant FOR2107-DA1151/5-1 and DA1151/5-2 to UD, and DFG grants HA7070/2-2, HA7070/3, HA7070/4 to TH). AJ and TK acknowledges data collected at the FOR2107-Marburg was funded by the German Research Foundation (DFG, grant FOR2107-JA 1890/7-1 and JA 1890/7-2 to AJ, and DFG, grant FOR2107-KI588/14-1 and FOR2107-KI588/14-2 to TK). KD acknowledges data collected for the Münster Neuroimaging Cohort was funded by the Medical Faculty Münster, Innovative Medizinische Forschung (Grant IMF KO 1218 06 to KD). JMF, PBM, BJO, and GR acknowledge that the “Kids and Sibs” Study was supported by the Australian National Medical and Health Research Council (Program Grant 1037196 and Investigator Grant 1177991 to PBM, Project Grant 1066177 to JMF), the Lansdowne Foundation, Good Talk and the Keith Pettigrew Family Bequest (PM). JMF gratefully acknowledges the Janette Mary O’Neil Research Fellowship. IHG is supported in part by R37MH101495. Support for TAD comes from the National Institute of Mental Health (K01MH106805). TH acknowledges support for TIGER includes the Klingenstein Third Generation Foundation, the National Institute of Mental Health (K01MH117442), the Stanford Maternal Child Health Research Institute, and the Stanford Center for Cognitive and Neurobiological Imaging. TCH receives partial support from the Ray and Dagmar Dolby Family Fund. KAM, ABM, MAS acknowledge data collected at Harvard University was funded by the National Institute of Mental Health (R01-MH103291). IN is supported by grants of the Deutsche Forschungsgemeinschaft (DFG grants NE2254/1-2, NE2254/3-1, NE2254/4-1).This study was supported by the National Center for Complementary and Integrative Health (NCCIH) R21AT009173 and R61AT009864 to TTY; by the National Center for Advancing Translational Sciences (CTSI), National Institutes of Health, through UCSF-CTSI UL1TR001872 to TTY; by the American Foundation for Suicide Prevention (AFSP) SRG-1-141-18 to TTY; by UCSF Research Evaluation and Allocation Committee (REAC) and J. Jacobson Fund to TTY; by the National Institute of Mental Health (NIMH) R01MH085734 and the Brain and Behavior Research Foundation (formerly NARSAD) to TTY. YC acknowledges the Medical Leader Foundation of Yunnan Province (L2019011) and Famous Doctors Project of Yunnan Province Plan (YNWR-MY-2018-041). DTG, BCF and RAA wish to thank all PAFIP patients and family members who participated in the study as well as PAFIP’s research team and Instituto de Investigación Marqués de Valdecilla. Work by the PAFIP group has been funded by Instituto de Salud Carlos III through the projects PI14/00639, PI14/00918 and PI17/01056 (Co-funded by European Regional Development Fund/European Social Fund “Investing in your future”) and Fundación Instituto de Investigación Marqués de Valdecilla (NCT0235832 and NCT02534363). MER received support from the Australian National Health and Medical Research Council (NHMRC) Centre for Research Excellence on Suicide Prevention (CRESP) [GNT1042580]. ETCL is supported by grants from NIAAA (K01AA027573, R21AA027884) and the American Foundation for Suicide Prevention. All authors thank the participants for volunteering their time and supporting our research.

