## Supplements for "Structural brain alterations associated with suicidal thoughts and behaviors in young people: results across 21 international studies from the ENIGMA Suicidal Thoughts and Behaviours consortium"

**Supplemental tables**

Table S1. Main diagnosis of participants and study exclusion criteria in the different sites

| **Site** | **Main diagnosis participants** | **Inclusion criteria** | **Exclusion criteria** |
| --- | --- | --- | --- |
| EPISCA | MDD, PTSD | For patients: aged between 12 and 19, estimated full scale IQ>80, right handed, normal or corrected to normal vision, sufficient understanding of the Dutch language, no history of neurological impairments and no contraindications for MRI.  Having a depressive or anxiety disorder as classified by DSM-IV criteria (including PTSD), no current or prior use of antidepressants.  For healthy controls:  aged between 12 and 19, estimated full scale IQ>80, right handed, normal or corrected to normal vision, sufficient understanding of the Dutch language, no history of neurological impairments and no contraindications for MRI.  No current or passed DSM-IV classifications, no clinical scores on validated mood and anxiety questionnaires, no history of traumatic experiences, and no current psychotherapeutic and/or psychopharmacological intervention of any kind | For both patients and healthy controls: primary DSM-IV clinical diagnosis of ODD, CD, pervasive developmental disorder, Tourette's syndrome, OCD, bipolar disorder, and psychotic disorder, current substance use, history of neurological disorders or severe head injury, and pregnancy |
| University of Texas - Austin - Bipolar Seed Program | BD | For patients: Meeting diagnostic criteria for bipolar disorder type I per the Structured Clinical Interview for DSM-5-Research Version, between 18-25 years of age | For patients: History of major medical illness or significant head trauma resulting in possible neurological or  central nervous system damage, Full-scale IQ <85, Positive pregnancy test, Having a medical condition/previous surgery preventing participation in a magnetic resonance  imaging scan  Not taking medications for greater than or equal to 4 weeks (i.e., participants must be stable on their medications)  For healthy controls: Meeting criteria for psychotic, affective or other anxiety disorders per the Structured Clinical  Interview for DSM-5-Research Version, History of major medical illness or significant head trauma resulting in possible neurological or  central nervous system damage, Full scale IQ <85, Positive pregnancy test, Having a medical condition/previous surgery preventing participation in a magnetic resonance  imaging scan |
| Boystown | MDD, GAD, SAD, PTSD | For patients: Demonstrated SUI during the first 90 days at a residential treatment center | For patients & healthy controls: IQ<75, Pregnancy, Ongoing medical illness other than those listed in the inclusion criteria for the respective groups that require use of any medication that may have psychotropic effects such as beta blockers or steroids Explicit exclusions include active psychosis, Pervasive Developmental Disorders and Tourette’s syndrome  Neurologic disorder (including seizures).  Any metallic objects in the body. Metal plates, certain types of dental braces, cardiac pacemakers, ext., that are sensitive to electromagnetic fields contraindicate MRI scans  Claustrophobia  In addition, for our healthy SUI controls, no psychiatric diagnoses are allowed. |
| FOR2107-Marburg | MDD | For patients: age 18-65 years; patients were diagnosed by SCID-Interview with major depressive  disorder (currently depressed or remitted) or with bipolar disorder  (currently  depressed, (hypo)manic or remitted)  For healthy controls: age  18-65 years | Exclusion criteria all: any MRI contraindications; any neurological abnormalities. Exclusion criteria controls: any current or former psychiatric disorder; Exclusion criteria patients:  substance dependence or current benzodiazepine treatment (wash out of at  least three half-lives  before study  participation)" |
| FOR2107-Münster | MDD | For patients: age 18-65 years; patients were diagnosed by SCID-Interview with major depressive  disorder (currently depressed or remitted) or with bipolar disorder (currently  depressed, (hypo)manic or remitted)  For healthy controls: age 18-65 year | Exclusion criteria all: any MRI contraindications; any neurological abnormalities. Exclusion criteria healthy controls: any current or former psychiatric disorder; Exclusion criteria patients: substance dependence or current benzodiazepine treatment (wash out of at least three half-lives before study participation) |
| Houston BD | BD | NA | Healthy controls reporting current or past Axis I disorders, suicidal history and a first-degree relative with any Axis I disorder were excluded. Participants with any endocrinological disease, head trauma, neurological disease, and family history of any hereditary neurological disorder or medical conditions such as hypertension, diabetes, active liver disease and kidney problems were excluded from this study. |
| MDD Cohort | MDD | For patients: right-handedness, Han Chinese background, between 18-60 years, HDRS scores of 17 or greater, first episode MDD and untreated  For healthy controls: matched on age, sex and years of education with the MDD group | For patients: other axis-I disorders, organic brain disease or neurological disorders, delusions or hallucinations, any physical illness, conditions that cause cerebral atrophy (hypertension, diabetes, stroke), MRI contraindications.  For healthy controls: psychiatric, neurological or organic diseases, first degree family member  with psychiatric illness |
| University of  Minnesota | MDD | Inclusion criteria: age 12-19  For patients: primary diagnosis of MDD  For healthy controls: no current of past psychiatric diagnosis | Exclusion criteria for  included the presence of a  neurologic or other  chronic medical condition, mental retardation, intellectual disability, pervasive developmental disorder, substance use disorder, bipolar disorder, or schizophrenia |
| Muenster Neuroimaging Cohort | MDD | Inclusion criteria: age 17-65 years; patients were diagnosed of MDD by SCID-Interview, HAMD >= 18); | For patients: presence of bipolar disorder, schizoaffective disorders and schizophrenia; substance-related disorders or current benzodiazepine treatment (wash out of at least three half-lives before study participation), and former electroconvulsive therapy. For controls: any current or former psychiatric disorder. For patients & controls: any neurological abnormalities, MRI  contra-indications |
| Yale School of Medicine | BD, MDD | For patients: Ages 13-25 years with major depressive disorder  (MDD) or bipolar disorder (BD) type 1 or type 2 determined by DSM-IV. | Exclusion criteria for patients: history of or  current medical or neurological condition that could affect the brain (except treated hypothyroidism), and magnetic resonance imaging (MRI) contraindications. |
| MR-IMPACT | MDD | Patients: age 11-17, current DSM-IV unipolar MDD diagnosis determined using child and parent interviews using the KSADS-PL, score of 27 or higher on the MFQ | Exclusion criteria for all participants: alcohol dependence, drug dependence, pervasive developmental disorder or generalized learning problem, pregnancy or breastfeeding, current medication use that could adversely interaction with SSRIs, MRI  contraindications, brain  abnormalities, intolerance to the MRI environment |
| SOCAT | MDD | Patients: DSM-5 diagnosis of MDD  Healthy participants: no history of psychiatric illness | For all participants: MRI contraindications, history of neurodevelopmental diseases, |
| Stanford TAD | MDD | For patients: DSM-IV current MDD diagnosis | For patients: Diagnosis of MDD, psychotic disorder, substance dependence,  contraindications for  scanning, lifetime history of neurological, cardiovascular or other major medical problems. |
| Sydney Brain and Mind Centre | MDD + OCD + BD + Psychosis + GAD + SAD | Patients: Age 12 - 25 and presented with an affective, psychotic or developmental/behavioural syndrome. | Exclusion criteria for all subjects included medical instability (as determined by a psychiatrist), history of neurological disease (e.g. tumour, head trauma, epilepsy), medical illness known to impact cognitive  and brain function (e.g. cancer), intellectual and/or developmental disability and insufficient English for neuropsychological assessment. All subjects were asked to abstain from drug or alcohol use for 48 hours prior to testing and informed about a drug screen protocol. |
| UCSF | MDD | All: between 13 and 18 years of age. Right handed. Particpants were well matched on age, sex, socioeconomic status, and pubertal status. MDD: Diagnosis of MDD based on KDADS-PL. HC: No diagnosis of psychiatric disorder based on Diagnostic Interview Schedule for Children and Diagnostic Predictive Scale. | Exclusion criteria for all participants included: 1) use of pharmacotherapeutics for treating psychiatric conditions within the past 6 months, 2) misuse of drugs within two months prior to MRI scanning; 3) two or more alcoholic drinks per week within the previous month (as  assessed by the  Customary Drinking and Drug Use Record; CDDR) (Brown et al, 1998); 4) a full scale IQ score of less than 75 (as assessed by the Wechsler Abbreviated Scale of Intelligence; WASI) (Wechsler, 1999); 5) contraindications for  MRI including ferromagnetic implants and claustrophobia; 6)  pregnancy or the  possibility of pregnancy; 7) left-handedness; 8) prepubertal status (as assessed as Tanner  stages of 1 or 2) (Tanner, 1962); 9) inability to understand and comply with procedures; 10) neurological disorder (including meningitis, migraine, or HIV); 11) head trauma; 12) learning disability; 13) serious  health problems; and 14) complicated or premature birth (i.e., birth before 33 weeks of gestation). The MDD group was subject to the additional exclusion criterion of a primary psychiatric diagnosis other than MDD. The healthy control group was subject to the additional exclusion criteria of: 1) history of mood or psychotic disorders in a first- or second-degree relative (as assessed by the Family Interview for Genetics; FIGS) (Maxwell, 1992); and 2) current or lifetime DSM-IV-TR Axis I psychiatric disorder. |
| UWashington/  Harvard | MDD + PTSD + GAD + PD + SAD | For all participants: 8-20 years of age, English speaking, with and without exposure to trauma.  For patients: Endorsement  of ideation or behaviour on the SITBI | Exclusion criteria for all participants: Psychiatric medication use (stimulants for ADHD were  discontinued for the scan),  MRI contraindications, active substance dependence, pervasive developmental disorder. For healthy controls: Endorsement of ideation or behaviour on the SITBI |
| Melbourne  (YODA) | MDD | For patients: aged 15–25 years; a diagnosis of MDD based on the Structured Clinical Interview for DSM-IV Axis I Disorders (SCID-IV); a score of 20 or higher on the Montgomery-Åsberg Depression Rating Scale (MADRS); and able to provide written informed consent (including adequate intellectual capacity and fluency in English). | For patients: lifetime or  current SCID-I diagnosis of psychotic disorder, or bipolar I or II disorder. For  healthy control subjects:  any SCID-I diagnosis or medication use. For both groups: Acute or unstable medical disorder; general MRI contraindications |
| DEP-ARREST-CLIN - MOODS | MDD | For patients: 18-65 years, current MDD diagnosis, minimum HDRS scores of 18, antidepressant-free at least one month prior to study inclusion  For healthy controls: absence of current or past psychiatric disorders or somatic conditions (including nasal polyposis and acute sinusitis or rhinitis) | For patients: diagnosis of bipolar disorder, psychotic disorder, eating disorder or addiction according to the DSM-5  For all participants: pregnancy or breastfeeding, nasal polyposis, acute or chronic sinusitis or rhinitis. |
| PAFIP 1 & 2 | Psychosis | For patients: 15-50 years, DSM-IV diagnosis of schizophrenia, schizophreniform disorder, schizoaffective disorder, delusional disorder, brief reactive psychosis or psychosis not otherwise specified  For healthy controls: no history of mental illness treatment and no axis-I disorder | For patients: history of neurological disease, head injury, mental retardation or drug dependence.  For healthy controls: current or past psychiatric diagnosis, mental retardation, neurological or general medical illnesses. Psychosis in first-degree relatives |
| Sydney Bipolar Kids and Siblings | BD | For patients: Any DSM-IV diagnosis  For all participants: age  between 12-30 years | For healthy controls: parent or sibling with bipolar I or II, recurrent MDD, schizoaffective disorder, schizophrenia,  recurrent substance  abuse, psychiatric hospitalization. Parent with a first degree relative who had a past mood disorder hospitalization or history of psychosis  For all participants: family members |
| Fondazione Santa Lucia - SZ sample | Psychosis | For patients: DSM-5 diagnosis  For all participants: age  between 18-65 years | Exclusion criteria for all participants: history of alcohol or drug abuse  within past 2 years,  lifetime drug dependence, traumatic head injury with loss of consciousness, MRI contraindications, past or present major medical illness or neurological disorder, mental retardation, brain abnormalities, vascular lesions.  For controls: current or lifetime DSM-V mental or personality disorder according to SCID, first-degree relative with psychosis, mental retardation, minor or major DSM-5 major neurocognitive disorder, any brain abnormality or vascular lesions |
| Stanford TIGER sample | MDD | For patients: DSM-IV criteria for MDD or Dysthymia OR if at threshold based on DSM-IV, CDRS-R t-score > 55  For controls: no present or lifetime diagnosis of Axis I based on DSM-IV | Exclusion criteria for patients: prepubertal status; younger than age 13; older than 18; lifetime or current criteria for Mania, Psychosis, or Alcohol Dependence (based on DSM-IV) or Moderate Substance Use Disorder with substance-specific threshold for withdrawal (based on DSM-5); concussion within the past 6 weeks or history of concussion with any loss  of consciousness;  contraindications to MRIs; serious neurological or intellectual disability that could impede participant ability to complete study components  For controls: prepubertal status; younger than age 13; older than 18; first-degree relative with (suspected) depression, suicide, psychosis, or mania; concussion within the past 6 weeks or history of concussion with any loss of consciousness; contraindications to MRIs; serious neurological or intellectual disability that could impede participant ability to complete study components |

Table S2. Scanner type, T1 scan acquisition parameters and FreeSurfer version per site

| **Site** | **Scanner vendor and type** | **Acquisition parameters** | **Freesurfer version** | **Slice orientation** | **Operating system** |
| --- | --- | --- | --- | --- | --- |
| EPISCA | 3T Philips Achieva | 3-dimensional gradient-echo T1-weighted image TR = 9.8 ms; TE = 4.6 ms; flip angle = 8°; 140 sagittal slices; no slice gap; FOV =256 × 256 mm; 1.17 × 1.17 × 1.2 mm voxels; duration = 4:56 min | 5.3 | Sagittal | Ubuntu 14.04.5 LTS (Linux 3.13.0-153-generic x86_64) |
| University of Texas - Austin -Bipolar Seed Program | 3T Siemens Skyra | TR=1900ms, TE=2.42ms, matrix=224x224, FOV=220x220mm2; 192 slices; 1 mm thickness; 1x1x1 mm | 5.3 | Sagittal | Linux |
| Boystown | 3T Siemens Skyra | MPRAGE, TR=2200 ms; TE=2.48 ms; 230 mm FOV; 8o flip angle; 256x208 matrix; 176 slices; 1 mm thickness; 0.9x0.9x1mm voxel size | 5.3 | Axial | CentOS 6.9 (Linux 2.6.32-754.12.1.el6.x86_64) |
| FOR2107-Marburg | 3T Siemens Magnetom TiroTim syngo MR B17 | 3D T1-weighted MPRAGE  TR=1900 ms, TE=2.26 ms, TI=900ms, fip angle=9 degrees, 176 slices, 0.5mm slice gap, voxel size: 1x1x1mm. | 5.3 | Sagittal | Red Hat Enterprise Linux Server release 5.11 (Tikanga) |
| FOR2107-Münster | 3T Siemens PRISMA | 3D T1-weighted MPRAGE  TR=1900ms, TE=2.28ms, TI=900 ms, flip angle=8 degrees, 192 slices, 0.5mm gap, voxel size:1x1x1 | 5.3 | Sagittal | Red Hat Enterprise Linux Server release 5.11 Tikanga |
| Houston BD | 1.5 T Philips Medical Systems Gyroscan Intera  1.5 T Philips Medical Systems Gyroscan Intera | T1 weighted fast field echo sequence (3D T1-FFE) TR = 25 ms, TE = 5 ms,  FOV = 240 mm × 220 mm, gap = 0,  and matrix size = 256 × 256.  MPRAGE  TR=24 ms, TE=5ms, voxel size=1x1x1mm, slice gap=1mm, 144 slices | 5.3  5.3 | Sagittal  Sagittal | Fedora19  Fedora19 |
| MDD Cohort | 3T Philips Achieva MRI scanner | TR = 7.38 ms, TE = 3.4 ms, matrix size = 256 mm × 256 mm, FOV = 250 mm × 250 mm, flip angle = 8º, slice thickness = 0.6 mm, slices = 230 with no gap, acquisition time = 6 min 53 s. | 6.0 | Axial | Linux Ubuntu (12.04) |
| University of Minnesota | 3T Siemens TIM Trio | TR = 2530ms; TE = 3.65ms; TI = 1100ms; flip angle = 7 degrees; FOV = 256; GRAPPA =  2; 224 slices 1mm thickness; 1x1x1  mm | 5.3 | Coronal | CentOS 6 |
| Muenster Neuroimaging Cohort | 3T Philips Gyroscan Intera | 3D fast gradient echo sequence, TR = 7.4 ms, TE = 3.4 ms, flip angle = 9°, FOV of 256 × 204 x 160, voxel size .5 mm × .5  mm × .5 mm | 5.3 | Sagittal | Red Hat Enterprise Linux Server release 5.11 Tikanga |
| Yale School of Medicine | 3T Siemens Trio scanner | 3D-MPRAGE, TR=1500 ms, TE=2.83 ms (65%) or 2.77 ms (35%), TI=700, flip angle=15°, matrix=256 × 256, FOV= 256 × 256 mm^2^, 160 slices of 1 mm without gap, voxel size= 1 mm^3^ | 6.0 | Sagittal | Linux Centos, version 7 |
| MR-  IMPACT | 3T Siemens Magnetom Trio Tim | 3D-MPRAGE  TR=2.3 ms, TE=2.98 ms, TI=900 ms, flip angle=9 degrees, FOV: 240x256, voxel size=1x1x1mm, 176 slices, 1mm thickness | 6.0 | Sagittal | Linux |
| SOCAT | 3T Siemens Magnetrom Verio | 3D T1-weighted sequence, TR=1600, TE=2.2, TI=900, FOV: 160 x 512 x 512, 1mm thickness, voxel size 1x0.5x0.5 | 6.0 | Sagittal | Ubuntu 18.04 LTS |
| Stanford TAD | 3T GE Discovery  MR750 | TR=6.24 ms,  TE=2.344 ms, flip angle =12⁰, voxel size=0.8984 x 0.8984 x 0.9000 mm, FOV: 230 x 230 mm | 6.0 | Sagittal | MacOS |
| Sydney Brain and Mind Centre | 3T GE MR750 | TR=7.2 msec; TE=2.78 msec; matrix =256; FOV=240; Number of slices=196; thickness=0.9mm | 5.1 | Coronal | Linux_Ubuntu12.04_64 |
| UCSF | 3T GE MR750 | TR/TE 8.1 Mx/3.17 ms, flip angle 12 degrees. 256 by 256 matrix, 168 slices; 1x1x1mm | 5.3 | Sagittal | Scientific Linux 6 |
| UWashington/  Harvard | 3T Philips  Achieva | MPRAGE,  TR=2530ms, TE=3.5ms, 7 degree Flip angle 256x256 matrix, 176 slices, 1mm thickness, 1x1x1mm | 5.3 | Sagittal | Linux  centos6_x86_64 |
| Melbourne (YODA) | 3T GE Signa | 3D BRAVO:  TR=7.9s, TR=3s, Flip angle=13 degrees, 140 slices  with 1mm thickness, FOV: 256x256 | 5.3 | Sagittal | Linux Debian x86 86 |
| DEP-ARREST-CLIN - MOODS | 3T Philips Achieva | 3D T1-weighted  image: TR=7, TE=3.5, FOV=352x352x180, Flip angle=8 degrees, number of slices : 180 slice, Slice gap 1 mm, , voxel size: 0.8x0.8x1 | 6.0 | Transverse | CentOS Linux 7 |
| PAFIP2 | 3T Philips Achieva | TR=8.10ms, TE=3.70ms, flip angle=8 degrees, 160 slices, slice thickness 1mm, slice gap 1mm, FOV:256x256, voxel size=0.9375 | 6.0 | Sagittal | Ubuntu 16.04 LTS |
| PAFIP1 | 1.5T GE | Spoiled grass (SPGR) sequence TE=5 ms, TR=24 ms, FOV=26×19.5 cm, slice thickness=1.5mm and a matrix of 256×192. | 5.0 | Coronal | Ubuntu 11.04 |
| Sydney Bipolar Kids and Siblings | 3T Philips Achieva | Fast-field echo scan: TR=5.5ms, TE=2.5ms, flip angle=8 degrees,  180 slices FOV=256x256x180mm3, voxel size 1x1x1mm | 5.3 | Sagittal | CentOSLinux 6.6 |
| Fondazione Santa Lucia | 3T Siemens Magnetom Allegra | 3D MDEFT, TE/TR=2.4/7.92ms, flip angle=15°  matrix 256x256 | 5.3 | Sagittal | Linux |
| Stanford TIGER | 3T GE MR750 | TR/TE/TI=8.2/3.2/600 ms; flip angle=12°, 156 slices, 1mm thickness, voxel size: 1x1x1mm | 6.0 | Axial | CentOS Linux 7 |

Table S3. Definitions used for current suicidal ideation and history of suicide attempt

| **Site** | **Definition**  **current suicidal ideation** | **Definition**  **history of suicide attempt** |
| --- | --- | --- |
| EPISCA | Childhood Depression Inventory item on suicidal ideation scored 1 (“I think about ending my life but would not do it”) or 2 (“I want to end my life”) | NA |
| University of Texas - Austin - Bipolar Seed Program | Structured Clinical Interview DSM-5 (SCID-RV/NP for DSM-5) Suicidality module: answer “yes” to question on ideation in the past week (“Have you in the past week wished you were dead or wished you could go to sleep and not wake up?”) | Structured Clinical Interview DSM-5 (SCID-RV/NP for DSM-5) Suicidality module: answer “yes” to question “Have you ever tried to kill yourself?” |
| Boystown | Mood and Feelings Questionnaire (self-report) item 19 (“I thought about killing myself”) coded as 1 “Sometimes” or 2 “True” OR Childhood Depression Inventory-2 item on suicidal ideation scored 1 (“I think about ending my life but would never do it”) or 2 (“I want to kill myself”) | NA |
| FOR2107-Marburg | Beck Depression Inventory item on suicidal ideation scored 1 (“I think about ending my life but would not do it”), 2 (“I would like to end my life”), 3 (“I would kill myself if I had the opportunity”) OR Hamilton Depression Rating Scale (HDRS) suicidal ideation item scored 2 “Wishes to be dead or thinks about possible death” or higher | NA |
| FOR2107-Münster | Beck Depression Inventory item on suicidal ideation scored 1 (“I think about ending my life but would not do it”), 2 (“I would like to end my life”), 3 (“I would kill myself if I had the opportunity”) OR Hamilton Depression Rating Scale (HDRS-21) suicidal ideation item scored 2 “Wishes to be dead or thinks about possible death” or higher | NA |
| Houston BD | Hamilton Depression Rating Scale suicidal ideation item scored 2 (“Wishes to be dead or thinks about possible death”) or higher | Clinical interview: number of lifetime suicide attempts > 0 |
| MDD_Cohort | Hamilton Depression Rating Scale suicidal ideation item scored 2 (“Wishes to be dead or thinks about  possible death”) or higher | NA |
| University of Minnesota | Beck Depression Inventory-2 item on suicidal ideation scored 1 (“I have thoughts of killing myself, but I would  not carry them out”), 2 (“I would like to kill myself”), 3 (“I would kill myself if I had the chance”) OR Childrens Depression Rating Scale 3 or higher | Kiddie Schedule for Affective Disorders and Schzophrenia for School Aged Children- Lifetime  version (K-SADS-PL_2009) item on suicide attempt answer 1 |
| Muenster Neuroimaging Cohort | Beck Depression Inventory-2 item on suicidal ideation scored 1 (“I thave thoughts of killing myself, but I would not carry them out”), 2 (“I would like to kill myself”), 3 (“I would kill myself if I had the chance”) OR Montgomery-Asberg Depression Rating Scale (MADRS) item on suicidal ideation scored between 2 (“Weary of life, only fleeting suicidal thoughts”) and 6 (“Explicit plans for  suicide when there is an opportunity. Active preparations for suicide”) | NA |
| Yale School of Medicine | Columbia Suicide Severity Rating Scale (C-SSRS) positive answer on one or more of the following items (in the past month): current non-specific active suicidal ideation, current active suicidal ideation with any method (not plan) without intent to act, current active suicidal ideation with some intent without specific plan, current active suicidal ideation with specific plan and intent | Columbia Suicide Severity Rating Scale (C-SSRS) question on lifetime actual suicide attempt answered “yes” |
| MR-IMPACT | Columbia Suicide Severity Rating Scale (C-SSRS) positive answer on one or more of the following items (in the past month): current non-specific active suicidal ideation, current active suicidal ideation with any method (not plan) without intent to act, current active suicidal ideation with some intent without specific plan, current active suicidal ideation with specific plan and intent | Columbia Suicide Severity Rating Scale (C-SSRS) question on lifetime actual suicide attempt answered “yes” |
| SOCAT | Beck Depression Inventory item on suicidal ideation scored 1 (“I think about ending my life but would not do it”), 2 (“I would like to end my life”), 3 (“I would kill myself if I had the opportunity”) OR Hamilton Depression Rating Scale (HDRS-21) suicidal ideation item scored 2 “Wishes to be dead or thinks about possible death” or higher | NA |
| Stanford TAD | Columbia Suicide Severity Rating  Scale (C-SSRS) positive answer on one or more of the following items (in the past week): current non-specific active suicidal ideation, current active suicidal ideation with any method (not plan) without intent to act, current active suicidal ideation with some intent without specific plan, current active suicidal ideation with specific plan and intent | Columbia Suicide Severity Rating  Scale (C-SSRS) question on lifetime actual attempt answered “yes” |
| Sydney Brain and Mind Centre | Hamilton Depression Rating Scale suicidal ideation item scored 2 “Wishes to be dead or thinks about  possible death” or higher | Clinical interview: lifetime suicide attempt coded “yes” |
| UCSF | Columbia Suicide Severity Rating Scale (C-SSRS) positive answer on one or more of the following items (in the past week): current non-specific active suicidal ideation, current active suicidal ideation with any method (not plan) without intent to act, current active suicidal ideation with some intent without specific plan, current active suicidal ideation with specific plan and intent | Columbia Suicide Severity Rating Scale (C-SSRS) question on lifetime actual attempt answered “yes” |
| UWashington/  Harvard | Childhood Depression Inventory item on suicidal ideation scored 1 (“I think about ending my life but would not do it”) or 2 (“I would like to end my life”) | Self-injurious Thoughts and Behaviors Interview (SITBI) measure on lifetime suicide attempt answered “yes” |
| Melbourne (YODA) | Columbia Suicide Severity Rating Scale (C-SSRS) positive answer on one or more of the following items (in the past month): current non-specific active suicidal ideation, current active suicidal ideation with any method (not plan) without intent to act, current active suicidal ideation with some intent without specific plan, current active suicidal ideation with specific plan and intent | Columbia Suicide Severity Rating Scale (C-SSRS) question on lifetime actual attempt answered “yes” |
| PAFIP1&2 | NA | Lifetime history of suicide attempt  determined using an interview |
| Sydney Bipolar Kids and Siblings | NA | Diagnostic Interview for Genetic Studies (DIGS) question on lifetime suicide attempt answered “yes” OR Kiddie Schedule for Affective  Disorders and Schzophrenia (KSADS) item on lifetime suicide attempt coded as “yes” |
| Fondazione Santa Lucia SZ sample | MINI suicide module item C3 (in the past month, have you thought about suicided?) answered as “yes” | Lifetime history of suicide attempt determined using an interview |
| DEP-ARREST-CLIN - MOODS | Hamilton Depression Rating Scale suicidal ideation item scored 2 (“Wishes to be dead or thinks about possible death”) or higher | Recent OR past suicide attempt determined in an interview |

Table S4. Association between lifetime intensity of ideation (scored 0-5) in the Columbia Suicide Severity Rating Scale and brain morphology (healthy controls were excluded). Beta: standardized beta, p: p-value, FDR-p: FDR corrected p-value.

| **Region** | **Beta** | **T-value** | **P** | **FDR-p** | **N** |
| --- | --- | --- | --- | --- | --- |
| **Subcortical volume** |  |  |  |  |  |
| Ventricle | 0.016 | 0.239 | 0.811 | 0.966 | 233 |
| Thalamus | -0.021 | -0.621 | 0.535 | 0.966 | 414 |
| Caudate | 0.017 | 0.392 | 0.695 | 0.966 | 414 |
| Putamen | 0.026 | 0.591 | 0.555 | 0.966 | 398 |
| Pallidum | -0.016 | -0.367 | 0.714 | 0.966 | 407 |
| Hippocampus | 0.082 | 2.100 | 0.036 | 0.936 | 419 |
| Amygdala | 0.049 | 1.291 | 0.197 | 0.966 | 416 |
| Accumbens | -0.003 | -0.058 | 0.954 | 0.966 | 414 |
| **Cortical thickness** |  |  |  |  |  |
| Banks superior temporal sulcus | 0.016 | 0.063 | 0.950 | 0.966 | 387 |
| Caudal anterior cingulate cortex | -0.021 | -0.471 | 0.638 | 0.966 | 420 |
| Caudal middle frontal gyrus | 0.017 | 0.702 | 0.483 | 0.966 | 418 |
| Cuneus | 0.026 | -0.745 | 0.457 | 0.966 | 415 |
| Entorhinal cortex | -0.016 | -0.207 | 0.836 | 0.966 | 389 |
| Fusiform gyrus | 0.082 | -0.724 | 0.470 | 0.966 | 419 |
| Inferior parietal cortex | 0.049 | 1.152 | 0.250 | 0.966 | 420 |
| Inferior temporal gyrus | -0.003 | -0.149 | 0.882 | 0.966 | 421 |
| Isthmus cingulate cortex | 0.016 | 1.495 | 0.136 | 0.966 | 421 |
| Lateral occipital cortex | -0.021 | -0.209 | 0.834 | 0.966 | 421 |
| Lateral orbitofrontal cortex | 0.017 | 0.301 | 0.764 | 0.966 | 413 |
| Lingual gyrus | 0.026 | -0.163 | 0.871 | 0.966 | 421 |
| Medial orbitofrontal gyrus | -0.016 | 0.663 | 0.508 | 0.966 | 416 |
| Middle temporal gyrus | 0.082 | -0.605 | 0.545 | 0.966 | 400 |
| Parahippocampal gyrus | 0.049 | 0.671 | 0.503 | 0.966 | 418 |
| Paracentral lobule | -0.003 | 1.060 | 0.290 | 0.966 | 421 |
| Pars opercularis | 0.016 | 0.109 | 0.913 | 0.966 | 420 |
| Pars orbitalis | -0.021 | 0.871 | 0.384 | 0.966 | 419 |
| Pars triangularis | 0.017 | 0.944 | 0.346 | 0.966 | 421 |
| Pericalcarine cortex | 0.026 | -1.434 | 0.152 | 0.966 | 421 |
| Postcentral gyrus | -0.016 | 0.362 | 0.718 | 0.966 | 419 |
| Posterior cingulate cortex | 0.082 | 0.398 | 0.691 | 0.966 | 420 |
| Precentral gyrus | 0.049 | 0.517 | 0.605 | 0.966 | 418 |
| Precuneus | -0.003 | 0.760 | 0.448 | 0.966 | 422 |
| Rostral anterior cingulate cortex | 0.016 | 0.631 | 0.528 | 0.966 | 420 |
| Rostral middle frontal gyrus | -0.021 | 0.122 | 0.903 | 0.966 | 418 |
| Superior frontal gyrus | 0.017 | 2.395 | 0.017 | 0.663 | 420 |
| Superior parietal gyrus | 0.026 | 0.749 | 0.454 | 0.966 | 421 |
| Superior temporal gyrus | -0.016 | 0.103 | 0.918 | 0.966 | 402 |
| Supramarginal gyrus | 0.082 | 1.168 | 0.243 | 0.966 | 420 |
| Frontal pole | 0.049 | 0.259 | 0.796 | 0.966 | 420 |
| Temporal pole | -0.003 | -0.521 | 0.602 | 0.966 | 422 |
| Transverse temporal gyrus | 0.016 | -0.356 | 0.722 | 0.966 | 422 |
| Insula | -0.021 | 0.807 | 0.420 | 0.966 | 421 |
| Mean Thickness | 0.017 | 0.347 | 0.728 | 0.966 | 421 |
| **Cortical surface area** |  |  |  |  |  |
| Banks superior temporal sulcus | -0.005 | -0.114 | 0.909 | 0.966 | 387 |
| Caudal anterior cingulate cortex | -0.004 | -0.105 | 0.917 | 0.966 | 419 |
| Caudal middle frontal gyrus | -0.012 | -0.275 | 0.784 | 0.966 | 420 |
| Cuneus | 0.065 | 1.567 | 0.118 | 0.966 | 411 |
| Entorhinal cortex | 0.059 | 1.220 | 0.223 | 0.966 | 388 |
| Fusiform gyrus | 0.003 | 0.079 | 0.937 | 0.966 | 419 |
| Inferior parietal cortex | -0.010 | -0.270 | 0.787 | 0.966 | 421 |
| Inferior temporal gyrus | -0.005 | -0.120 | 0.904 | 0.966 | 419 |
| Isthmus cingulate cortex | 0.044 | 1.094 | 0.275 | 0.966 | 417 |
| Lateral occipital cortex | 0.018 | 0.474 | 0.636 | 0.966 | 417 |
| Lateral orbitofrontal cortex | -0.037 | -0.976 | 0.330 | 0.966 | 416 |
| Lingual gyrus | 0.043 | 1.036 | 0.301 | 0.966 | 419 |
| Medial orbitofrontal gyrus | -0.002 | -0.057 | 0.954 | 0.966 | 415 |
| Middle temporal gyrus | 0.027 | 0.710 | 0.478 | 0.966 | 402 |
| Parahippocampal gyrus | 0.061 | 1.399 | 0.163 | 0.966 | 415 |
| Paracentral lobule | 0.022 | 0.519 | 0.604 | 0.966 | 419 |
| Pars opercularis | 0.031 | 0.687 | 0.493 | 0.966 | 418 |
| Pars orbitalis | -0.009 | -0.232 | 0.817 | 0.966 | 419 |
| Pars triangularis | 0.049 | 1.133 | 0.258 | 0.966 | 421 |
| Pericalcarine cortex | 0.065 | 1.467 | 0.143 | 0.966 | 419 |
| Postcentral gyrus | 0.056 | 1.414 | 0.158 | 0.966 | 419 |
| Posterior cingulate cortex | -0.018 | -0.454 | 0.650 | 0.966 | 421 |
| Precentral gyrus | 0.021 | 0.556 | 0.578 | 0.966 | 420 |
| Precuneus | -0.009 | -0.267 | 0.790 | 0.966 | 420 |
| Rostral anterior cingulate cortex | -0.065 | -1.726 | 0.085 | 0.966 | 418 |
| Rostral middle frontal gyrus | -0.050 | -1.409 | 0.160 | 0.966 | 419 |
| Superior frontal gyrus | -0.042 | -1.213 | 0.226 | 0.966 | 421 |
| Superior parietal gyrus | 0.014 | 0.363 | 0.717 | 0.966 | 423 |
| Superior temporal gyrus | -0.001 | -0.036 | 0.971 | 0.971 | 401 |
| Supramarginal gyrus | 0.009 | 0.241 | 0.810 | 0.966 | 421 |
| Frontal pole | -0.111 | -2.515 | 0.012 | 0.663 | 420 |
| Temporal pole | -0.056 | -1.303 | 0.193 | 0.966 | 423 |
| Transverse temporal gyrus | 0.025 | 0.584 | 0.560 | 0.966 | 422 |
| Insula | -0.033 | -0.917 | 0.359 | 0.966 | 421 |
| Full surface area | 0.006 | 0.222 | 0.824 | 0.966 | 422 |

Table S5. Association between recent intensity of ideation (in the past week or month, scored 0-5) in the Columbia Suicide Severity Rating Scale and brain morphology (healthy controls were excluded). Beta: standardized beta, p: p-value, FDR-p: FDR corrected p-value.

| **Region** | **Beta** | **T-value** | **P** | **FDR-p** | **N** |
| --- | --- | --- | --- | --- | --- |
| **Subcortical volume** |  |  |  |  |  |
| Ventricle | -0.126 | -2.143 | 0.033 | 0.683 | 285 |
| Thalamus | -0.036 | -1.171 | 0.242 | 0.796 | 486 |
| Caudate | 0.041 | 1.064 | 0.288 | 0.826 | 486 |
| Putamen | 0.019 | 0.467 | 0.641 | 0.984 | 456 |
| Pallidum | 0.012 | 0.287 | 0.774 | 0.984 | 464 |
| Hippocampus | -0.003 | -0.076 | 0.939 | 0.984 | 486 |
| Amygdala | 0.012 | 0.342 | 0.732 | 0.984 | 489 |
| Accumbens | 0.049 | 1.206 | 0.228 | 0.796 | 487 |
| **Cortical thickness** |  |  |  |  |  |
| Banks superior temporal sulcus | -0.041 | -0.890 | 0.374 | 0.826 | 451 |
| Caudal anterior cingulate cortex | -0.061 | -1.358 | 0.175 | 0.758 | 492 |
| Caudal middle frontal gyrus | -0.067 | -1.493 | 0.136 | 0.683 | 490 |
| Cuneus | 0.051 | 1.141 | 0.255 | 0.796 | 485 |
| Entorhinal cortex | -0.008 | -0.176 | 0.861 | 0.984 | 462 |
| Fusiform gyrus | 0.039 | 0.879 | 0.380 | 0.826 | 492 |
| Inferior parietal cortex | -0.002 | -0.044 | 0.965 | 0.984 | 491 |
| Inferior temporal gyrus | -0.013 | -0.288 | 0.773 | 0.984 | 492 |
| Isthmus cingulate cortex | -0.003 | -0.078 | 0.937 | 0.984 | 494 |
| Lateral occipital cortex | 0.001 | 0.021 | 0.983 | 0.984 | 494 |
| Lateral orbitofrontal cortex | -0.005 | -0.112 | 0.911 | 0.984 | 485 |
| Lingual gyrus | -0.004 | -0.090 | 0.929 | 0.984 | 494 |
| Medial orbitofrontal gyrus | -0.014 | -0.329 | 0.742 | 0.984 | 487 |
| Middle temporal gyrus | -0.071 | -1.538 | 0.125 | 0.683 | 464 |
| Parahippocampal gyrus | 0.004 | 0.085 | 0.932 | 0.984 | 488 |
| Paracentral lobule | -0.008 | -0.182 | 0.856 | 0.984 | 494 |
| Pars opercularis | -0.027 | -0.611 | 0.542 | 0.983 | 492 |
| Pars orbitalis | -0.074 | -1.658 | 0.098 | 0.683 | 491 |
| Pars triangularis | -0.082 | -1.868 | 0.062 | 0.683 | 493 |
| Pericalcarine cortex | -0.026 | -0.568 | 0.570 | 0.984 | 490 |
| Postcentral gyrus | -0.081 | -1.804 | 0.072 | 0.683 | 491 |
| Posterior cingulate cortex | -0.036 | -0.843 | 0.400 | 0.826 | 493 |
| Precentral gyrus | -0.041 | -0.896 | 0.371 | 0.826 | 489 |
| Precuneus | -0.004 | -0.088 | 0.930 | 0.984 | 495 |
| Rostral anterior cingulate cortex | -0.073 | -1.626 | 0.105 | 0.683 | 493 |
| Rostral middle frontal gyrus | -0.095 | -2.178 | 0.030 | 0.683 | 490 |
| Superior frontal gyrus | -0.001 | -0.020 | 0.984 | 0.984 | 491 |
| Superior parietal gyrus | -0.020 | -0.447 | 0.655 | 0.984 | 493 |
| Superior temporal gyrus | -0.067 | -1.479 | 0.140 | 0.683 | 471 |
| Supramarginal gyrus | -0.072 | -1.623 | 0.105 | 0.683 | 492 |
| Frontal pole | 0.037 | 0.819 | 0.413 | 0.826 | 493 |
| Temporal pole | 0.007 | 0.164 | 0.870 | 0.984 | 495 |
| Transverse temporal gyrus | -0.009 | -0.201 | 0.840 | 0.984 | 495 |
| Insula | 0.020 | 0.454 | 0.650 | 0.984 | 493 |
| Mean Thickness | -0.042 | -0.965 | 0.335 | 0.826 | 494 |
| **Cortical surface area** |  |  |  |  |  |
| Banks superior temporal sulcus | 0.034 | 0.837 | 0.403 | 0.826 | 451 |
| Caudal anterior cingulate cortex | 0.039 | 1.022 | 0.307 | 0.826 | 492 |
| Caudal middle frontal gyrus | -0.007 | -0.166 | 0.868 | 0.984 | 491 |
| Cuneus | 0.024 | 0.632 | 0.528 | 0.981 | 481 |
| Entorhinal cortex | 0.039 | 0.902 | 0.367 | 0.826 | 460 |
| Fusiform gyrus | 0.037 | 1.163 | 0.245 | 0.796 | 492 |
| Inferior parietal cortex | -0.002 | -0.054 | 0.957 | 0.984 | 493 |
| Inferior temporal gyrus | 0.065 | 1.925 | 0.055 | 0.683 | 490 |
| Isthmus cingulate cortex | 0.047 | 1.314 | 0.189 | 0.776 | 490 |
| Lateral occipital cortex | 0.002 | 0.054 | 0.957 | 0.984 | 490 |
| Lateral orbitofrontal cortex | -0.009 | -0.257 | 0.797 | 0.984 | 488 |
| Lingual gyrus | 0.033 | 0.890 | 0.374 | 0.826 | 492 |
| Medial orbitofrontal gyrus | -0.007 | -0.218 | 0.827 | 0.984 | 486 |
| Middle temporal gyrus | 0.035 | 1.002 | 0.317 | 0.826 | 467 |
| Parahippocampal gyrus | 0.060 | 1.535 | 0.125 | 0.683 | 485 |
| Paracentral lobule | 0.010 | 0.256 | 0.798 | 0.984 | 492 |
| Pars opercularis | 0.074 | 1.874 | 0.062 | 0.683 | 490 |
| Pars orbitalis | 0.008 | 0.227 | 0.820 | 0.984 | 492 |
| Pars triangularis | 0.084 | 2.128 | 0.034 | 0.683 | 493 |
| Pericalcarine cortex | 0.043 | 1.057 | 0.291 | 0.826 | 489 |
| Postcentral gyrus | 0.052 | 1.477 | 0.140 | 0.683 | 492 |
| Posterior cingulate cortex | 0.047 | 1.266 | 0.206 | 0.796 | 494 |
| Precentral gyrus | -0.002 | -0.046 | 0.963 | 0.984 | 492 |
| Precuneus | -0.012 | -0.373 | 0.710 | 0.984 | 493 |
| Rostral anterior cingulate cortex | -0.007 | -0.193 | 0.847 | 0.984 | 491 |
| Rostral middle frontal gyrus | -0.012 | -0.353 | 0.724 | 0.984 | 491 |
| Superior frontal gyrus | 0.007 | 0.218 | 0.828 | 0.984 | 492 |
| Superior parietal gyrus | 0.005 | 0.150 | 0.881 | 0.984 | 496 |
| Superior temporal gyrus | 0.023 | 0.681 | 0.497 | 0.946 | 470 |
| Supramarginal gyrus | 0.048 | 1.389 | 0.166 | 0.758 | 493 |
| Frontal pole | -0.032 | -0.794 | 0.428 | 0.835 | 493 |
| Temporal pole | -0.034 | -0.868 | 0.386 | 0.826 | 496 |
| Transverse temporal gyrus | 0.070 | 1.799 | 0.073 | 0.683 | 494 |
| Insula | -0.019 | -0.564 | 0.573 | 0.984 | 492 |
| Full surface area | 0.030 | 1.187 | 0.236 | 0.796 | 495 |

Table S6. Differences in brain morphology between young people with a lifetime history of an actual suicide attempt and those without a history of an actual suicide attempt determined using the Columbia Suicide Severity Rating Scale (healthy controls were excluded).

D: Cohen’s d effect size, SE: standard error; p: p-value, FDR-p: FDR corrected p-value, CI: confidence interval, HC: healthy controls, CC: clinical controls.

| **Region** | **D** | **SE** | **P** | **FDR-p** | **N attempt** | **N no attempt** |
| --- | --- | --- | --- | --- | --- | --- |
| **Subcortical volume** |  |  |  |  |  |  |
| Ventricle | -0.025 | 0.143 | 0.842 | 0.911 | 98 | 219 |
| Thalamus | -0.118 | 0.113 | 0.215 | 0.689 | 157 | 392 |
| Caudate | 0.058 | 0.112 | 0.538 | 0.819 | 160 | 386 |
| Putamen | -0.027 | 0.116 | 0.779 | 0.911 | 149 | 367 |
| Pallidum | -0.107 | 0.114 | 0.267 | 0.689 | 155 | 370 |
| Hippocampus | -0.033 | 0.111 | 0.725 | 0.898 | 162 | 387 |
| Amygdala | -0.057 | 0.112 | 0.546 | 0.819 | 159 | 393 |
| Accumbens | -0.018 | 0.112 | 0.853 | 0.911 | 160 | 386 |
| **Cortical thickness** |  |  |  |  |  |  |
| Banks superior temporal sulcus | -0.087 | 0.116 | 0.376 | 0.733 | 148 | 357 |
| Caudal anterior cingulate cortex | 0.091 | 0.111 | 0.333 | 0.733 | 162 | 394 |
| Caudal middle frontal gyrus | 0.005 | 0.111 | 0.961 | 0.961 | 163 | 391 |
| Cuneus | -0.023 | 0.111 | 0.804 | 0.911 | 161 | 388 |
| Entorhinal cortex | -0.041 | 0.117 | 0.680 | 0.873 | 147 | 364 |
| Fusiform gyrus | -0.192 | 0.111 | 0.041 | 0.457 | 162 | 393 |
| Inferior parietal cortex | 0.084 | 0.111 | 0.372 | 0.733 | 162 | 391 |
| Inferior temporal gyrus | -0.108 | 0.111 | 0.248 | 0.689 | 163 | 393 |
| Isthmus cingulate cortex | 0.122 | 0.111 | 0.194 | 0.689 | 163 | 395 |
| Lateral occipital cortex | 0.038 | 0.111 | 0.685 | 0.873 | 163 | 393 |
| Lateral orbitofrontal cortex | -0.137 | 0.112 | 0.148 | 0.689 | 160 | 388 |
| Lingual gyrus | -0.102 | 0.111 | 0.274 | 0.689 | 164 | 394 |
| Medial orbitofrontal gyrus | -0.031 | 0.112 | 0.745 | 0.908 | 160 | 390 |
| Middle temporal gyrus | -0.115 | 0.115 | 0.238 | 0.689 | 152 | 368 |
| Parahippocampal gyrus | -0.037 | 0.111 | 0.693 | 0.873 | 161 | 391 |
| Paracentral lobule | 0.076 | 0.110 | 0.416 | 0.753 | 164 | 393 |
| Pars opercularis | -0.089 | 0.111 | 0.343 | 0.733 | 163 | 393 |
| Pars orbitalis | 0.072 | 0.111 | 0.440 | 0.753 | 163 | 392 |
| Pars triangularis | 0.020 | 0.110 | 0.831 | 0.911 | 164 | 393 |
| Pericalcarine cortex | -0.144 | 0.111 | 0.125 | 0.689 | 162 | 392 |
| Postcentral gyrus | -0.081 | 0.111 | 0.389 | 0.740 | 163 | 392 |
| Posterior cingulate cortex | 0.103 | 0.111 | 0.272 | 0.689 | 164 | 393 |
| Precentral gyrus | -0.049 | 0.111 | 0.598 | 0.873 | 163 | 390 |
| Precuneus | -0.084 | 0.110 | 0.371 | 0.733 | 164 | 395 |
| Rostral anterior cingulate cortex | 0.010 | 0.111 | 0.911 | 0.940 | 161 | 396 |
| Rostral middle frontal gyrus | -0.028 | 0.110 | 0.764 | 0.911 | 164 | 390 |
| Superior frontal gyrus | 0.105 | 0.111 | 0.264 | 0.689 | 163 | 392 |
| Superior parietal gyrus | 0.010 | 0.111 | 0.916 | 0.940 | 163 | 394 |
| Superior temporal gyrus | -0.061 | 0.114 | 0.530 | 0.819 | 153 | 374 |
| Supramarginal gyrus | -0.023 | 0.111 | 0.806 | 0.911 | 161 | 393 |
| Frontal pole | 0.038 | 0.111 | 0.685 | 0.873 | 162 | 395 |
| Temporal pole | -0.117 | 0.111 | 0.210 | 0.689 | 164 | 395 |
| Transverse temporal gyrus | -0.071 | 0.110 | 0.444 | 0.753 | 164 | 395 |
| Insula | -0.037 | 0.111 | 0.694 | 0.873 | 163 | 394 |
| Mean Thickness | -0.038 | 0.110 | 0.682 | 0.873 | 164 | 394 |
| **Cortical surface area** |  |  |  |  |  |  |
| Banks superior temporal sulcus | -0.163 | 0.116 | 0.097 | 0.689 | 149 | 355 |
| Caudal anterior cingulate cortex | -0.260 | 0.112 | 0.006 | 0.156 | 162 | 394 |
| Caudal middle frontal gyrus | -0.059 | 0.111 | 0.531 | 0.819 | 163 | 392 |
| Cuneus | 0.020 | 0.112 | 0.836 | 0.911 | 159 | 386 |
| Entorhinal cortex | -0.043 | 0.117 | 0.663 | 0.873 | 146 | 363 |
| Fusiform gyrus | -0.131 | 0.111 | 0.164 | 0.689 | 162 | 394 |
| Inferior parietal cortex | -0.282 | 0.112 | 0.003 | 0.117 | 162 | 393 |
| Inferior temporal gyrus | -0.231 | 0.111 | 0.014 | 0.273 | 162 | 392 |
| Isthmus cingulate cortex | -0.014 | 0.111 | 0.883 | 0.931 | 163 | 391 |
| Lateral occipital cortex | -0.062 | 0.111 | 0.508 | 0.819 | 162 | 391 |
| Lateral orbitofrontal cortex | -0.125 | 0.112 | 0.183 | 0.689 | 161 | 390 |
| Lingual gyrus | 0.037 | 0.111 | 0.689 | 0.873 | 162 | 394 |
| Medial orbitofrontal gyrus | -0.142 | 0.112 | 0.134 | 0.689 | 160 | 389 |
| Middle temporal gyrus | -0.138 | 0.114 | 0.155 | 0.689 | 153 | 369 |
| Parahippocampal gyrus | 0.103 | 0.111 | 0.272 | 0.689 | 162 | 387 |
| Paracentral lobule | -0.125 | 0.111 | 0.181 | 0.689 | 164 | 392 |
| Pars opercularis | 0.024 | 0.111 | 0.797 | 0.911 | 163 | 390 |
| Pars orbitalis | -0.096 | 0.111 | 0.308 | 0.728 | 163 | 393 |
| Pars triangularis | 0.008 | 0.111 | 0.935 | 0.947 | 163 | 393 |
| Pericalcarine cortex | 0.074 | 0.112 | 0.431 | 0.753 | 160 | 393 |
| Postcentral gyrus | -0.120 | 0.111 | 0.198 | 0.689 | 164 | 392 |
| Posterior cingulate cortex | -0.176 | 0.111 | 0.060 | 0.585 | 164 | 394 |
| Precentral gyrus | -0.107 | 0.111 | 0.255 | 0.689 | 162 | 394 |
| Precuneus | -0.083 | 0.110 | 0.375 | 0.733 | 164 | 393 |
| Rostral anterior cingulate cortex | -0.223 | 0.112 | 0.018 | 0.281 | 161 | 394 |
| Rostral middle frontal gyrus | -0.203 | 0.111 | 0.030 | 0.390 | 164 | 391 |
| Superior frontal gyrus | -0.139 | 0.111 | 0.139 | 0.689 | 163 | 393 |
| Superior parietal gyrus | -0.096 | 0.110 | 0.303 | 0.728 | 164 | 396 |
| Superior temporal gyrus | -0.061 | 0.114 | 0.528 | 0.819 | 154 | 370 |
| Supramarginal gyrus | -0.106 | 0.111 | 0.260 | 0.689 | 163 | 392 |
| Frontal pole | -0.334 | 0.112 | 0.000 | 0.000 | 163 | 394 |
| Temporal pole | -0.074 | 0.110 | 0.429 | 0.753 | 164 | 396 |
| Transverse temporal gyrus | -0.038 | 0.110 | 0.685 | 0.873 | 164 | 394 |
| Insula | -0.092 | 0.111 | 0.326 | 0.733 | 163 | 393 |
| Full surface area | -0.150 | 0.111 | 0.108 | 0.689 | 164 | 395 |

Table S7. Differences in brain morphology between young people with a lifetime history of an actual suicide and those without a history of an actual suicide attempt but with lifetime suicidal ideation determined with the Columbia Suicide Severity Rating Scale (healthy controls were excluded). D: Cohen’s d effect size, SE: standard error; p: p-value, FDR-p: FDR corrected p-value, CI: confidence interval.

| **Region** | **D** | **SE** | **P** | **FDR-p** | **N attempt** | **N ideation** |
| --- | --- | --- | --- | --- | --- | --- |
| **Subcortical volume** |  |  |  |  |  |  |
| Ventricle | -0.038 | 0.138 | 0.786 | 0.924 | 98 | 112 |
| Thalamus | -0.120 | 0.108 | 0.273 | 0.755 | 157 | 190 |
| Caudate | 0.028 | 0.108 | 0.798 | 0.924 | 160 | 188 |
| Putamen | -0.004 | 0.110 | 0.972 | 0.978 | 149 | 184 |
| Pallidum | -0.063 | 0.109 | 0.568 | 0.851 | 155 | 186 |
| Hippocampus | -0.123 | 0.107 | 0.256 | 0.755 | 162 | 189 |
| Amygdala | -0.131 | 0.108 | 0.228 | 0.755 | 159 | 190 |
| Accumbens | -0.008 | 0.108 | 0.942 | 0.978 | 160 | 188 |
| **Cortical thickness** |  |  |  |  |  |  |
| Banks superior temporal sulcus | -0.114 | 0.111 | 0.308 | 0.755 | 148 | 179 |
| Caudal anterior cingulate cortex | 0.070 | 0.107 | 0.518 | 0.832 | 162 | 191 |
| Caudal middle frontal gyrus | -0.068 | 0.107 | 0.531 | 0.832 | 163 | 187 |
| Cuneus | 0.014 | 0.108 | 0.895 | 0.978 | 161 | 186 |
| Entorhinal cortex | -0.060 | 0.111 | 0.593 | 0.873 | 147 | 183 |
| Fusiform gyrus | -0.224 | 0.107 | 0.038 | 0.426 | 162 | 190 |
| Inferior parietal cortex | 0.107 | 0.107 | 0.320 | 0.755 | 162 | 189 |
| Inferior temporal gyrus | -0.160 | 0.107 | 0.136 | 0.665 | 163 | 191 |
| Isthmus cingulate cortex | 0.078 | 0.107 | 0.466 | 0.832 | 163 | 191 |
| Lateral occipital cortex | 0.082 | 0.107 | 0.449 | 0.832 | 163 | 190 |
| Lateral orbitofrontal cortex | -0.117 | 0.108 | 0.282 | 0.755 | 160 | 187 |
| Lingual gyrus | -0.166 | 0.107 | 0.123 | 0.665 | 164 | 189 |
| Medial orbitofrontal gyrus | -0.052 | 0.107 | 0.632 | 0.913 | 160 | 190 |
| Middle temporal gyrus | -0.073 | 0.109 | 0.510 | 0.832 | 152 | 186 |
| Parahippocampal gyrus | -0.095 | 0.107 | 0.381 | 0.762 | 161 | 188 |
| Paracentral lobule | 0.023 | 0.107 | 0.830 | 0.931 | 164 | 189 |
| Pars opercularis | -0.045 | 0.107 | 0.679 | 0.924 | 163 | 188 |
| Pars orbitalis | 0.128 | 0.107 | 0.236 | 0.755 | 163 | 189 |
| Pars triangularis | 0.043 | 0.107 | 0.686 | 0.924 | 164 | 190 |
| Pericalcarine cortex | -0.128 | 0.107 | 0.235 | 0.755 | 162 | 190 |
| Postcentral gyrus | -0.136 | 0.107 | 0.208 | 0.755 | 163 | 189 |
| Posterior cingulate cortex | 0.065 | 0.107 | 0.544 | 0.832 | 164 | 188 |
| Precentral gyrus | -0.066 | 0.107 | 0.544 | 0.832 | 163 | 187 |
| Precuneus | -0.118 | 0.107 | 0.270 | 0.755 | 164 | 190 |
| Rostral anterior cingulate cortex | -0.030 | 0.107 | 0.780 | 0.924 | 161 | 191 |
| Rostral middle frontal gyrus | -0.008 | 0.107 | 0.944 | 0.978 | 164 | 187 |
| Superior frontal gyrus | 0.036 | 0.107 | 0.740 | 0.924 | 163 | 190 |
| Superior parietal gyrus | 0.011 | 0.107 | 0.920 | 0.978 | 163 | 190 |
| Superior temporal gyrus | -0.040 | 0.109 | 0.716 | 0.924 | 153 | 186 |
| Supramarginal gyrus | -0.027 | 0.107 | 0.806 | 0.924 | 161 | 190 |
| Frontal pole | 0.068 | 0.107 | 0.527 | 0.832 | 162 | 191 |
| Temporal pole | -0.127 | 0.107 | 0.236 | 0.755 | 164 | 190 |
| Transverse temporal gyrus | -0.096 | 0.107 | 0.370 | 0.762 | 164 | 190 |
| Insula | -0.101 | 0.107 | 0.349 | 0.757 | 163 | 191 |
| Mean Thickness | -0.044 | 0.107 | 0.684 | 0.924 | 164 | 190 |
| **Cortical surface area** |  |  |  |  |  |  |
| Banks superior temporal sulcus | -0.193 | 0.111 | 0.087 | 0.522 | 149 | 179 |
| Caudal anterior cingulate cortex | -0.286 | 0.107 | 0.008 | 0.164 | 162 | 191 |
| Caudal middle frontal gyrus | -0.103 | 0.107 | 0.343 | 0.757 | 163 | 189 |
| Cuneus | 0.069 | 0.108 | 0.528 | 0.832 | 159 | 186 |
| Entorhinal cortex | -0.114 | 0.111 | 0.310 | 0.755 | 146 | 184 |
| Fusiform gyrus | -0.140 | 0.107 | 0.199 | 0.755 | 162 | 189 |
| Inferior parietal cortex | -0.333 | 0.108 | 0.002 | 0.085 | 162 | 191 |
| Inferior temporal gyrus | -0.318 | 0.108 | 0.003 | 0.091 | 162 | 190 |
| Isthmus cingulate cortex | 0.033 | 0.107 | 0.762 | 0.924 | 163 | 190 |
| Lateral occipital cortex | -0.084 | 0.107 | 0.438 | 0.832 | 162 | 188 |
| Lateral orbitofrontal cortex | -0.106 | 0.107 | 0.329 | 0.755 | 161 | 188 |
| Lingual gyrus | 0.091 | 0.107 | 0.398 | 0.776 | 162 | 190 |
| Medial orbitofrontal gyrus | -0.212 | 0.108 | 0.052 | 0.448 | 160 | 189 |
| Middle temporal gyrus | -0.231 | 0.109 | 0.037 | 0.426 | 153 | 187 |
| Parahippocampal gyrus | 0.144 | 0.108 | 0.185 | 0.755 | 162 | 186 |
| Paracentral lobule | -0.149 | 0.107 | 0.168 | 0.755 | 164 | 187 |
| Pars opercularis | -0.023 | 0.107 | 0.835 | 0.931 | 163 | 186 |
| Pars orbitalis | -0.115 | 0.107 | 0.290 | 0.755 | 163 | 188 |
| Pars triangularis | 0.003 | 0.107 | 0.978 | 0.978 | 163 | 191 |
| Pericalcarine cortex | 0.113 | 0.107 | 0.296 | 0.755 | 160 | 191 |
| Postcentral gyrus | -0.185 | 0.107 | 0.087 | 0.522 | 164 | 189 |
| Posterior cingulate cortex | -0.186 | 0.107 | 0.085 | 0.522 | 164 | 189 |
| Precentral gyrus | -0.163 | 0.107 | 0.132 | 0.665 | 162 | 190 |
| Precuneus | -0.031 | 0.107 | 0.776 | 0.924 | 164 | 189 |
| Rostral anterior cingulate cortex | -0.187 | 0.107 | 0.086 | 0.522 | 161 | 190 |
| Rostral middle frontal gyrus | -0.219 | 0.107 | 0.044 | 0.426 | 164 | 188 |
| Superior frontal gyrus | -0.075 | 0.107 | 0.490 | 0.832 | 163 | 190 |
| Superior parietal gyrus | -0.037 | 0.106 | 0.730 | 0.924 | 164 | 191 |
| Superior temporal gyrus | -0.035 | 0.109 | 0.748 | 0.924 | 154 | 185 |
| Supramarginal gyrus | -0.095 | 0.107 | 0.378 | 0.762 | 163 | 190 |
| Frontal pole | -0.354 | 0.108 | 0.001 | 0.085 | 163 | 190 |
| Temporal pole | -0.006 | 0.106 | 0.955 | 0.978 | 164 | 191 |
| Transverse temporal gyrus | -0.032 | 0.107 | 0.767 | 0.924 | 164 | 190 |
| Insula | -0.004 | 0.107 | 0.970 | 0.978 | 163 | 191 |
| Full surface area | -0.220 | 0.107 | 0.042 | 0.426 | 164 | 191 |

Table S8. Differences in regional brain morphology between healthy controls and young people with current suicidal ideation.

D: Cohen’s d effect size, SE: standard error; p: p-value, FDR-p: FDR corrected p-value, CI: confidence interval, HC: healthy controls, CC: clinical controls.

| **Region** | **D** | **SE** | **P** | **FDR-p** | **Lower CI** | **Upper CI** | **N ideation** | **N HC** |
| --- | --- | --- | --- | --- | --- | --- | --- | --- |
| **Subcortical volume** | | | | | | | | |
| Ventricle | -0.002 | 0.090 | 0.979 | 0.981 | -0.180 | 0.175 | 202 | 309 |
| Thalamus | -0.044 | 0.064 | 0.492 | 0.898 | -0.170 | 0.082 | 381 | 662 |
| Caudate | 0.061 | 0.064 | 0.340 | 0.827 | -0.064 | 0.187 | 390 | 658 |
| Putamen | -0.020 | 0.065 | 0.763 | 0.981 | -0.147 | 0.108 | 381 | 623 |
| Pallidum | 0.155 | 0.066 | 0.019 | 0.449 | 0.026 | 0.284 | 373 | 605 |
| Hippocampus | -0.116 | 0.064 | 0.069 | 0.791 | -0.241 | 0.009 | 392 | 668 |
| Amygdala | 0.049 | 0.064 | 0.443 | 0.864 | -0.076 | 0.174 | 393 | 662 |
| Accumbens | 0.011 | 0.064 | 0.857 | 0.981 | -0.113 | 0.136 | 392 | 666 |
| **Cortical thickness** | | | | | | | | |
| Banks superior temporal sulcus | 0.032 | 0.066 | 0.627 | 0.926 | -0.098 | 0.162 | 360 | 619 |
| Caudal anterior cingulate cortex | 0.029 | 0.063 | 0.653 | 0.926 | -0.095 | 0.153 | 398 | 671 |
| Caudal middle frontal gyrus | -0.065 | 0.064 | 0.308 | 0.827 | -0.189 | 0.060 | 392 | 674 |
| Cuneus | -0.021 | 0.064 | 0.750 | 0.981 | -0.147 | 0.106 | 380 | 656 |
| Entorhinal cortex | -0.067 | 0.067 | 0.317 | 0.827 | -0.199 | 0.064 | 352 | 600 |
| Fusiform gyrus | -0.014 | 0.063 | 0.830 | 0.981 | -0.138 | 0.110 | 394 | 679 |
| Inferior parietal cortex | -0.146 | 0.064 | 0.023 | 0.449 | -0.272 | -0.021 | 383 | 671 |
| Inferior temporal gyrus | -0.080 | 0.064 | 0.211 | 0.827 | -0.206 | 0.045 | 381 | 675 |
| Isthmus cingulate cortex | -0.106 | 0.063 | 0.096 | 0.827 | -0.230 | 0.018 | 393 | 676 |
| Lateral occipital cortex | -0.133 | 0.064 | 0.038 | 0.593 | -0.257 | -0.008 | 391 | 675 |
| Lateral orbitofrontal cortex | 0.060 | 0.064 | 0.350 | 0.827 | -0.065 | 0.184 | 391 | 677 |
| Lingual gyrus | 0.002 | 0.064 | 0.981 | 0.981 | -0.124 | 0.127 | 385 | 678 |
| Medial orbitofrontal gyrus | -0.038 | 0.064 | 0.548 | 0.926 | -0.163 | 0.086 | 395 | 665 |
| Middle temporal gyrus | -0.077 | 0.066 | 0.240 | 0.827 | -0.206 | 0.051 | 363 | 647 |
| Parahippocampal gyrus | 0.015 | 0.063 | 0.812 | 0.981 | -0.109 | 0.139 | 396 | 682 |
| Paracentral lobule | -0.066 | 0.063 | 0.301 | 0.827 | -0.190 | 0.058 | 393 | 677 |
| Pars opercularis | -0.060 | 0.063 | 0.342 | 0.827 | -0.185 | 0.064 | 394 | 676 |
| Pars orbitalis | -0.031 | 0.064 | 0.627 | 0.926 | -0.156 | 0.094 | 386 | 675 |
| Pars triangularis | -0.086 | 0.063 | 0.173 | 0.827 | -0.211 | 0.038 | 396 | 672 |
| Pericalcarine cortex | 0.019 | 0.064 | 0.771 | 0.981 | -0.107 | 0.144 | 385 | 660 |
| Postcentral gyrus | -0.029 | 0.063 | 0.644 | 0.926 | -0.154 | 0.095 | 396 | 666 |
| Posterior cingulate cortex | -0.091 | 0.063 | 0.154 | 0.827 | -0.215 | 0.034 | 393 | 676 |
| Precentral gyrus | -0.042 | 0.064 | 0.514 | 0.898 | -0.166 | 0.083 | 393 | 671 |
| Precuneus | -0.012 | 0.063 | 0.847 | 0.981 | -0.137 | 0.112 | 392 | 676 |
| Rostral anterior cingulate cortex | -0.081 | 0.064 | 0.206 | 0.827 | -0.205 | 0.044 | 395 | 659 |
| Rostral middle frontal gyrus | -0.055 | 0.064 | 0.389 | 0.827 | -0.181 | 0.070 | 387 | 664 |
| Superior frontal gyrus | -0.060 | 0.064 | 0.347 | 0.827 | -0.185 | 0.065 | 391 | 670 |
| Superior parietal gyrus | -0.030 | 0.063 | 0.634 | 0.926 | -0.154 | 0.094 | 396 | 679 |
| Superior temporal gyrus | -0.056 | 0.067 | 0.403 | 0.827 | -0.186 | 0.075 | 353 | 624 |
| Supramarginal gyrus | -0.059 | 0.065 | 0.365 | 0.827 | -0.185 | 0.068 | 378 | 655 |
| Frontal pole | 0.096 | 0.063 | 0.130 | 0.827 | -0.028 | 0.220 | 396 | 676 |
| Temporal pole | -0.041 | 0.063 | 0.518 | 0.898 | -0.165 | 0.083 | 395 | 673 |
| Transverse temporal gyrus | 0.053 | 0.063 | 0.401 | 0.827 | -0.071 | 0.177 | 396 | 683 |
| Insula | -0.042 | 0.065 | 0.514 | 0.898 | -0.169 | 0.084 | 388 | 627 |
| Mean Thickness | -0.071 | 0.063 | 0.263 | 0.827 | -0.194 | 0.053 | 399 | 686 |
| **Cortical surface area** | | | | | | | | |
| Banks superior temporal sulcus | -0.032 | 0.066 | 0.630 | 0.926 | -0.162 | 0.098 | 360 | 613 |
| Caudal anterior cingulate cortex | -0.005 | 0.063 | 0.934 | 0.981 | -0.130 | 0.119 | 396 | 669 |
| Caudal middle frontal gyrus | -0.115 | 0.063 | 0.071 | 0.791 | -0.239 | 0.009 | 394 | 676 |
| Cuneus | -0.014 | 0.065 | 0.831 | 0.981 | -0.140 | 0.113 | 379 | 652 |
| Entorhinal cortex | 0.168 | 0.067 | 0.013 | 0.449 | 0.036 | 0.299 | 354 | 599 |
| Fusiform gyrus | -0.017 | 0.063 | 0.791 | 0.981 | -0.141 | 0.107 | 392 | 677 |
| Inferior parietal cortex | -0.057 | 0.064 | 0.379 | 0.827 | -0.182 | 0.069 | 383 | 667 |
| Inferior temporal gyrus | -0.005 | 0.064 | 0.938 | 0.981 | -0.131 | 0.121 | 382 | 672 |
| Isthmus cingulate cortex | -0.005 | 0.064 | 0.936 | 0.981 | -0.130 | 0.119 | 391 | 678 |
| Lateral occipital cortex | 0.034 | 0.064 | 0.592 | 0.926 | -0.091 | 0.159 | 389 | 675 |
| Lateral orbitofrontal cortex | -0.003 | 0.064 | 0.960 | 0.981 | -0.128 | 0.121 | 391 | 674 |
| Lingual gyrus | 0.006 | 0.064 | 0.926 | 0.981 | -0.119 | 0.131 | 386 | 679 |
| Medial orbitofrontal gyrus | 0.089 | 0.064 | 0.164 | 0.827 | -0.036 | 0.213 | 394 | 668 |
| Middle temporal gyrus | -0.055 | 0.066 | 0.402 | 0.827 | -0.184 | 0.074 | 362 | 642 |
| Parahippocampal gyrus | -0.155 | 0.064 | 0.015 | 0.449 | -0.279 | -0.030 | 393 | 677 |
| Paracentral lobule | -0.066 | 0.064 | 0.301 | 0.827 | -0.191 | 0.059 | 391 | 674 |
| Pars opercularis | -0.061 | 0.064 | 0.340 | 0.827 | -0.186 | 0.064 | 391 | 671 |
| Pars orbitalis | 0.030 | 0.064 | 0.640 | 0.926 | -0.095 | 0.155 | 387 | 676 |
| Pars triangularis | -0.080 | 0.063 | 0.209 | 0.827 | -0.204 | 0.044 | 396 | 674 |
| Pericalcarine cortex | 0.017 | 0.064 | 0.791 | 0.981 | -0.109 | 0.143 | 385 | 657 |
| Postcentral gyrus | -0.010 | 0.063 | 0.871 | 0.981 | -0.135 | 0.114 | 396 | 664 |
| Posterior cingulate cortex | 0.017 | 0.063 | 0.794 | 0.981 | -0.108 | 0.141 | 394 | 676 |
| Precentral gyrus | -0.003 | 0.063 | 0.959 | 0.981 | -0.128 | 0.121 | 395 | 670 |
| Precuneus | -0.090 | 0.064 | 0.158 | 0.827 | -0.215 | 0.034 | 389 | 678 |
| Rostral anterior cingulate cortex | 0.044 | 0.064 | 0.491 | 0.898 | -0.081 | 0.169 | 391 | 661 |
| Rostral middle frontal gyrus | -0.062 | 0.064 | 0.331 | 0.827 | -0.188 | 0.063 | 388 | 666 |
| Superior frontal gyrus | -0.089 | 0.064 | 0.165 | 0.827 | -0.214 | 0.036 | 391 | 667 |
| Superior parietal gyrus | -0.071 | 0.063 | 0.266 | 0.827 | -0.195 | 0.053 | 396 | 678 |
| Superior temporal gyrus | -0.068 | 0.067 | 0.308 | 0.827 | -0.199 | 0.062 | 354 | 618 |
| Supramarginal gyrus | 0.029 | 0.065 | 0.651 | 0.926 | -0.097 | 0.156 | 378 | 652 |
| Frontal pole | 0.022 | 0.063 | 0.729 | 0.981 | -0.102 | 0.146 | 395 | 677 |
| Temporal pole | 0.081 | 0.063 | 0.201 | 0.827 | -0.043 | 0.206 | 394 | 673 |
| Transverse temporal gyrus | -0.002 | 0.063 | 0.980 | 0.981 | -0.125 | 0.122 | 397 | 684 |
| Insula | -0.019 | 0.065 | 0.767 | 0.981 | -0.146 | 0.108 | 385 | 622 |
| Full surface area | -0.049 | 0.063 | 0.435 | 0.864 | -0.173 | 0.074 | 397 | 685 |

Table S9. Differences in regional brain morphology between clinical controls and young people with current suicidal ideation.

D: Cohen’s d effect size, SE: standard error; p: p-value, FDR-p: FDR corrected p-value, CI: confidence interval, HC: healthy controls, CC: clinical controls

| **Region** | **D** | **SE** | **P** | **FDR-p** | **Lower CI** | **Upper CI** | **N ideation** | **N CC** |
| --- | --- | --- | --- | --- | --- | --- | --- | --- |
| **Subcortical volume** | | | | | | | | |
| Ventricle | -0.068 | 0.089 | 0.449 | 0.761 | -0.243 | 0.107 | 202 | 333 |
| Thalamus | 0.119 | 0.065 | 0.070 | 0.608 | -0.009 | 0.247 | 381 | 618 |
| Caudate | 0.053 | 0.065 | 0.414 | 0.761 | -0.074 | 0.179 | 390 | 626 |
| Putamen | 0.070 | 0.066 | 0.285 | 0.635 | -0.058 | 0.199 | 381 | 601 |
| Pallidum | 0.195 | 0.066 | 0.003 | 0.117 | 0.065 | 0.325 | 373 | 592 |
| Hippocampus | 0.056 | 0.065 | 0.386 | 0.753 | -0.070 | 0.183 | 392 | 619 |
| Amygdala | 0.117 | 0.064 | 0.069 | 0.608 | -0.009 | 0.243 | 393 | 634 |
| Accumbens | 0.035 | 0.064 | 0.593 | 0.830 | -0.092 | 0.161 | 392 | 628 |
| **Cortical thickness** | | | | | | | | |
| Banks superior temporal sulcus | 0.101 | 0.067 | 0.136 | 0.608 | -0.031 | 0.232 | 360 | 575 |
| Caudal anterior cingulate cortex | 0.010 | 0.064 | 0.877 | 0.939 | -0.115 | 0.135 | 398 | 638 |
| Caudal middle frontal gyrus | 0.099 | 0.064 | 0.124 | 0.608 | -0.027 | 0.225 | 392 | 637 |
| Cuneus | 0.062 | 0.065 | 0.341 | 0.708 | -0.065 | 0.189 | 380 | 627 |
| Entorhinal cortex | -0.034 | 0.067 | 0.617 | 0.830 | -0.165 | 0.098 | 352 | 603 |
| Fusiform gyrus | 0.095 | 0.064 | 0.141 | 0.608 | -0.031 | 0.221 | 394 | 631 |
| Inferior parietal cortex | 0.039 | 0.065 | 0.551 | 0.830 | -0.088 | 0.165 | 383 | 635 |
| Inferior temporal gyrus | -0.007 | 0.065 | 0.919 | 0.943 | -0.135 | 0.121 | 381 | 613 |
| Isthmus cingulate cortex | -0.070 | 0.064 | 0.279 | 0.635 | -0.195 | 0.056 | 393 | 636 |
| Lateral occipital cortex | 0.036 | 0.064 | 0.578 | 0.830 | -0.090 | 0.162 | 391 | 633 |
| Lateral orbitofrontal cortex | 0.133 | 0.065 | 0.039 | 0.608 | 0.007 | 0.260 | 391 | 625 |
| Lingual gyrus | 0.092 | 0.065 | 0.156 | 0.608 | -0.035 | 0.219 | 385 | 635 |
| Medial orbitofrontal gyrus | 0.012 | 0.064 | 0.855 | 0.939 | -0.114 | 0.138 | 395 | 627 |
| Middle temporal gyrus | 0.049 | 0.067 | 0.470 | 0.780 | -0.083 | 0.180 | 363 | 579 |
| Parahippocampal gyrus | 0.060 | 0.064 | 0.354 | 0.708 | -0.066 | 0.185 | 396 | 635 |
| Paracentral lobule | 0.084 | 0.064 | 0.189 | 0.621 | -0.041 | 0.210 | 393 | 639 |
| Pars opercularis | -0.024 | 0.064 | 0.707 | 0.889 | -0.150 | 0.101 | 394 | 637 |
| Pars orbitalis | 0.050 | 0.065 | 0.442 | 0.761 | -0.077 | 0.177 | 386 | 628 |
| Pars triangularis | 0.071 | 0.064 | 0.267 | 0.631 | -0.054 | 0.197 | 396 | 639 |
| Pericalcarine cortex | 0.016 | 0.065 | 0.811 | 0.939 | -0.111 | 0.142 | 385 | 627 |
| Postcentral gyrus | 0.049 | 0.064 | 0.449 | 0.761 | -0.077 | 0.175 | 396 | 624 |
| Posterior cingulate cortex | -0.027 | 0.064 | 0.674 | 0.862 | -0.153 | 0.099 | 393 | 639 |
| Precentral gyrus | 0.117 | 0.064 | 0.071 | 0.608 | -0.009 | 0.243 | 393 | 628 |
| Precuneus | 0.100 | 0.064 | 0.119 | 0.608 | -0.025 | 0.226 | 392 | 636 |
| Rostral anterior cingulate cortex | 0.005 | 0.064 | 0.939 | 0.951 | -0.121 | 0.130 | 395 | 638 |
| Rostral middle frontal gyrus | 0.033 | 0.065 | 0.610 | 0.830 | -0.094 | 0.160 | 387 | 627 |
| Superior frontal gyrus | 0.111 | 0.064 | 0.085 | 0.608 | -0.015 | 0.238 | 391 | 627 |
| Superior parietal gyrus | 0.093 | 0.064 | 0.149 | 0.608 | -0.033 | 0.219 | 396 | 632 |
| Superior temporal gyrus | 0.076 | 0.067 | 0.259 | 0.631 | -0.056 | 0.209 | 353 | 583 |
| Supramarginal gyrus | 0.089 | 0.065 | 0.172 | 0.621 | -0.039 | 0.217 | 378 | 626 |
| Frontal pole | 0.063 | 0.064 | 0.324 | 0.702 | -0.062 | 0.189 | 396 | 638 |
| Temporal pole | -0.008 | 0.064 | 0.907 | 0.943 | -0.133 | 0.118 | 395 | 629 |
| Transverse temporal gyrus | 0.029 | 0.064 | 0.646 | 0.853 | -0.096 | 0.155 | 396 | 639 |
| Insula | 0.037 | 0.064 | 0.565 | 0.830 | -0.089 | 0.163 | 388 | 636 |
| Mean Thickness | 0.085 | 0.064 | 0.182 | 0.621 | -0.040 | 0.211 | 399 | 638 |
| **Cortical surface area** | | | | | | | | |
| Banks superior temporal sulcus | 0.079 | 0.067 | 0.242 | 0.631 | -0.053 | 0.211 | 360 | 567 |
| Caudal anterior cingulate cortex | 0.018 | 0.064 | 0.774 | 0.939 | -0.107 | 0.144 | 396 | 633 |
| Caudal middle frontal gyrus | 0.041 | 0.064 | 0.525 | 0.830 | -0.085 | 0.167 | 394 | 632 |
| Cuneus | 0.080 | 0.065 | 0.223 | 0.631 | -0.048 | 0.207 | 379 | 627 |
| Entorhinal cortex | 0.208 | 0.067 | 0.002 | 0.117 | 0.077 | 0.340 | 354 | 600 |
| Fusiform gyrus | 0.051 | 0.064 | 0.430 | 0.761 | -0.075 | 0.177 | 392 | 633 |
| Inferior parietal cortex | 0.011 | 0.065 | 0.867 | 0.939 | -0.116 | 0.138 | 383 | 635 |
| Inferior temporal gyrus | 0.119 | 0.065 | 0.070 | 0.608 | -0.009 | 0.246 | 382 | 614 |
| Isthmus cingulate cortex | 0.081 | 0.064 | 0.208 | 0.631 | -0.045 | 0.207 | 391 | 633 |
| Lateral occipital cortex | 0.021 | 0.064 | 0.748 | 0.926 | -0.106 | 0.147 | 389 | 631 |
| Lateral orbitofrontal cortex | 0.075 | 0.064 | 0.248 | 0.631 | -0.052 | 0.201 | 391 | 626 |
| Lingual gyrus | 0.096 | 0.065 | 0.140 | 0.608 | -0.031 | 0.222 | 386 | 636 |
| Medial orbitofrontal gyrus | 0.178 | 0.064 | 0.006 | 0.156 | 0.052 | 0.304 | 394 | 626 |
| Middle temporal gyrus | -0.010 | 0.067 | 0.879 | 0.939 | -0.142 | 0.121 | 362 | 579 |
| Parahippocampal gyrus | -0.096 | 0.064 | 0.136 | 0.608 | -0.222 | 0.030 | 393 | 630 |
| Paracentral lobule | -0.035 | 0.064 | 0.587 | 0.830 | -0.161 | 0.091 | 391 | 635 |
| Pars opercularis | 0.015 | 0.064 | 0.812 | 0.939 | -0.111 | 0.141 | 391 | 637 |
| Pars orbitalis | 0.054 | 0.065 | 0.406 | 0.761 | -0.073 | 0.180 | 387 | 632 |
| Pars triangularis | 0.029 | 0.064 | 0.656 | 0.853 | -0.097 | 0.154 | 396 | 636 |
| Pericalcarine cortex | 0.135 | 0.065 | 0.038 | 0.608 | 0.008 | 0.262 | 385 | 628 |
| Postcentral gyrus | 0.102 | 0.064 | 0.114 | 0.608 | -0.024 | 0.228 | 396 | 626 |
| Posterior cingulate cortex | 0.080 | 0.064 | 0.212 | 0.631 | -0.045 | 0.206 | 394 | 637 |
| Precentral gyrus | 0.084 | 0.064 | 0.191 | 0.621 | -0.042 | 0.210 | 395 | 625 |
| Precuneus | 0.078 | 0.064 | 0.227 | 0.631 | -0.048 | 0.204 | 389 | 637 |
| Rostral anterior cingulate cortex | 0.060 | 0.064 | 0.354 | 0.708 | -0.066 | 0.186 | 391 | 636 |
| Rostral middle frontal gyrus | -0.008 | 0.065 | 0.905 | 0.943 | -0.134 | 0.119 | 388 | 627 |
| Superior frontal gyrus | 0.013 | 0.064 | 0.840 | 0.939 | -0.113 | 0.140 | 391 | 624 |
| Superior parietal gyrus | -0.039 | 0.064 | 0.541 | 0.830 | -0.165 | 0.086 | 396 | 635 |
| Superior temporal gyrus | -0.045 | 0.067 | 0.511 | 0.830 | -0.177 | 0.088 | 354 | 580 |
| Supramarginal gyrus | 0.101 | 0.065 | 0.122 | 0.608 | -0.026 | 0.229 | 378 | 624 |
| Frontal pole | -0.012 | 0.064 | 0.846 | 0.939 | -0.138 | 0.113 | 395 | 635 |
| Temporal pole | 0.002 | 0.064 | 0.978 | 0.978 | -0.124 | 0.128 | 394 | 630 |
| Transverse temporal gyrus | 0.033 | 0.064 | 0.603 | 0.830 | -0.092 | 0.159 | 397 | 640 |
| Insula | 0.015 | 0.065 | 0.816 | 0.939 | -0.112 | 0.142 | 385 | 632 |
| Full surface area | 0.072 | 0.064 | 0.259 | 0.631 | -0.053 | 0.198 | 397 | 639 |

Table S10. Differences in regional brain morphology between clinical controls and young people with current suicidal ideation, additionally corrected for type of diagnosis. D: Cohen’s d effect size, SE: standard error; p: p-value, FDR-p: FDR corrected p-value, CI: confidence interval, HC: healthy controls, CC: clinical controls.

| **Region** | **D** | **SE** | **P** | **FDR-p** | **Lower CI** | **Upper CI** | **N ideation** | **N CC** |
| --- | --- | --- | --- | --- | --- | --- | --- | --- |
| **Subcortical volume** | | | | | | | | |
| Ventricle | -0.128 | 0.112 | 0.258 | 0.766 | -0.347 | 0.091 | 136 | 196 |
| Thalamus | 0.086 | 0.081 | 0.294 | 0.766 | -0.073 | 0.245 | 259 | 366 |
| Caudate | 0.136 | 0.081 | 0.100 | 0.766 | -0.024 | 0.295 | 257 | 366 |
| Putamen | 0.090 | 0.083 | 0.285 | 0.766 | -0.073 | 0.252 | 249 | 355 |
| Pallidum | 0.153 | 0.084 | 0.070 | 0.766 | -0.011 | 0.318 | 242 | 347 |
| Hippocampus | 0.058 | 0.081 | 0.482 | 0.814 | -0.102 | 0.217 | 259 | 361 |
| Amygdala | 0.094 | 0.081 | 0.249 | 0.766 | -0.064 | 0.253 | 258 | 372 |
| Accumbens | 0.066 | 0.081 | 0.423 | 0.791 | -0.094 | 0.225 | 257 | 367 |
| **Cortical thickness** | | | | | | | | |
| Banks superior temporal sulcus | 0.111 | 0.084 | 0.192 | 0.766 | -0.054 | 0.276 | 237 | 348 |
| Caudal anterior cingulate cortex | 0.050 | 0.080 | 0.541 | 0.828 | -0.108 | 0.207 | 262 | 377 |
| Caudal middle frontal gyrus | 0.116 | 0.081 | 0.156 | 0.766 | -0.043 | 0.274 | 257 | 378 |
| Cuneus | 0.047 | 0.082 | 0.572 | 0.842 | -0.114 | 0.208 | 246 | 371 |
| Entorhinal cortex | -0.085 | 0.086 | 0.328 | 0.766 | -0.254 | 0.084 | 220 | 346 |
| Fusiform gyrus | 0.117 | 0.081 | 0.151 | 0.766 | -0.042 | 0.275 | 261 | 372 |
| Inferior parietal cortex | 0.087 | 0.082 | 0.291 | 0.766 | -0.073 | 0.247 | 249 | 374 |
| Inferior temporal gyrus | 0.016 | 0.082 | 0.847 | 0.964 | -0.144 | 0.176 | 253 | 368 |
| Isthmus cingulate cortex | -0.006 | 0.081 | 0.942 | 0.964 | -0.165 | 0.153 | 257 | 375 |
| Lateral occipital cortex | 0.084 | 0.081 | 0.301 | 0.766 | -0.074 | 0.243 | 259 | 372 |
| Lateral orbitofrontal cortex | 0.144 | 0.081 | 0.078 | 0.766 | -0.015 | 0.304 | 257 | 372 |
| Lingual gyrus | 0.203 | 0.082 | 0.014 | 0.766 | 0.043 | 0.363 | 252 | 376 |
| Medial orbitofrontal gyrus | 0.007 | 0.081 | 0.933 | 0.964 | -0.152 | 0.166 | 259 | 370 |
| Middle temporal gyrus | 0.025 | 0.084 | 0.768 | 0.964 | -0.139 | 0.189 | 240 | 354 |
| Parahippocampal gyrus | 0.115 | 0.081 | 0.159 | 0.766 | -0.044 | 0.273 | 260 | 373 |
| Paracentral lobule | 0.079 | 0.081 | 0.334 | 0.766 | -0.080 | 0.237 | 259 | 377 |
| Pars opercularis | -0.004 | 0.081 | 0.964 | 0.964 | -0.162 | 0.155 | 258 | 377 |
| Pars orbitalis | 0.072 | 0.081 | 0.377 | 0.791 | -0.087 | 0.232 | 252 | 375 |
| Pars triangularis | 0.081 | 0.081 | 0.319 | 0.766 | -0.077 | 0.239 | 260 | 377 |
| Pericalcarine cortex | 0.054 | 0.082 | 0.513 | 0.814 | -0.106 | 0.214 | 251 | 372 |
| Postcentral gyrus | 0.065 | 0.081 | 0.427 | 0.791 | -0.094 | 0.223 | 261 | 372 |
| Posterior cingulate cortex | -0.018 | 0.081 | 0.822 | 0.964 | -0.176 | 0.140 | 259 | 377 |
| Precentral gyrus | 0.102 | 0.081 | 0.213 | 0.766 | -0.057 | 0.260 | 258 | 374 |
| Precuneus | 0.122 | 0.081 | 0.134 | 0.766 | -0.037 | 0.280 | 259 | 376 |
| Rostral anterior cingulate cortex | 0.011 | 0.081 | 0.894 | 0.964 | -0.147 | 0.169 | 260 | 376 |
| Rostral middle frontal gyrus | 0.027 | 0.081 | 0.738 | 0.964 | -0.132 | 0.186 | 256 | 374 |
| Superior frontal gyrus | 0.135 | 0.081 | 0.098 | 0.766 | -0.024 | 0.294 | 257 | 375 |
| Superior parietal gyrus | 0.090 | 0.081 | 0.266 | 0.766 | -0.068 | 0.249 | 261 | 375 |
| Superior temporal gyrus | 0.015 | 0.085 | 0.856 | 0.964 | -0.151 | 0.182 | 230 | 352 |
| Supramarginal gyrus | 0.109 | 0.082 | 0.190 | 0.766 | -0.053 | 0.270 | 245 | 372 |
| Frontal pole | 0.088 | 0.080 | 0.276 | 0.766 | -0.069 | 0.246 | 262 | 378 |
| Temporal pole | -0.005 | 0.080 | 0.955 | 0.964 | -0.162 | 0.153 | 261 | 379 |
| Transverse temporal gyrus | -0.015 | 0.080 | 0.849 | 0.964 | -0.173 | 0.142 | 261 | 378 |
| Insula | 0.081 | 0.081 | 0.325 | 0.766 | -0.079 | 0.240 | 253 | 376 |
| Mean Thickness | 0.090 | 0.080 | 0.264 | 0.766 | -0.067 | 0.248 | 263 | 377 |
| **Cortical surface area** | | | | | | | | |
| Banks superior temporal sulcus | 0.091 | 0.085 | 0.288 | 0.766 | -0.075 | 0.257 | 237 | 342 |
| Caudal anterior cingulate cortex | 0.026 | 0.081 | 0.746 | 0.964 | -0.132 | 0.185 | 261 | 372 |
| Caudal middle frontal gyrus | 0.081 | 0.081 | 0.320 | 0.766 | -0.077 | 0.240 | 259 | 374 |
| Cuneus | 0.063 | 0.082 | 0.451 | 0.799 | -0.099 | 0.224 | 245 | 371 |
| Entorhinal cortex | 0.170 | 0.086 | 0.051 | 0.766 | 0.001 | 0.339 | 221 | 344 |
| Fusiform gyrus | 0.006 | 0.081 | 0.939 | 0.964 | -0.152 | 0.164 | 260 | 375 |
| Inferior parietal cortex | -0.005 | 0.082 | 0.954 | 0.964 | -0.165 | 0.155 | 250 | 375 |
| Inferior temporal gyrus | 0.150 | 0.082 | 0.069 | 0.766 | -0.010 | 0.311 | 253 | 368 |
| Isthmus cingulate cortex | 0.067 | 0.081 | 0.413 | 0.791 | -0.092 | 0.226 | 256 | 372 |
| Lateral occipital cortex | -0.056 | 0.081 | 0.492 | 0.814 | -0.215 | 0.103 | 259 | 371 |
| Lateral orbitofrontal cortex | 0.052 | 0.081 | 0.522 | 0.814 | -0.106 | 0.211 | 257 | 375 |
| Lingual gyrus | 0.110 | 0.081 | 0.179 | 0.766 | -0.049 | 0.270 | 253 | 377 |
| Medial orbitofrontal gyrus | 0.160 | 0.081 | 0.050 | 0.766 | 0.001 | 0.320 | 258 | 369 |
| Middle temporal gyrus | 0.076 | 0.084 | 0.369 | 0.791 | -0.088 | 0.240 | 239 | 353 |
| Parahippocampal gyrus | -0.031 | 0.081 | 0.705 | 0.964 | -0.190 | 0.128 | 257 | 368 |
| Paracentral lobule | -0.012 | 0.081 | 0.879 | 0.964 | -0.171 | 0.146 | 259 | 377 |
| Pars opercularis | 0.006 | 0.081 | 0.945 | 0.964 | -0.153 | 0.164 | 257 | 378 |
| Pars orbitalis | -0.041 | 0.081 | 0.617 | 0.875 | -0.200 | 0.118 | 253 | 377 |
| Pars triangularis | -0.026 | 0.081 | 0.752 | 0.964 | -0.184 | 0.132 | 261 | 377 |
| Pericalcarine cortex | 0.162 | 0.082 | 0.050 | 0.766 | 0.002 | 0.323 | 251 | 372 |
| Postcentral gyrus | 0.092 | 0.081 | 0.261 | 0.766 | -0.066 | 0.250 | 261 | 374 |
| Posterior cingulate cortex | 0.067 | 0.081 | 0.408 | 0.791 | -0.091 | 0.226 | 260 | 376 |
| Precentral gyrus | 0.064 | 0.081 | 0.436 | 0.791 | -0.095 | 0.222 | 260 | 371 |
| Precuneus | 0.054 | 0.081 | 0.510 | 0.814 | -0.105 | 0.212 | 258 | 375 |
| Rostral anterior cingulate cortex | 0.047 | 0.081 | 0.568 | 0.842 | -0.112 | 0.205 | 258 | 375 |
| Rostral middle frontal gyrus | -0.030 | 0.081 | 0.718 | 0.964 | -0.188 | 0.129 | 257 | 374 |
| Superior frontal gyrus | -0.005 | 0.081 | 0.949 | 0.964 | -0.164 | 0.153 | 258 | 374 |
| Superior parietal gyrus | -0.016 | 0.081 | 0.847 | 0.964 | -0.174 | 0.142 | 261 | 377 |
| Superior temporal gyrus | -0.044 | 0.085 | 0.610 | 0.875 | -0.210 | 0.123 | 231 | 347 |
| Supramarginal gyrus | 0.069 | 0.082 | 0.411 | 0.791 | -0.093 | 0.230 | 245 | 369 |
| Frontal pole | -0.011 | 0.081 | 0.896 | 0.964 | -0.168 | 0.147 | 261 | 377 |
| Temporal pole | -0.092 | 0.080 | 0.256 | 0.766 | -0.250 | 0.065 | 261 | 379 |
| Transverse temporal gyrus | 0.011 | 0.080 | 0.890 | 0.964 | -0.146 | 0.169 | 263 | 378 |
| Insula | -0.053 | 0.082 | 0.518 | 0.814 | -0.213 | 0.107 | 252 | 373 |
| Full surface area | 0.065 | 0.080 | 0.421 | 0.791 | -0.092 | 0.223 | 262 | 378 |

Table 11. Differences in regional brain morphology between healthy controls and young people with current suicidal ideation **in men only**.

D: Cohen’s d effect size, SE: standard error; p: p-value, FDR-p: FDR corrected p-value, CI: confidence interval, HC: healthy controls, CC: clinical controls.

| **Region** | **D** | **SE** | **P** | **FDR-p** | **Lower CI** | **Upper CI** | **N ideation** | **N HC** |
| --- | --- | --- | --- | --- | --- | --- | --- | --- |
| **Subcortical volume** | | | | | | | | |
| Ventricle | 0.179 | 0.203 | 0.389 | 0.994 | -0.219 | 0.576 | 46 | 52 |
| Thalamus | 0.190 | 0.120 | 0.118 | 0.979 | -0.046 | 0.426 | 112 | 181 |
| Caudate | 0.120 | 0.122 | 0.330 | 0.994 | -0.119 | 0.358 | 109 | 178 |
| Putamen | 0.209 | 0.124 | 0.096 | 0.979 | -0.034 | 0.452 | 110 | 160 |
| Pallidum | 0.134 | 0.127 | 0.297 | 0.994 | -0.115 | 0.382 | 102 | 159 |
| Hippocampus | -0.090 | 0.121 | 0.459 | 0.994 | -0.327 | 0.147 | 109 | 185 |
| Amygdala | 0.186 | 0.121 | 0.128 | 0.979 | -0.051 | 0.422 | 112 | 180 |
| Accumbens | -0.010 | 0.121 | 0.938 | 0.994 | -0.247 | 0.227 | 111 | 178 |
| **Cortical thickness** | | | | | | | | |
| Banks superior temporal sulcus | 0.082 | 0.132 | 0.539 | 0.994 | -0.177 | 0.340 | 95 | 147 |
| Caudal anterior cingulate cortex | 0.050 | 0.120 | 0.678 | 0.994 | -0.185 | 0.286 | 113 | 179 |
| Caudal middle frontal gyrus | -0.051 | 0.120 | 0.676 | 0.994 | -0.287 | 0.185 | 111 | 182 |
| Cuneus | -0.029 | 0.125 | 0.816 | 0.994 | -0.274 | 0.215 | 104 | 168 |
| Entorhinal cortex | -0.021 | 0.129 | 0.870 | 0.994 | -0.274 | 0.232 | 95 | 163 |
| Fusiform gyrus | 0.208 | 0.121 | 0.088 | 0.979 | -0.030 | 0.445 | 110 | 181 |
| Inferior parietal cortex | -0.122 | 0.123 | 0.327 | 0.994 | -0.363 | 0.120 | 105 | 177 |
| Inferior temporal gyrus | 0.002 | 0.123 | 0.986 | 0.994 | -0.239 | 0.244 | 104 | 180 |
| Isthmus cingulate cortex | -0.121 | 0.121 | 0.321 | 0.994 | -0.358 | 0.116 | 111 | 179 |
| Lateral occipital cortex | 0.031 | 0.122 | 0.803 | 0.994 | -0.208 | 0.269 | 109 | 178 |
| Lateral orbitofrontal cortex | 0.133 | 0.121 | 0.275 | 0.994 | -0.104 | 0.370 | 109 | 183 |
| Lingual gyrus | 0.004 | 0.121 | 0.973 | 0.994 | -0.234 | 0.242 | 108 | 182 |
| Medial orbitofrontal gyrus | 0.063 | 0.120 | 0.600 | 0.994 | -0.172 | 0.299 | 113 | 180 |
| Middle temporal gyrus | 0.039 | 0.127 | 0.760 | 0.994 | -0.211 | 0.289 | 99 | 163 |
| Parahippocampal gyrus | 0.017 | 0.120 | 0.886 | 0.994 | -0.218 | 0.252 | 112 | 184 |
| Paracentral lobule | -0.024 | 0.121 | 0.842 | 0.994 | -0.260 | 0.212 | 111 | 181 |
| Pars opercularis | -0.013 | 0.120 | 0.913 | 0.994 | -0.249 | 0.223 | 110 | 185 |
| Pars orbitalis | 0.083 | 0.122 | 0.500 | 0.994 | -0.156 | 0.322 | 107 | 181 |
| Pars triangularis | 0.092 | 0.120 | 0.445 | 0.994 | -0.143 | 0.328 | 113 | 179 |
| Pericalcarine cortex | -0.013 | 0.122 | 0.918 | 0.994 | -0.252 | 0.227 | 109 | 173 |
| Postcentral gyrus | -0.029 | 0.120 | 0.810 | 0.994 | -0.265 | 0.207 | 112 | 179 |
| Posterior cingulate cortex | 0.022 | 0.120 | 0.854 | 0.994 | -0.213 | 0.257 | 112 | 183 |
| Precentral gyrus | -0.059 | 0.121 | 0.627 | 0.994 | -0.296 | 0.178 | 110 | 180 |
| Precuneus | 0.055 | 0.121 | 0.654 | 0.994 | -0.183 | 0.293 | 109 | 180 |
| Rostral anterior cingulate cortex | 0.058 | 0.122 | 0.639 | 0.994 | -0.182 | 0.297 | 110 | 172 |
| Rostral middle frontal gyrus | 0.017 | 0.122 | 0.888 | 0.994 | -0.222 | 0.256 | 108 | 178 |
| Superior frontal gyrus | 0.004 | 0.121 | 0.972 | 0.994 | -0.233 | 0.242 | 110 | 179 |
| Superior parietal gyrus | -0.028 | 0.120 | 0.815 | 0.994 | -0.264 | 0.208 | 111 | 183 |
| Superior temporal gyrus | -0.010 | 0.131 | 0.938 | 0.994 | -0.268 | 0.247 | 93 | 153 |
| Supramarginal gyrus | -0.014 | 0.125 | 0.910 | 0.994 | -0.260 | 0.232 | 101 | 171 |
| Frontal pole | 0.177 | 0.121 | 0.145 | 0.979 | -0.059 | 0.414 | 111 | 181 |
| Temporal pole | -0.129 | 0.120 | 0.287 | 0.994 | -0.365 | 0.107 | 113 | 179 |
| Transverse temporal gyrus | 0.193 | 0.120 | 0.110 | 0.979 | -0.042 | 0.428 | 112 | 185 |
| Insula | 0.018 | 0.125 | 0.887 | 0.994 | -0.227 | 0.262 | 107 | 161 |
| Mean Thickness | 0.024 | 0.119 | 0.838 | 0.994 | -0.209 | 0.258 | 114 | 186 |
| **Cortical surface area** | | | | | | | | |
| Banks superior temporal sulcus | 0.052 | 0.132 | 0.694 | 0.994 | -0.206 | 0.310 | 95 | 147 |
| Caudal anterior cingulate cortex | 0.074 | 0.121 | 0.543 | 0.994 | -0.162 | 0.310 | 112 | 179 |
| Caudal middle frontal gyrus | -0.186 | 0.121 | 0.128 | 0.979 | -0.423 | 0.051 | 110 | 182 |
| Cuneus | 0.003 | 0.125 | 0.983 | 0.994 | -0.243 | 0.248 | 103 | 167 |
| Entorhinal cortex | 0.072 | 0.129 | 0.581 | 0.994 | -0.181 | 0.325 | 96 | 160 |
| Fusiform gyrus | 0.109 | 0.121 | 0.372 | 0.994 | -0.129 | 0.347 | 110 | 179 |
| Inferior parietal cortex | 0.231 | 0.123 | 0.064 | 0.979 | -0.011 | 0.472 | 106 | 176 |
| Inferior temporal gyrus | 0.060 | 0.123 | 0.631 | 0.994 | -0.182 | 0.302 | 104 | 178 |
| Isthmus cingulate cortex | -0.072 | 0.121 | 0.557 | 0.994 | -0.309 | 0.165 | 110 | 180 |
| Lateral occipital cortex | 0.217 | 0.123 | 0.080 | 0.979 | -0.024 | 0.457 | 108 | 176 |
| Lateral orbitofrontal cortex | 0.082 | 0.121 | 0.503 | 0.994 | -0.156 | 0.320 | 109 | 180 |
| Lingual gyrus | 0.027 | 0.122 | 0.828 | 0.994 | -0.212 | 0.266 | 107 | 182 |
| Medial orbitofrontal gyrus | 0.063 | 0.120 | 0.602 | 0.994 | -0.172 | 0.299 | 112 | 182 |
| Middle temporal gyrus | -0.001 | 0.128 | 0.994 | 0.994 | -0.251 | 0.249 | 99 | 161 |
| Parahippocampal gyrus | -0.073 | 0.121 | 0.553 | 0.994 | -0.310 | 0.165 | 109 | 181 |
| Paracentral lobule | -0.268 | 0.122 | 0.030 | 0.979 | -0.507 | -0.028 | 108 | 181 |
| Pars opercularis | 0.038 | 0.121 | 0.757 | 0.994 | -0.199 | 0.274 | 110 | 182 |
| Pars orbitalis | -0.021 | 0.123 | 0.863 | 0.994 | -0.262 | 0.220 | 105 | 179 |
| Pars triangularis | -0.128 | 0.120 | 0.294 | 0.994 | -0.364 | 0.108 | 113 | 178 |
| Pericalcarine cortex | 0.065 | 0.122 | 0.600 | 0.994 | -0.175 | 0.305 | 109 | 172 |
| Postcentral gyrus | 0.098 | 0.121 | 0.421 | 0.994 | -0.138 | 0.335 | 112 | 178 |
| Posterior cingulate cortex | 0.054 | 0.119 | 0.654 | 0.994 | -0.180 | 0.288 | 114 | 182 |
| Precentral gyrus | -0.023 | 0.121 | 0.849 | 0.994 | -0.261 | 0.214 | 110 | 180 |
| Precuneus | -0.035 | 0.121 | 0.773 | 0.994 | -0.273 | 0.202 | 109 | 181 |
| Rostral anterior cingulate cortex | 0.206 | 0.122 | 0.096 | 0.979 | -0.034 | 0.446 | 109 | 174 |
| Rostral middle frontal gyrus | -0.025 | 0.122 | 0.837 | 0.994 | -0.264 | 0.213 | 109 | 178 |
| Superior frontal gyrus | -0.128 | 0.122 | 0.299 | 0.994 | -0.366 | 0.111 | 110 | 177 |
| Superior parietal gyrus | -0.173 | 0.121 | 0.156 | 0.979 | -0.409 | 0.064 | 111 | 182 |
| Superior temporal gyrus | 0.145 | 0.132 | 0.275 | 0.994 | -0.113 | 0.403 | 93 | 153 |
| Supramarginal gyrus | 0.157 | 0.126 | 0.217 | 0.994 | -0.090 | 0.403 | 101 | 170 |
| Frontal pole | 0.067 | 0.121 | 0.580 | 0.994 | -0.169 | 0.304 | 111 | 179 |
| Temporal pole | 0.170 | 0.121 | 0.163 | 0.979 | -0.067 | 0.406 | 112 | 179 |
| Transverse temporal gyrus | -0.042 | 0.120 | 0.728 | 0.994 | -0.277 | 0.193 | 112 | 185 |
| Insula | -0.063 | 0.125 | 0.617 | 0.994 | -0.309 | 0.182 | 106 | 160 |
| Full surface area | 0.037 | 0.119 | 0.761 | 0.994 | -0.197 | 0.271 | 113 | 185 |

Table S12. Differences in regional brain morphology between clinical controls and young people with current suicidal ideation **in men only**.

D: Cohen’s d effect size, SE: standard error; p: p-value, FDR-p: FDR corrected p-value, CI: confidence interval, HC: healthy controls, CC: clinical controls.

| **Region** | **D** | **SE** | **P** | **FDR-p** | **Lower CI** | **Upper CI** | **N ideation** | **N CC** |
| --- | --- | --- | --- | --- | --- | --- | --- | --- |
| **Subcortical volume** | | | | | | | | |
| Ventricle | -0.139 | 0.190 | 0.475 | 0.817 | -0.511 | 0.234 | 46 | 70 |
| Thalamus | 0.283 | 0.126 | 0.027 | 0.432 | 0.036 | 0.529 | 112 | 149 |
| Caudate | 0.104 | 0.126 | 0.415 | 0.817 | -0.143 | 0.350 | 109 | 151 |
| Putamen | 0.293 | 0.128 | 0.025 | 0.432 | 0.041 | 0.544 | 110 | 138 |
| Pallidum | 0.041 | 0.131 | 0.760 | 0.872 | -0.217 | 0.298 | 102 | 134 |
| Hippocampus | 0.091 | 0.127 | 0.481 | 0.817 | -0.158 | 0.340 | 109 | 144 |
| Amygdala | 0.105 | 0.125 | 0.408 | 0.817 | -0.141 | 0.351 | 112 | 148 |
| Accumbens | 0.113 | 0.125 | 0.375 | 0.817 | -0.133 | 0.358 | 111 | 150 |
| **Cortical thickness** | | | | | | | | |
| Banks superior temporal sulcus | 0.233 | 0.134 | 0.084 | 0.697 | -0.029 | 0.496 | 95 | 137 |
| Caudal anterior cingulate cortex | 0.041 | 0.124 | 0.745 | 0.870 | -0.203 | 0.284 | 113 | 151 |
| Caudal middle frontal gyrus | 0.082 | 0.126 | 0.518 | 0.819 | -0.165 | 0.328 | 111 | 148 |
| Cuneus | 0.153 | 0.129 | 0.240 | 0.817 | -0.100 | 0.406 | 104 | 142 |
| Entorhinal cortex | 0.011 | 0.133 | 0.933 | 0.970 | -0.250 | 0.272 | 95 | 139 |
| Fusiform gyrus | 0.350 | 0.127 | 0.006 | 0.432 | 0.100 | 0.600 | 110 | 145 |
| Inferior parietal cortex | 0.093 | 0.128 | 0.469 | 0.817 | -0.157 | 0.344 | 105 | 147 |
| Inferior temporal gyrus | 0.191 | 0.130 | 0.143 | 0.791 | -0.063 | 0.446 | 104 | 140 |
| Isthmus cingulate cortex | 0.061 | 0.125 | 0.628 | 0.823 | -0.184 | 0.306 | 111 | 150 |
| Lateral occipital cortex | 0.227 | 0.126 | 0.074 | 0.697 | -0.020 | 0.475 | 109 | 150 |
| Lateral orbitofrontal cortex | 0.282 | 0.127 | 0.028 | 0.432 | 0.033 | 0.532 | 109 | 145 |
| Lingual gyrus | 0.106 | 0.127 | 0.404 | 0.817 | -0.142 | 0.355 | 108 | 148 |
| Medial orbitofrontal gyrus | 0.106 | 0.126 | 0.401 | 0.817 | -0.140 | 0.352 | 113 | 145 |
| Middle temporal gyrus | 0.246 | 0.133 | 0.067 | 0.697 | -0.015 | 0.506 | 99 | 134 |
| Parahippocampal gyrus | 0.041 | 0.126 | 0.748 | 0.870 | -0.206 | 0.287 | 112 | 145 |
| Paracentral lobule | 0.103 | 0.125 | 0.413 | 0.817 | -0.142 | 0.349 | 111 | 150 |
| Pars opercularis | 0.077 | 0.126 | 0.541 | 0.819 | -0.169 | 0.324 | 110 | 149 |
| Pars orbitalis | 0.103 | 0.128 | 0.424 | 0.817 | -0.147 | 0.353 | 107 | 145 |
| Pars triangularis | 0.162 | 0.125 | 0.198 | 0.791 | -0.083 | 0.406 | 113 | 150 |
| Pericalcarine cortex | 0.020 | 0.127 | 0.874 | 0.947 | -0.229 | 0.270 | 109 | 143 |
| Postcentral gyrus | 0.111 | 0.126 | 0.385 | 0.817 | -0.137 | 0.359 | 112 | 142 |
| Posterior cingulate cortex | 0.064 | 0.125 | 0.613 | 0.823 | -0.181 | 0.308 | 112 | 151 |
| Precentral gyrus | 0.094 | 0.127 | 0.464 | 0.817 | -0.155 | 0.343 | 110 | 142 |
| Precuneus | 0.307 | 0.126 | 0.016 | 0.432 | 0.059 | 0.555 | 109 | 151 |
| Rostral anterior cingulate cortex | 0.028 | 0.126 | 0.825 | 0.906 | -0.218 | 0.274 | 110 | 149 |
| Rostral middle frontal gyrus | 0.061 | 0.127 | 0.635 | 0.823 | -0.188 | 0.310 | 108 | 145 |
| Superior frontal gyrus | 0.125 | 0.127 | 0.330 | 0.817 | -0.124 | 0.373 | 110 | 143 |
| Superior parietal gyrus | 0.166 | 0.126 | 0.191 | 0.791 | -0.081 | 0.413 | 111 | 147 |
| Superior temporal gyrus | 0.141 | 0.135 | 0.302 | 0.817 | -0.125 | 0.406 | 93 | 133 |
| Supramarginal gyrus | 0.223 | 0.130 | 0.089 | 0.697 | -0.032 | 0.478 | 101 | 144 |
| Frontal pole | 0.079 | 0.126 | 0.534 | 0.819 | -0.167 | 0.325 | 111 | 148 |
| Temporal pole | -0.013 | 0.125 | 0.919 | 0.969 | -0.259 | 0.233 | 113 | 145 |
| Transverse temporal gyrus | 0.138 | 0.125 | 0.273 | 0.817 | -0.107 | 0.382 | 112 | 152 |
| Insula | 0.043 | 0.127 | 0.736 | 0.870 | -0.205 | 0.291 | 107 | 149 |
| Mean Thickness | 0.168 | 0.124 | 0.180 | 0.791 | -0.076 | 0.412 | 114 | 151 |
| **Cortical surface area** | | | | | | | | |
| Banks superior temporal sulcus | 0.053 | 0.134 | 0.698 | 0.870 | -0.210 | 0.315 | 95 | 135 |
| Caudal anterior cingulate cortex | 0.099 | 0.125 | 0.435 | 0.817 | -0.146 | 0.345 | 112 | 148 |
| Caudal middle frontal gyrus | -0.002 | 0.126 | 0.987 | 0.987 | -0.250 | 0.246 | 110 | 145 |
| Cuneus | 0.152 | 0.129 | 0.246 | 0.817 | -0.102 | 0.406 | 103 | 143 |
| Entorhinal cortex | 0.079 | 0.133 | 0.556 | 0.819 | -0.182 | 0.340 | 96 | 137 |
| Fusiform gyrus | 0.117 | 0.127 | 0.364 | 0.817 | -0.132 | 0.365 | 110 | 144 |
| Inferior parietal cortex | 0.074 | 0.127 | 0.567 | 0.819 | -0.176 | 0.323 | 106 | 148 |
| Inferior temporal gyrus | 0.004 | 0.129 | 0.976 | 0.987 | -0.250 | 0.258 | 104 | 140 |
| Isthmus cingulate cortex | -0.168 | 0.126 | 0.189 | 0.791 | -0.414 | 0.079 | 110 | 149 |
| Lateral occipital cortex | 0.090 | 0.127 | 0.481 | 0.817 | -0.158 | 0.338 | 108 | 148 |
| Lateral orbitofrontal cortex | -0.095 | 0.127 | 0.460 | 0.817 | -0.344 | 0.153 | 109 | 145 |
| Lingual gyrus | 0.150 | 0.127 | 0.242 | 0.817 | -0.098 | 0.399 | 107 | 149 |
| Medial orbitofrontal gyrus | 0.125 | 0.126 | 0.328 | 0.817 | -0.123 | 0.373 | 112 | 142 |
| Middle temporal gyrus | -0.070 | 0.132 | 0.602 | 0.823 | -0.329 | 0.189 | 99 | 136 |
| Parahippocampal gyrus | -0.084 | 0.127 | 0.515 | 0.819 | -0.333 | 0.166 | 109 | 143 |
| Paracentral lobule | -0.172 | 0.127 | 0.179 | 0.791 | -0.421 | 0.077 | 108 | 147 |
| Pars opercularis | -0.017 | 0.126 | 0.897 | 0.959 | -0.263 | 0.230 | 110 | 148 |
| Pars orbitalis | 0.091 | 0.128 | 0.482 | 0.817 | -0.160 | 0.342 | 105 | 145 |
| Pars triangularis | 0.059 | 0.125 | 0.644 | 0.823 | -0.186 | 0.304 | 113 | 148 |
| Pericalcarine cortex | 0.192 | 0.127 | 0.138 | 0.791 | -0.058 | 0.442 | 109 | 143 |
| Postcentral gyrus | 0.128 | 0.126 | 0.319 | 0.817 | -0.120 | 0.375 | 112 | 143 |
| Posterior cingulate cortex | -0.045 | 0.124 | 0.719 | 0.870 | -0.289 | 0.198 | 114 | 150 |
| Precentral gyrus | 0.175 | 0.128 | 0.176 | 0.791 | -0.076 | 0.425 | 110 | 139 |
| Precuneus | 0.080 | 0.126 | 0.529 | 0.819 | -0.166 | 0.327 | 109 | 150 |
| Rostral anterior cingulate cortex | 0.163 | 0.126 | 0.203 | 0.791 | -0.085 | 0.410 | 109 | 150 |
| Rostral middle frontal gyrus | 0.064 | 0.127 | 0.621 | 0.823 | -0.185 | 0.313 | 109 | 144 |
| Superior frontal gyrus | -0.044 | 0.127 | 0.734 | 0.870 | -0.293 | 0.206 | 110 | 141 |
| Superior parietal gyrus | -0.229 | 0.126 | 0.073 | 0.697 | -0.476 | 0.018 | 111 | 148 |
| Superior temporal gyrus | 0.081 | 0.135 | 0.554 | 0.819 | -0.184 | 0.346 | 93 | 134 |
| Supramarginal gyrus | 0.171 | 0.130 | 0.196 | 0.791 | -0.084 | 0.425 | 101 | 145 |
| Frontal pole | 0.005 | 0.126 | 0.971 | 0.987 | -0.241 | 0.251 | 111 | 148 |
| Temporal pole | -0.101 | 0.126 | 0.430 | 0.817 | -0.348 | 0.146 | 112 | 144 |
| Transverse temporal gyrus | -0.031 | 0.125 | 0.808 | 0.901 | -0.275 | 0.214 | 112 | 151 |
| Insula | -0.031 | 0.127 | 0.808 | 0.901 | -0.281 | 0.218 | 106 | 148 |
| Full surface area | 0.059 | 0.125 | 0.642 | 0.823 | -0.186 | 0.303 | 113 | 150 |

Table 13. Differences in regional brain morphology between healthy controls and young people with current suicidal ideation **in women only**.

D: Cohen’s d effect size, SE: standard error; p: p-value, FDR-p: FDR corrected p-value, CI: confidence interval, HC: healthy controls, CC: clinical controls.

| **Region** | **D** | **SE** | **P** | **FDR-p** | **Lower CI** | **Upper CI** | **N ideation** | **N HC** |
| --- | --- | --- | --- | --- | --- | --- | --- | --- |
| **Subcortical volume** | | | | | | | | |
| Ventricle | -0.080 | 0.117 | 0.499 | 0.865 | -0.308 | 0.149 | 117 | 198 |
| Thalamus | -0.104 | 0.086 | 0.233 | 0.658 | -0.273 | 0.066 | 212 | 367 |
| Caudate | 0.043 | 0.086 | 0.618 | 0.877 | -0.126 | 0.212 | 212 | 366 |
| Putamen | -0.109 | 0.088 | 0.219 | 0.658 | -0.282 | 0.064 | 202 | 351 |
| Pallidum | 0.084 | 0.089 | 0.349 | 0.777 | -0.091 | 0.258 | 204 | 334 |
| Hippocampus | -0.141 | 0.086 | 0.104 | 0.477 | -0.310 | 0.028 | 212 | 370 |
| Amygdala | -0.074 | 0.086 | 0.393 | 0.806 | -0.244 | 0.095 | 211 | 366 |
| Accumbens | 0.023 | 0.086 | 0.793 | 0.908 | -0.146 | 0.192 | 210 | 371 |
| **Cortical thickness** | | | | | | | | |
| Banks superior temporal sulcus | -0.058 | 0.089 | 0.515 | 0.873 | -0.232 | 0.116 | 198 | 356 |
| Caudal anterior cingulate cortex | 0.045 | 0.086 | 0.603 | 0.877 | -0.123 | 0.213 | 214 | 374 |
| Caudal middle frontal gyrus | -0.147 | 0.086 | 0.089 | 0.477 | -0.316 | 0.022 | 210 | 375 |
| Cuneus | -0.020 | 0.087 | 0.816 | 0.909 | -0.191 | 0.150 | 206 | 370 |
| Entorhinal cortex | -0.004 | 0.092 | 0.969 | 0.990 | -0.184 | 0.177 | 187 | 320 |
| Fusiform gyrus | -0.096 | 0.086 | 0.262 | 0.680 | -0.264 | 0.071 | 213 | 381 |
| Inferior parietal cortex | -0.187 | 0.087 | 0.031 | 0.477 | -0.357 | -0.017 | 207 | 377 |
| Inferior temporal gyrus | -0.086 | 0.086 | 0.321 | 0.736 | -0.255 | 0.083 | 208 | 377 |
| Isthmus cingulate cortex | -0.047 | 0.086 | 0.589 | 0.877 | -0.215 | 0.122 | 211 | 380 |
| Lateral occipital cortex | -0.255 | 0.086 | 0.003 | 0.170 | -0.424 | -0.086 | 211 | 381 |
| Lateral orbitofrontal cortex | 0.043 | 0.086 | 0.614 | 0.877 | -0.125 | 0.212 | 212 | 379 |
| Lingual gyrus | 0.049 | 0.087 | 0.574 | 0.877 | -0.121 | 0.218 | 206 | 379 |
| Medial orbitofrontal gyrus | -0.112 | 0.086 | 0.193 | 0.658 | -0.281 | 0.056 | 213 | 371 |
| Middle temporal gyrus | -0.094 | 0.088 | 0.290 | 0.706 | -0.267 | 0.079 | 198 | 367 |
| Parahippocampal gyrus | 0.050 | 0.086 | 0.563 | 0.877 | -0.118 | 0.217 | 213 | 380 |
| Paracentral lobule | -0.227 | 0.086 | 0.009 | 0.225 | -0.396 | -0.058 | 211 | 380 |
| Pars opercularis | -0.102 | 0.086 | 0.236 | 0.658 | -0.270 | 0.066 | 213 | 374 |
| Pars orbitalis | -0.026 | 0.086 | 0.767 | 0.908 | -0.195 | 0.144 | 208 | 378 |
| Pars triangularis | -0.157 | 0.086 | 0.069 | 0.477 | -0.326 | 0.012 | 212 | 375 |
| Pericalcarine cortex | 0.036 | 0.087 | 0.676 | 0.908 | -0.134 | 0.207 | 206 | 369 |
| Postcentral gyrus | -0.112 | 0.086 | 0.194 | 0.658 | -0.281 | 0.057 | 213 | 370 |
| Posterior cingulate cortex | -0.153 | 0.086 | 0.078 | 0.477 | -0.322 | 0.017 | 210 | 375 |
| Precentral gyrus | -0.123 | 0.086 | 0.155 | 0.577 | -0.291 | 0.046 | 212 | 374 |
| Precuneus | -0.124 | 0.086 | 0.150 | 0.577 | -0.292 | 0.044 | 212 | 380 |
| Rostral anterior cingulate cortex | -0.092 | 0.086 | 0.288 | 0.706 | -0.260 | 0.077 | 214 | 369 |
| Rostral middle frontal gyrus | -0.142 | 0.087 | 0.103 | 0.477 | -0.311 | 0.028 | 209 | 370 |
| Superior frontal gyrus | -0.152 | 0.086 | 0.078 | 0.477 | -0.321 | 0.016 | 211 | 377 |
| Superior parietal gyrus | -0.152 | 0.086 | 0.077 | 0.477 | -0.319 | 0.016 | 214 | 379 |
| Superior temporal gyrus | -0.154 | 0.089 | 0.086 | 0.477 | -0.330 | 0.021 | 194 | 354 |
| Supramarginal gyrus | -0.142 | 0.087 | 0.105 | 0.477 | -0.313 | 0.029 | 206 | 367 |
| Frontal pole | 0.075 | 0.086 | 0.384 | 0.806 | -0.093 | 0.242 | 214 | 379 |
| Temporal pole | 0.031 | 0.086 | 0.718 | 0.908 | -0.137 | 0.199 | 212 | 377 |
| Transverse temporal gyrus | -0.040 | 0.085 | 0.639 | 0.890 | -0.208 | 0.127 | 214 | 380 |
| Insula | -0.033 | 0.087 | 0.704 | 0.908 | -0.204 | 0.138 | 210 | 349 |
| Mean Thickness | -0.173 | 0.086 | 0.043 | 0.477 | -0.341 | -0.006 | 214 | 382 |
| **Cortical surface area** | | | | | | | | |
| Banks superior temporal sulcus | -0.143 | 0.089 | 0.110 | 0.477 | -0.317 | 0.031 | 199 | 350 |
| Caudal anterior cingulate cortex | 0.003 | 0.086 | 0.970 | 0.990 | -0.165 | 0.171 | 214 | 374 |
| Caudal middle frontal gyrus | -0.038 | 0.086 | 0.659 | 0.902 | -0.206 | 0.130 | 213 | 376 |
| Cuneus | -0.065 | 0.087 | 0.457 | 0.828 | -0.236 | 0.105 | 206 | 369 |
| Entorhinal cortex | 0.148 | 0.092 | 0.108 | 0.477 | -0.032 | 0.328 | 189 | 322 |
| Fusiform gyrus | -0.021 | 0.086 | 0.804 | 0.908 | -0.189 | 0.147 | 212 | 380 |
| Inferior parietal cortex | -0.248 | 0.087 | 0.004 | 0.170 | -0.418 | -0.078 | 208 | 375 |
| Inferior temporal gyrus | -0.022 | 0.086 | 0.795 | 0.908 | -0.192 | 0.147 | 209 | 376 |
| Isthmus cingulate cortex | 0.006 | 0.086 | 0.945 | 0.990 | -0.162 | 0.174 | 212 | 381 |
| Lateral occipital cortex | -0.079 | 0.086 | 0.360 | 0.780 | -0.247 | 0.089 | 212 | 381 |
| Lateral orbitofrontal cortex | -0.025 | 0.086 | 0.774 | 0.908 | -0.193 | 0.143 | 212 | 378 |
| Lingual gyrus | -0.002 | 0.086 | 0.981 | 0.990 | -0.171 | 0.167 | 208 | 379 |
| Medial orbitofrontal gyrus | 0.067 | 0.086 | 0.435 | 0.808 | -0.101 | 0.236 | 213 | 371 |
| Middle temporal gyrus | -0.129 | 0.088 | 0.147 | 0.577 | -0.302 | 0.045 | 198 | 364 |
| Parahippocampal gyrus | -0.210 | 0.086 | 0.015 | 0.287 | -0.379 | -0.042 | 213 | 380 |
| Paracentral lobule | -0.028 | 0.086 | 0.742 | 0.908 | -0.196 | 0.140 | 213 | 379 |
| Pars opercularis | -0.097 | 0.086 | 0.261 | 0.680 | -0.266 | 0.072 | 211 | 374 |
| Pars orbitalis | -0.047 | 0.086 | 0.586 | 0.877 | -0.215 | 0.121 | 211 | 380 |
| Pars triangularis | -0.051 | 0.086 | 0.554 | 0.877 | -0.219 | 0.117 | 212 | 379 |
| Pericalcarine cortex | 0.014 | 0.087 | 0.868 | 0.954 | -0.156 | 0.185 | 206 | 368 |
| Postcentral gyrus | -0.007 | 0.086 | 0.937 | 0.990 | -0.175 | 0.162 | 213 | 370 |
| Posterior cingulate cortex | -0.030 | 0.086 | 0.725 | 0.908 | -0.199 | 0.138 | 210 | 377 |
| Precentral gyrus | -0.027 | 0.086 | 0.754 | 0.908 | -0.195 | 0.141 | 214 | 374 |
| Precuneus | -0.104 | 0.086 | 0.227 | 0.658 | -0.272 | 0.064 | 212 | 380 |
| Rostral anterior cingulate cortex | 0.009 | 0.086 | 0.914 | 0.990 | -0.159 | 0.178 | 213 | 370 |
| Rostral middle frontal gyrus | -0.070 | 0.086 | 0.416 | 0.808 | -0.239 | 0.099 | 210 | 374 |
| Superior frontal gyrus | -0.069 | 0.086 | 0.424 | 0.808 | -0.237 | 0.099 | 212 | 377 |
| Superior parietal gyrus | -0.001 | 0.086 | 0.990 | 0.990 | -0.169 | 0.166 | 214 | 379 |
| Superior temporal gyrus | -0.144 | 0.090 | 0.109 | 0.477 | -0.320 | 0.031 | 195 | 348 |
| Supramarginal gyrus | 0.046 | 0.087 | 0.598 | 0.877 | -0.125 | 0.217 | 206 | 365 |
| Frontal pole | -0.069 | 0.085 | 0.425 | 0.808 | -0.236 | 0.099 | 214 | 381 |
| Temporal pole | 0.026 | 0.086 | 0.766 | 0.908 | -0.142 | 0.194 | 213 | 377 |
| Transverse temporal gyrus | 0.062 | 0.085 | 0.472 | 0.837 | -0.106 | 0.229 | 214 | 381 |
| Insula | -0.106 | 0.088 | 0.228 | 0.658 | -0.278 | 0.066 | 209 | 348 |
| Full surface area | -0.088 | 0.085 | 0.306 | 0.724 | -0.255 | 0.080 | 214 | 382 |

Table S14. Differences in regional brain morphology between clinical controls and young people current suicidal ideation **in women only**.

D: Cohen’s d effect size, SE: standard error; p: p-value, FDR-p: FDR corrected p-value, CI: confidence interval, HC: healthy controls, CC: clinical controls.

| **Region** | **D** | **SE** | **P** | **FDR-p** | **Lower CI** | **Upper CI** | **N ideation** | **N CC** |
| --- | --- | --- | --- | --- | --- | --- | --- | --- |
| **Subcortical volume** | | | | | | | | |
| Ventricle | -0.179 | 0.120 | 0.141 | 0.920 | -0.415 | 0.057 | 117 | 170 |
| Thalamus | 0.105 | 0.089 | 0.241 | 0.920 | -0.070 | 0.280 | 212 | 309 |
| Caudate | 0.063 | 0.089 | 0.481 | 0.920 | -0.112 | 0.238 | 212 | 308 |
| Putamen | 0.051 | 0.091 | 0.580 | 0.927 | -0.128 | 0.229 | 202 | 301 |
| Pallidum | 0.308 | 0.091 | 0.001 | 0.063 | 0.128 | 0.487 | 204 | 297 |
| Hippocampus | 0.058 | 0.089 | 0.519 | 0.920 | -0.117 | 0.233 | 212 | 309 |
| Amygdala | 0.089 | 0.089 | 0.318 | 0.920 | -0.085 | 0.263 | 211 | 316 |
| Accumbens | 0.021 | 0.089 | 0.817 | 0.933 | -0.154 | 0.196 | 210 | 312 |
| **Cortical thickness** | | | | | | | | |
| Banks superior temporal sulcus | 0.027 | 0.092 | 0.772 | 0.933 | -0.154 | 0.208 | 198 | 289 |
| Caudal anterior cingulate cortex | 0.002 | 0.089 | 0.981 | 0.994 | -0.172 | 0.176 | 214 | 315 |
| Caudal middle frontal gyrus | 0.064 | 0.089 | 0.474 | 0.920 | -0.110 | 0.238 | 210 | 318 |
| Cuneus | 0.070 | 0.090 | 0.439 | 0.920 | -0.106 | 0.245 | 206 | 315 |
| Entorhinal cortex | -0.056 | 0.094 | 0.553 | 0.927 | -0.239 | 0.128 | 187 | 294 |
| Fusiform gyrus | 0.000 | 0.089 | 0.996 | 0.996 | -0.173 | 0.174 | 213 | 315 |
| Inferior parietal cortex | 0.025 | 0.089 | 0.783 | 0.933 | -0.151 | 0.200 | 207 | 316 |
| Inferior temporal gyrus | -0.067 | 0.090 | 0.458 | 0.920 | -0.243 | 0.109 | 208 | 305 |
| Isthmus cingulate cortex | -0.065 | 0.089 | 0.463 | 0.920 | -0.240 | 0.109 | 211 | 314 |
| Lateral occipital cortex | 0.014 | 0.089 | 0.873 | 0.933 | -0.161 | 0.189 | 211 | 310 |
| Lateral orbitofrontal cortex | 0.086 | 0.089 | 0.337 | 0.920 | -0.089 | 0.260 | 212 | 311 |
| Lingual gyrus | 0.222 | 0.090 | 0.014 | 0.269 | 0.046 | 0.398 | 206 | 316 |
| Medial orbitofrontal gyrus | -0.051 | 0.089 | 0.566 | 0.927 | -0.225 | 0.123 | 213 | 312 |
| Middle temporal gyrus | -0.063 | 0.092 | 0.497 | 0.920 | -0.243 | 0.117 | 198 | 296 |
| Parahippocampal gyrus | 0.162 | 0.089 | 0.069 | 0.669 | -0.012 | 0.336 | 213 | 318 |
| Paracentral lobule | 0.020 | 0.089 | 0.825 | 0.933 | -0.154 | 0.194 | 211 | 317 |
| Pars opercularis | -0.077 | 0.089 | 0.387 | 0.920 | -0.251 | 0.097 | 213 | 317 |
| Pars orbitalis | 0.047 | 0.089 | 0.603 | 0.933 | -0.129 | 0.222 | 208 | 314 |
| Pars triangularis | 0.071 | 0.089 | 0.423 | 0.920 | -0.103 | 0.245 | 212 | 317 |
| Pericalcarine cortex | 0.077 | 0.090 | 0.393 | 0.920 | -0.099 | 0.253 | 206 | 313 |
| Postcentral gyrus | -0.014 | 0.089 | 0.872 | 0.933 | -0.188 | 0.160 | 213 | 315 |
| Posterior cingulate cortex | -0.071 | 0.089 | 0.428 | 0.920 | -0.245 | 0.104 | 210 | 315 |
| Precentral gyrus | 0.073 | 0.089 | 0.412 | 0.920 | -0.101 | 0.247 | 212 | 315 |
| Precuneus | 0.031 | 0.089 | 0.726 | 0.933 | -0.143 | 0.205 | 212 | 316 |
| Rostral anterior cingulate cortex | -0.037 | 0.089 | 0.676 | 0.933 | -0.211 | 0.136 | 214 | 316 |
| Rostral middle frontal gyrus | -0.018 | 0.089 | 0.841 | 0.933 | -0.193 | 0.157 | 209 | 315 |
| Superior frontal gyrus | 0.043 | 0.089 | 0.628 | 0.933 | -0.131 | 0.217 | 211 | 316 |
| Superior parietal gyrus | 0.043 | 0.089 | 0.626 | 0.933 | -0.130 | 0.217 | 214 | 315 |
| Superior temporal gyrus | 0.015 | 0.092 | 0.872 | 0.933 | -0.166 | 0.196 | 194 | 294 |
| Supramarginal gyrus | 0.022 | 0.090 | 0.807 | 0.933 | -0.154 | 0.198 | 206 | 315 |
| Frontal pole | 0.058 | 0.088 | 0.515 | 0.920 | -0.116 | 0.231 | 214 | 318 |
| Temporal pole | -0.023 | 0.089 | 0.798 | 0.933 | -0.197 | 0.151 | 212 | 316 |
| Transverse temporal gyrus | -0.028 | 0.089 | 0.752 | 0.933 | -0.202 | 0.146 | 214 | 316 |
| Insula | 0.058 | 0.089 | 0.516 | 0.920 | -0.117 | 0.233 | 210 | 316 |
| Mean Thickness | 0.040 | 0.089 | 0.653 | 0.933 | -0.134 | 0.213 | 214 | 316 |
| **Cortical surface area** | | | | | | | | |
| Banks superior temporal sulcus | -0.019 | 0.092 | 0.835 | 0.933 | -0.200 | 0.162 | 199 | 286 |
| Caudal anterior cingulate cortex | -0.007 | 0.089 | 0.938 | 0.976 | -0.181 | 0.167 | 214 | 315 |
| Caudal middle frontal gyrus | 0.109 | 0.089 | 0.221 | 0.920 | -0.065 | 0.283 | 213 | 316 |
| Cuneus | 0.093 | 0.090 | 0.304 | 0.920 | -0.083 | 0.268 | 206 | 314 |
| Entorhinal cortex | 0.235 | 0.094 | 0.013 | 0.269 | 0.051 | 0.419 | 189 | 292 |
| Fusiform gyrus | -0.005 | 0.089 | 0.952 | 0.977 | -0.179 | 0.169 | 212 | 316 |
| Inferior parietal cortex | -0.165 | 0.090 | 0.067 | 0.669 | -0.340 | 0.011 | 208 | 314 |
| Inferior temporal gyrus | 0.106 | 0.090 | 0.240 | 0.920 | -0.070 | 0.282 | 209 | 308 |
| Isthmus cingulate cortex | 0.229 | 0.089 | 0.010 | 0.269 | 0.054 | 0.404 | 212 | 315 |
| Lateral occipital cortex | 0.027 | 0.089 | 0.765 | 0.933 | -0.148 | 0.201 | 212 | 313 |
| Lateral orbitofrontal cortex | 0.086 | 0.089 | 0.339 | 0.920 | -0.089 | 0.260 | 212 | 311 |
| Lingual gyrus | 0.111 | 0.089 | 0.216 | 0.920 | -0.064 | 0.286 | 208 | 316 |
| Medial orbitofrontal gyrus | 0.188 | 0.089 | 0.035 | 0.504 | 0.014 | 0.363 | 213 | 312 |
| Middle temporal gyrus | -0.078 | 0.092 | 0.397 | 0.920 | -0.258 | 0.102 | 198 | 296 |
| Parahippocampal gyrus | -0.184 | 0.089 | 0.039 | 0.504 | -0.358 | -0.010 | 213 | 317 |
| Paracentral lobule | -0.011 | 0.089 | 0.901 | 0.949 | -0.185 | 0.162 | 213 | 318 |
| Pars opercularis | 0.022 | 0.089 | 0.805 | 0.933 | -0.152 | 0.196 | 211 | 317 |
| Pars orbitalis | -0.096 | 0.089 | 0.285 | 0.920 | -0.270 | 0.079 | 211 | 317 |
| Pars triangularis | 0.049 | 0.089 | 0.583 | 0.927 | -0.125 | 0.223 | 212 | 317 |
| Pericalcarine cortex | 0.159 | 0.090 | 0.078 | 0.675 | -0.017 | 0.335 | 206 | 314 |
| Postcentral gyrus | 0.062 | 0.089 | 0.487 | 0.920 | -0.112 | 0.236 | 213 | 315 |
| Posterior cingulate cortex | 0.102 | 0.089 | 0.252 | 0.920 | -0.072 | 0.277 | 210 | 317 |
| Precentral gyrus | 0.017 | 0.089 | 0.851 | 0.933 | -0.157 | 0.190 | 214 | 315 |
| Precuneus | 0.109 | 0.089 | 0.224 | 0.920 | -0.066 | 0.283 | 212 | 316 |
| Rostral anterior cingulate cortex | 0.052 | 0.089 | 0.561 | 0.927 | -0.122 | 0.226 | 213 | 315 |
| Rostral middle frontal gyrus | -0.068 | 0.089 | 0.446 | 0.920 | -0.243 | 0.107 | 210 | 315 |
| Superior frontal gyrus | 0.029 | 0.089 | 0.741 | 0.933 | -0.145 | 0.203 | 212 | 316 |
| Superior parietal gyrus | 0.017 | 0.089 | 0.848 | 0.933 | -0.156 | 0.191 | 214 | 316 |
| Superior temporal gyrus | -0.108 | 0.092 | 0.244 | 0.920 | -0.289 | 0.073 | 195 | 295 |
| Supramarginal gyrus | 0.101 | 0.090 | 0.263 | 0.920 | -0.075 | 0.277 | 206 | 312 |
| Frontal pole | -0.094 | 0.089 | 0.288 | 0.920 | -0.268 | 0.079 | 214 | 317 |
| Temporal pole | -0.060 | 0.089 | 0.497 | 0.920 | -0.234 | 0.113 | 213 | 317 |
| Transverse temporal gyrus | 0.103 | 0.088 | 0.245 | 0.920 | -0.070 | 0.277 | 214 | 318 |
| Insula | -0.060 | 0.089 | 0.500 | 0.920 | -0.235 | 0.114 | 209 | 316 |
| Full surface area | 0.039 | 0.088 | 0.660 | 0.933 | -0.134 | 0.213 | 214 | 317 |

Table S15. Differences in regional brain morphology between healthy controls and young people with a history of suicide attempt.

D: Cohen’s d effect size, SE: standard error; p: p-value, FDR-p: FDR corrected p-value, CI: confidence interval, HC: healthy controls, CC: clinical controls.

| **Region** | **D** | **SE** | **P** | **FDR-p** | **Lower CI** | **Upper CI** | **N attempt** | **N HC** |
| --- | --- | --- | --- | --- | --- | --- | --- | --- |
| **Subcortical volume** | | | | | | | | |
| Ventricle | 0.018 | 0.116 | 0.880 | 0.928 | -0.211 | 0.246 | 132 | 167 |
| Thalamus | -0.205 | 0.086 | 0.018 | 0.351 | -0.374 | -0.036 | 240 | 310 |
| Caudate | 0.023 | 0.085 | 0.788 | 0.904 | -0.144 | 0.191 | 242 | 315 |
| Putamen | -0.030 | 0.089 | 0.737 | 0.904 | -0.203 | 0.144 | 228 | 290 |
| Pallidum | -0.055 | 0.087 | 0.533 | 0.848 | -0.226 | 0.116 | 237 | 295 |
| Hippocampus | -0.087 | 0.085 | 0.309 | 0.585 | -0.254 | 0.080 | 247 | 316 |
| Amygdala | -0.057 | 0.086 | 0.507 | 0.841 | -0.225 | 0.111 | 242 | 314 |
| Accumbens | -0.025 | 0.085 | 0.772 | 0.904 | -0.191 | 0.142 | 240 | 326 |
| **Cortical thickness** | | | | | | | | |
| Banks superior temporal sulcus | -0.108 | 0.086 | 0.215 | 0.531 | -0.277 | 0.062 | 229 | 324 |
| Caudal anterior cingulate cortex | 0.029 | 0.084 | 0.731 | 0.904 | -0.136 | 0.194 | 247 | 331 |
| Caudal middle frontal gyrus | -0.212 | 0.084 | 0.012 | 0.351 | -0.377 | -0.046 | 248 | 331 |
| Cuneus | -0.011 | 0.085 | 0.896 | 0.928 | -0.178 | 0.155 | 244 | 322 |
| Entorhinal cortex | 0.047 | 0.086 | 0.589 | 0.901 | -0.121 | 0.215 | 232 | 330 |
| Fusiform gyrus | -0.196 | 0.084 | 0.021 | 0.351 | -0.361 | -0.031 | 247 | 333 |
| Inferior parietal cortex | -0.061 | 0.084 | 0.469 | 0.813 | -0.227 | 0.104 | 245 | 331 |
| Inferior temporal gyrus | -0.085 | 0.084 | 0.315 | 0.585 | -0.250 | 0.080 | 247 | 332 |
| Isthmus cingulate cortex | -0.018 | 0.084 | 0.827 | 0.910 | -0.183 | 0.146 | 248 | 332 |
| Lateral occipital cortex | -0.154 | 0.084 | 0.069 | 0.412 | -0.319 | 0.011 | 247 | 331 |
| Lateral orbitofrontal cortex | -0.113 | 0.085 | 0.185 | 0.515 | -0.278 | 0.053 | 245 | 327 |
| Lingual gyrus | -0.174 | 0.084 | 0.039 | 0.412 | -0.339 | -0.009 | 249 | 330 |
| Medial orbitofrontal gyrus | -0.059 | 0.085 | 0.490 | 0.831 | -0.225 | 0.107 | 244 | 326 |
| Middle temporal gyrus | -0.203 | 0.086 | 0.019 | 0.351 | -0.372 | -0.034 | 232 | 325 |
| Parahippocampal gyrus | 0.122 | 0.084 | 0.149 | 0.484 | -0.043 | 0.287 | 246 | 333 |
| Paracentral lobule | -0.112 | 0.084 | 0.184 | 0.515 | -0.277 | 0.053 | 249 | 331 |
| Pars opercularis | -0.145 | 0.084 | 0.087 | 0.452 | -0.310 | 0.020 | 247 | 331 |
| Pars orbitalis | 0.030 | 0.084 | 0.726 | 0.904 | -0.135 | 0.194 | 247 | 331 |
| Pars triangularis | -0.163 | 0.084 | 0.053 | 0.412 | -0.328 | 0.002 | 249 | 331 |
| Pericalcarine cortex | -0.033 | 0.085 | 0.696 | 0.904 | -0.199 | 0.133 | 247 | 322 |
| Postcentral gyrus | -0.131 | 0.084 | 0.121 | 0.482 | -0.296 | 0.034 | 248 | 331 |
| Posterior cingulate cortex | -0.053 | 0.084 | 0.531 | 0.848 | -0.217 | 0.112 | 249 | 331 |
| Precentral gyrus | -0.123 | 0.084 | 0.145 | 0.484 | -0.288 | 0.042 | 247 | 332 |
| Precuneus | -0.137 | 0.084 | 0.105 | 0.464 | -0.301 | 0.028 | 249 | 332 |
| Rostral anterior cingulate cortex | -0.108 | 0.084 | 0.200 | 0.520 | -0.273 | 0.057 | 246 | 332 |
| Rostral middle frontal gyrus | -0.157 | 0.084 | 0.064 | 0.412 | -0.322 | 0.008 | 248 | 326 |
| Superior frontal gyrus | -0.088 | 0.084 | 0.300 | 0.585 | -0.253 | 0.077 | 248 | 327 |
| Superior parietal gyrus | -0.151 | 0.084 | 0.074 | 0.412 | -0.316 | 0.014 | 248 | 331 |
| Superior temporal gyrus | -0.143 | 0.086 | 0.098 | 0.464 | -0.311 | 0.026 | 232 | 326 |
| Supramarginal gyrus | -0.116 | 0.084 | 0.172 | 0.515 | -0.281 | 0.049 | 246 | 331 |
| Frontal pole | 0.037 | 0.084 | 0.664 | 0.904 | -0.128 | 0.202 | 247 | 331 |
| Temporal pole | 0.040 | 0.084 | 0.638 | 0.904 | -0.125 | 0.204 | 248 | 332 |
| Transverse temporal gyrus | -0.039 | 0.084 | 0.641 | 0.904 | -0.204 | 0.125 | 248 | 333 |
| Insula | -0.037 | 0.084 | 0.659 | 0.904 | -0.202 | 0.127 | 248 | 329 |
| Mean Thickness | -0.155 | 0.084 | 0.066 | 0.412 | -0.320 | 0.009 | 249 | 333 |
| **Cortical surface area** | | | | | | | | |
| Banks superior temporal sulcus | -0.196 | 0.087 | 0.026 | 0.351 | -0.367 | -0.025 | 226 | 316 |
| Caudal anterior cingulate cortex | -0.137 | 0.084 | 0.107 | 0.464 | -0.302 | 0.028 | 246 | 330 |
| Caudal middle frontal gyrus | -0.097 | 0.084 | 0.253 | 0.546 | -0.262 | 0.068 | 248 | 331 |
| Cuneus | 0.030 | 0.085 | 0.724 | 0.904 | -0.137 | 0.197 | 243 | 320 |
| Entorhinal cortex | -0.010 | 0.086 | 0.910 | 0.928 | -0.178 | 0.158 | 230 | 331 |
| Fusiform gyrus | -0.127 | 0.084 | 0.136 | 0.482 | -0.292 | 0.038 | 246 | 332 |
| Inferior parietal cortex | -0.188 | 0.084 | 0.027 | 0.351 | -0.353 | -0.023 | 247 | 329 |
| Inferior temporal gyrus | -0.165 | 0.084 | 0.053 | 0.412 | -0.330 | 0.001 | 246 | 329 |
| Isthmus cingulate cortex | 0.021 | 0.084 | 0.807 | 0.910 | -0.144 | 0.185 | 248 | 332 |
| Lateral occipital cortex | 0.017 | 0.084 | 0.840 | 0.910 | -0.148 | 0.182 | 246 | 332 |
| Lateral orbitofrontal cortex | -0.096 | 0.085 | 0.259 | 0.546 | -0.262 | 0.069 | 245 | 327 |
| Lingual gyrus | -0.028 | 0.084 | 0.745 | 0.904 | -0.192 | 0.137 | 247 | 331 |
| Medial orbitofrontal gyrus | -0.044 | 0.085 | 0.606 | 0.904 | -0.210 | 0.122 | 245 | 327 |
| Middle temporal gyrus | -0.096 | 0.086 | 0.269 | 0.552 | -0.265 | 0.073 | 232 | 323 |
| Parahippocampal gyrus | -0.026 | 0.084 | 0.759 | 0.904 | -0.191 | 0.139 | 247 | 331 |
| Paracentral lobule | 0.034 | 0.084 | 0.684 | 0.904 | -0.130 | 0.199 | 249 | 330 |
| Pars opercularis | -0.018 | 0.084 | 0.835 | 0.910 | -0.182 | 0.147 | 248 | 329 |
| Pars orbitalis | 0.031 | 0.084 | 0.719 | 0.904 | -0.134 | 0.195 | 247 | 331 |
| Pars triangularis | 0.009 | 0.084 | 0.916 | 0.928 | -0.156 | 0.173 | 248 | 332 |
| Pericalcarine cortex | 0.049 | 0.085 | 0.568 | 0.886 | -0.117 | 0.215 | 245 | 322 |
| Postcentral gyrus | -0.005 | 0.084 | 0.957 | 0.957 | -0.169 | 0.160 | 248 | 330 |
| Posterior cingulate cortex | -0.096 | 0.084 | 0.258 | 0.546 | -0.260 | 0.069 | 249 | 332 |
| Precentral gyrus | -0.105 | 0.084 | 0.218 | 0.531 | -0.270 | 0.060 | 246 | 330 |
| Precuneus | -0.023 | 0.084 | 0.782 | 0.904 | -0.188 | 0.141 | 249 | 332 |
| Rostral anterior cingulate cortex | -0.128 | 0.085 | 0.132 | 0.482 | -0.294 | 0.037 | 244 | 330 |
| Rostral middle frontal gyrus | -0.062 | 0.084 | 0.469 | 0.813 | -0.227 | 0.104 | 248 | 326 |
| Superior frontal gyrus | -0.097 | 0.084 | 0.252 | 0.546 | -0.263 | 0.068 | 248 | 326 |
| Superior parietal gyrus | 0.010 | 0.084 | 0.910 | 0.928 | -0.155 | 0.174 | 248 | 331 |
| Superior temporal gyrus | -0.131 | 0.086 | 0.134 | 0.482 | -0.300 | 0.039 | 231 | 321 |
| Supramarginal gyrus | -0.065 | 0.084 | 0.446 | 0.809 | -0.230 | 0.100 | 247 | 328 |
| Frontal pole | -0.155 | 0.084 | 0.068 | 0.412 | -0.320 | 0.010 | 248 | 330 |
| Temporal pole | 0.118 | 0.084 | 0.162 | 0.505 | -0.046 | 0.283 | 248 | 331 |
| Transverse temporal gyrus | -0.096 | 0.084 | 0.258 | 0.546 | -0.260 | 0.069 | 249 | 333 |
| Insula | -0.091 | 0.085 | 0.284 | 0.568 | -0.257 | 0.074 | 245 | 327 |
| Full surface area | -0.109 | 0.084 | 0.198 | 0.520 | -0.273 | 0.056 | 249 | 332 |

Table S16. Differences in regional brain morphology between clinical controls and young people with a history of suicide attempt.

D: Cohen’s d effect size, SE: standard error; p: p-value, FDR-p: FDR corrected p-value, CI: confidence interval, HC: healthy controls, CC: clinical controls.

| **Region** | **D** | **SE** | **P** | **FDR-p** | **Lower CI** | **Upper CI** | **N attempt** | **N CC** |
| --- | --- | --- | --- | --- | --- | --- | --- | --- |
| **Subcortical volume** | | | | | | | | |
| Ventricle | 0.010 | 0.102 | 0.925 | 0.961 | -0.190 | 0.210 | 132 | 352 |
| Thalamus | -0.099 | 0.074 | 0.186 | 0.764 | -0.244 | 0.047 | 240 | 738 |
| Caudate | 0.017 | 0.074 | 0.823 | 0.961 | -0.129 | 0.162 | 242 | 736 |
| Putamen | 0.005 | 0.076 | 0.946 | 0.961 | -0.144 | 0.154 | 228 | 721 |
| Pallidum | -0.130 | 0.075 | 0.085 | 0.741 | -0.276 | 0.017 | 237 | 718 |
| Hippocampus | -0.022 | 0.074 | 0.770 | 0.961 | -0.166 | 0.123 | 247 | 734 |
| Amygdala | -0.009 | 0.074 | 0.901 | 0.961 | -0.154 | 0.136 | 242 | 748 |
| Accumbens | -0.013 | 0.074 | 0.866 | 0.961 | -0.158 | 0.133 | 240 | 741 |
| **Cortical thickness** | | | | | | | | |
| Banks superior temporal sulcus | -0.112 | 0.076 | 0.144 | 0.741 | -0.261 | 0.038 | 229 | 695 |
| Caudal anterior cingulate cortex | 0.066 | 0.073 | 0.369 | 0.849 | -0.078 | 0.210 | 247 | 753 |
| Caudal middle frontal gyrus | 0.022 | 0.073 | 0.768 | 0.961 | -0.122 | 0.165 | 248 | 753 |
| Cuneus | -0.013 | 0.074 | 0.860 | 0.961 | -0.158 | 0.132 | 244 | 747 |
| Entorhinal cortex | 0.069 | 0.075 | 0.364 | 0.849 | -0.079 | 0.217 | 232 | 722 |
| Fusiform gyrus | -0.105 | 0.073 | 0.152 | 0.741 | -0.249 | 0.038 | 247 | 752 |
| Inferior parietal cortex | 0.117 | 0.074 | 0.112 | 0.741 | -0.027 | 0.262 | 245 | 753 |
| Inferior temporal gyrus | -0.041 | 0.073 | 0.578 | 0.961 | -0.185 | 0.103 | 247 | 752 |
| Isthmus cingulate cortex | 0.098 | 0.073 | 0.182 | 0.764 | -0.046 | 0.242 | 248 | 754 |
| Lateral occipital cortex | -0.004 | 0.073 | 0.957 | 0.961 | -0.148 | 0.140 | 247 | 750 |
| Lateral orbitofrontal cortex | -0.045 | 0.074 | 0.546 | 0.961 | -0.189 | 0.100 | 245 | 747 |
| Lingual gyrus | -0.087 | 0.073 | 0.236 | 0.787 | -0.230 | 0.057 | 249 | 753 |
| Medial orbitofrontal gyrus | -0.017 | 0.074 | 0.822 | 0.961 | -0.161 | 0.128 | 244 | 747 |
| Middle temporal gyrus | -0.114 | 0.076 | 0.132 | 0.741 | -0.263 | 0.034 | 232 | 708 |
| Parahippocampal gyrus | 0.082 | 0.073 | 0.264 | 0.824 | -0.062 | 0.226 | 246 | 753 |
| Paracentral lobule | 0.044 | 0.073 | 0.546 | 0.961 | -0.099 | 0.188 | 249 | 754 |
| Pars opercularis | -0.035 | 0.073 | 0.632 | 0.961 | -0.179 | 0.108 | 247 | 755 |
| Pars orbitalis | 0.068 | 0.073 | 0.359 | 0.849 | -0.076 | 0.211 | 247 | 750 |
| Pars triangularis | -0.037 | 0.073 | 0.609 | 0.961 | -0.181 | 0.106 | 249 | 755 |
| Pericalcarine cortex | -0.095 | 0.073 | 0.196 | 0.764 | -0.239 | 0.049 | 247 | 753 |
| Postcentral gyrus | -0.009 | 0.073 | 0.898 | 0.961 | -0.153 | 0.134 | 248 | 753 |
| Posterior cingulate cortex | 0.029 | 0.073 | 0.690 | 0.961 | -0.114 | 0.173 | 249 | 753 |
| Precentral gyrus | 0.004 | 0.073 | 0.961 | 0.961 | -0.140 | 0.147 | 247 | 750 |
| Precuneus | -0.033 | 0.073 | 0.648 | 0.961 | -0.177 | 0.110 | 249 | 757 |
| Rostral anterior cingulate cortex | 0.037 | 0.073 | 0.619 | 0.961 | -0.107 | 0.180 | 246 | 758 |
| Rostral middle frontal gyrus | 0.004 | 0.073 | 0.957 | 0.961 | -0.140 | 0.148 | 248 | 748 |
| Superior frontal gyrus | 0.067 | 0.073 | 0.363 | 0.849 | -0.077 | 0.210 | 248 | 753 |
| Superior parietal gyrus | 0.013 | 0.073 | 0.858 | 0.961 | -0.130 | 0.157 | 248 | 754 |
| Superior temporal gyrus | -0.067 | 0.076 | 0.379 | 0.849 | -0.215 | 0.081 | 232 | 716 |
| Supramarginal gyrus | -0.035 | 0.073 | 0.633 | 0.961 | -0.179 | 0.109 | 246 | 753 |
| Frontal pole | -0.009 | 0.073 | 0.907 | 0.961 | -0.152 | 0.135 | 247 | 754 |
| Temporal pole | -0.008 | 0.073 | 0.908 | 0.961 | -0.152 | 0.135 | 248 | 755 |
| Transverse temporal gyrus | -0.056 | 0.073 | 0.442 | 0.932 | -0.200 | 0.087 | 248 | 755 |
| Insula | -0.025 | 0.073 | 0.735 | 0.961 | -0.168 | 0.119 | 248 | 756 |
| Mean Thickness | -0.007 | 0.073 | 0.924 | 0.961 | -0.150 | 0.136 | 249 | 756 |
| **Cortical surface area** | | | | | | | | |
| Banks superior temporal sulcus | -0.145 | 0.077 | 0.060 | 0.741 | -0.296 | 0.005 | 226 | 688 |
| Caudal anterior cingulate cortex | -0.208 | 0.074 | 0.005 | 0.390 | -0.352 | -0.064 | 246 | 752 |
| Caudal middle frontal gyrus | -0.077 | 0.073 | 0.298 | 0.849 | -0.220 | 0.067 | 248 | 754 |
| Cuneus | 0.050 | 0.074 | 0.499 | 0.961 | -0.095 | 0.195 | 243 | 745 |
| Entorhinal cortex | -0.014 | 0.076 | 0.854 | 0.961 | -0.162 | 0.134 | 230 | 720 |
| Fusiform gyrus | -0.141 | 0.073 | 0.056 | 0.741 | -0.285 | 0.003 | 246 | 754 |
| Inferior parietal cortex | -0.173 | 0.073 | 0.019 | 0.741 | -0.317 | -0.030 | 247 | 751 |
| Inferior temporal gyrus | -0.133 | 0.074 | 0.072 | 0.741 | -0.277 | 0.011 | 246 | 752 |
| Isthmus cingulate cortex | 0.019 | 0.073 | 0.799 | 0.961 | -0.125 | 0.162 | 248 | 747 |
| Lateral occipital cortex | 0.054 | 0.074 | 0.468 | 0.961 | -0.091 | 0.198 | 246 | 746 |
| Lateral orbitofrontal cortex | -0.124 | 0.074 | 0.095 | 0.741 | -0.268 | 0.021 | 245 | 748 |
| Lingual gyrus | 0.010 | 0.073 | 0.896 | 0.961 | -0.134 | 0.153 | 247 | 755 |
| Medial orbitofrontal gyrus | -0.058 | 0.074 | 0.436 | 0.932 | -0.202 | 0.087 | 245 | 748 |
| Middle temporal gyrus | -0.018 | 0.076 | 0.814 | 0.961 | -0.166 | 0.131 | 232 | 704 |
| Parahippocampal gyrus | 0.031 | 0.073 | 0.674 | 0.961 | -0.113 | 0.175 | 247 | 748 |
| Paracentral lobule | -0.109 | 0.073 | 0.136 | 0.741 | -0.253 | 0.034 | 249 | 750 |
| Pars opercularis | -0.026 | 0.073 | 0.720 | 0.961 | -0.170 | 0.117 | 248 | 750 |
| Pars orbitalis | -0.033 | 0.073 | 0.652 | 0.961 | -0.177 | 0.111 | 247 | 752 |
| Pars triangularis | 0.044 | 0.073 | 0.548 | 0.961 | -0.099 | 0.188 | 248 | 755 |
| Pericalcarine cortex | 0.065 | 0.074 | 0.381 | 0.849 | -0.080 | 0.209 | 245 | 752 |
| Postcentral gyrus | 0.009 | 0.073 | 0.901 | 0.961 | -0.134 | 0.153 | 248 | 753 |
| Posterior cingulate cortex | -0.125 | 0.073 | 0.088 | 0.741 | -0.269 | 0.018 | 249 | 750 |
| Precentral gyrus | -0.086 | 0.073 | 0.242 | 0.787 | -0.230 | 0.058 | 246 | 751 |
| Precuneus | -0.024 | 0.073 | 0.739 | 0.961 | -0.168 | 0.119 | 249 | 751 |
| Rostral anterior cingulate cortex | -0.136 | 0.074 | 0.066 | 0.741 | -0.280 | 0.008 | 244 | 755 |
| Rostral middle frontal gyrus | -0.079 | 0.073 | 0.281 | 0.843 | -0.223 | 0.064 | 248 | 751 |
| Superior frontal gyrus | -0.088 | 0.073 | 0.233 | 0.787 | -0.232 | 0.056 | 248 | 750 |
| Superior parietal gyrus | -0.069 | 0.073 | 0.349 | 0.849 | -0.212 | 0.074 | 248 | 758 |
| Superior temporal gyrus | -0.106 | 0.076 | 0.164 | 0.752 | -0.254 | 0.042 | 231 | 714 |
| Supramarginal gyrus | -0.065 | 0.073 | 0.378 | 0.849 | -0.209 | 0.079 | 247 | 749 |
| Frontal pole | -0.158 | 0.073 | 0.031 | 0.741 | -0.302 | -0.015 | 248 | 752 |
| Temporal pole | 0.012 | 0.073 | 0.871 | 0.961 | -0.131 | 0.155 | 248 | 757 |
| Transverse temporal gyrus | -0.090 | 0.073 | 0.219 | 0.787 | -0.234 | 0.053 | 249 | 754 |
| Insula | -0.047 | 0.074 | 0.528 | 0.961 | -0.191 | 0.098 | 245 | 747 |
| Full surface area | -0.110 | 0.073 | 0.133 | 0.741 | -0.254 | 0.033 | 249 | 756 |

Table 17. Differences in regional brain morphology between clinical controls and young people with a history of suicide attempt, additionally corrected for type of diagnosis. D: Cohen’s d effect size, SE: standard error; p: p-value, FDR-p: FDR corrected p-value, CI: confidence interval, HC: healthy controls, CC: clinical controls.

| **Region** | **D** | **SE** | **P** | **FDR-p** | **Lower CI** | **Upper CI** | **N attempt** | **N CC** |
| --- | --- | --- | --- | --- | --- | --- | --- | --- |
| **Subcortical volume** | | | | | | | | |
| Ventricle | -0.037 | 0.109 | 0.739 | 0.908 | -0.250 | 0.176 | 120 | 287 |
| Thalamus | -0.124 | 0.079 | 0.122 | 0.574 | -0.279 | 0.032 | 220 | 581 |
| Caudate | 0.032 | 0.079 | 0.686 | 0.900 | -0.123 | 0.187 | 221 | 581 |
| Putamen | 0.038 | 0.081 | 0.639 | 0.900 | -0.120 | 0.197 | 209 | 564 |
| Pallidum | -0.121 | 0.080 | 0.134 | 0.574 | -0.277 | 0.036 | 217 | 564 |
| Hippocampus | -0.008 | 0.078 | 0.922 | 0.946 | -0.161 | 0.146 | 226 | 580 |
| Amygdala | -0.012 | 0.079 | 0.884 | 0.946 | -0.166 | 0.143 | 221 | 589 |
| Accumbens | -0.009 | 0.079 | 0.907 | 0.946 | -0.165 | 0.146 | 219 | 583 |
| **Cortical thickness** | | | | | | | | |
| Banks superior temporal sulcus | -0.114 | 0.081 | 0.162 | 0.574 | -0.272 | 0.045 | 212 | 563 |
| Caudal anterior cingulate cortex | 0.046 | 0.078 | 0.557 | 0.900 | -0.107 | 0.199 | 226 | 596 |
| Caudal middle frontal gyrus | -0.007 | 0.078 | 0.934 | 0.946 | -0.159 | 0.146 | 227 | 595 |
| Cuneus | -0.031 | 0.079 | 0.694 | 0.900 | -0.185 | 0.123 | 223 | 588 |
| Entorhinal cortex | 0.101 | 0.081 | 0.212 | 0.660 | -0.057 | 0.259 | 212 | 566 |
| Fusiform gyrus | -0.119 | 0.078 | 0.130 | 0.574 | -0.273 | 0.034 | 226 | 593 |
| Inferior parietal cortex | 0.087 | 0.078 | 0.272 | 0.786 | -0.067 | 0.240 | 224 | 594 |
| Inferior temporal gyrus | -0.057 | 0.078 | 0.467 | 0.900 | -0.210 | 0.096 | 226 | 596 |
| Isthmus cingulate cortex | 0.111 | 0.078 | 0.159 | 0.574 | -0.042 | 0.264 | 227 | 596 |
| Lateral occipital cortex | -0.050 | 0.078 | 0.524 | 0.900 | -0.203 | 0.103 | 226 | 593 |
| Lateral orbitofrontal cortex | -0.062 | 0.079 | 0.432 | 0.900 | -0.216 | 0.092 | 224 | 589 |
| Lingual gyrus | -0.111 | 0.078 | 0.156 | 0.574 | -0.264 | 0.042 | 228 | 595 |
| Medial orbitofrontal gyrus | -0.032 | 0.079 | 0.682 | 0.900 | -0.187 | 0.122 | 223 | 588 |
| Middle temporal gyrus | -0.129 | 0.080 | 0.111 | 0.574 | -0.286 | 0.028 | 214 | 573 |
| Parahippocampal gyrus | 0.078 | 0.078 | 0.323 | 0.869 | -0.075 | 0.231 | 226 | 594 |
| Paracentral lobule | 0.029 | 0.078 | 0.714 | 0.900 | -0.124 | 0.181 | 228 | 595 |
| Pars opercularis | -0.047 | 0.078 | 0.552 | 0.900 | -0.200 | 0.106 | 227 | 596 |
| Pars orbitalis | 0.020 | 0.078 | 0.804 | 0.908 | -0.134 | 0.173 | 226 | 592 |
| Pars triangularis | -0.051 | 0.078 | 0.512 | 0.900 | -0.204 | 0.101 | 228 | 596 |
| Pericalcarine cortex | -0.126 | 0.078 | 0.110 | 0.574 | -0.279 | 0.028 | 226 | 594 |
| Postcentral gyrus | -0.041 | 0.078 | 0.603 | 0.900 | -0.194 | 0.112 | 227 | 594 |
| Posterior cingulate cortex | 0.044 | 0.078 | 0.578 | 0.900 | -0.109 | 0.196 | 228 | 596 |
| Precentral gyrus | -0.012 | 0.078 | 0.882 | 0.946 | -0.165 | 0.142 | 226 | 593 |
| Precuneus | -0.061 | 0.078 | 0.438 | 0.900 | -0.213 | 0.092 | 228 | 598 |
| Rostral anterior cingulate cortex | 0.029 | 0.078 | 0.714 | 0.900 | -0.124 | 0.182 | 225 | 599 |
| Rostral middle frontal gyrus | -0.035 | 0.078 | 0.653 | 0.900 | -0.188 | 0.118 | 227 | 590 |
| Superior frontal gyrus | 0.058 | 0.078 | 0.462 | 0.900 | -0.095 | 0.211 | 227 | 595 |
| Superior parietal gyrus | -0.010 | 0.078 | 0.902 | 0.946 | -0.163 | 0.143 | 227 | 595 |
| Superior temporal gyrus | -0.138 | 0.080 | 0.087 | 0.574 | -0.296 | 0.019 | 213 | 571 |
| Supramarginal gyrus | -0.075 | 0.078 | 0.340 | 0.869 | -0.229 | 0.078 | 225 | 595 |
| Frontal pole | -0.029 | 0.078 | 0.708 | 0.900 | -0.183 | 0.124 | 226 | 595 |
| Temporal pole | -0.029 | 0.078 | 0.715 | 0.900 | -0.181 | 0.124 | 227 | 597 |
| Transverse temporal gyrus | -0.050 | 0.078 | 0.523 | 0.900 | -0.203 | 0.103 | 227 | 596 |
| Insula | -0.033 | 0.078 | 0.672 | 0.900 | -0.186 | 0.120 | 227 | 597 |
| Mean Thickness | -0.042 | 0.078 | 0.594 | 0.900 | -0.194 | 0.111 | 228 | 597 |
| **Cortical surface area** | | | | | | | | |
| Banks superior temporal sulcus | -0.186 | 0.081 | 0.024 | 0.574 | -0.345 | -0.026 | 208 | 556 |
| Caudal anterior cingulate cortex | -0.182 | 0.078 | 0.021 | 0.574 | -0.336 | -0.028 | 225 | 595 |
| Caudal middle frontal gyrus | -0.056 | 0.078 | 0.474 | 0.900 | -0.209 | 0.097 | 227 | 596 |
| Cuneus | 0.053 | 0.079 | 0.509 | 0.900 | -0.102 | 0.207 | 222 | 586 |
| Entorhinal cortex | -0.016 | 0.081 | 0.849 | 0.946 | -0.174 | 0.143 | 210 | 562 |
| Fusiform gyrus | -0.154 | 0.078 | 0.052 | 0.574 | -0.307 | 0.000 | 225 | 596 |
| Inferior parietal cortex | -0.168 | 0.078 | 0.034 | 0.574 | -0.321 | -0.014 | 226 | 593 |
| Inferior temporal gyrus | -0.129 | 0.078 | 0.102 | 0.574 | -0.283 | 0.024 | 225 | 595 |
| Isthmus cingulate cortex | 0.024 | 0.078 | 0.757 | 0.908 | -0.129 | 0.177 | 227 | 590 |
| Lateral occipital cortex | 0.040 | 0.078 | 0.611 | 0.900 | -0.113 | 0.194 | 225 | 590 |
| Lateral orbitofrontal cortex | -0.134 | 0.079 | 0.091 | 0.574 | -0.288 | 0.020 | 224 | 592 |
| Lingual gyrus | 0.022 | 0.078 | 0.779 | 0.908 | -0.131 | 0.175 | 226 | 596 |
| Medial orbitofrontal gyrus | -0.039 | 0.079 | 0.619 | 0.900 | -0.193 | 0.115 | 224 | 589 |
| Middle temporal gyrus | -0.026 | 0.080 | 0.748 | 0.908 | -0.183 | 0.131 | 214 | 570 |
| Parahippocampal gyrus | 0.062 | 0.078 | 0.430 | 0.900 | -0.091 | 0.215 | 227 | 589 |
| Paracentral lobule | -0.119 | 0.078 | 0.131 | 0.574 | -0.272 | 0.034 | 228 | 594 |
| Pars opercularis | -0.020 | 0.078 | 0.801 | 0.908 | -0.173 | 0.133 | 228 | 593 |
| Pars orbitalis | -0.048 | 0.078 | 0.546 | 0.900 | -0.201 | 0.106 | 226 | 594 |
| Pars triangularis | 0.021 | 0.078 | 0.786 | 0.908 | -0.131 | 0.174 | 227 | 596 |
| Pericalcarine cortex | 0.080 | 0.078 | 0.312 | 0.868 | -0.074 | 0.234 | 224 | 593 |
| Postcentral gyrus | 0.000 | 0.078 | 0.998 | 0.998 | -0.153 | 0.153 | 227 | 595 |
| Posterior cingulate cortex | -0.119 | 0.078 | 0.130 | 0.574 | -0.272 | 0.034 | 228 | 593 |
| Precentral gyrus | -0.099 | 0.078 | 0.209 | 0.660 | -0.253 | 0.054 | 225 | 593 |
| Precuneus | -0.036 | 0.078 | 0.650 | 0.900 | -0.188 | 0.117 | 228 | 594 |
| Rostral anterior cingulate cortex | -0.111 | 0.079 | 0.161 | 0.574 | -0.265 | 0.043 | 223 | 598 |
| Rostral middle frontal gyrus | -0.120 | 0.078 | 0.129 | 0.574 | -0.273 | 0.034 | 227 | 594 |
| Superior frontal gyrus | -0.122 | 0.078 | 0.123 | 0.574 | -0.275 | 0.031 | 227 | 595 |
| Superior parietal gyrus | -0.069 | 0.078 | 0.379 | 0.900 | -0.222 | 0.084 | 227 | 599 |
| Superior temporal gyrus | -0.109 | 0.081 | 0.182 | 0.616 | -0.266 | 0.049 | 212 | 568 |
| Supramarginal gyrus | -0.059 | 0.078 | 0.458 | 0.900 | -0.212 | 0.095 | 226 | 591 |
| Frontal pole | -0.159 | 0.078 | 0.044 | 0.574 | -0.312 | -0.006 | 227 | 595 |
| Temporal pole | -0.008 | 0.078 | 0.923 | 0.946 | -0.160 | 0.145 | 227 | 598 |
| Transverse temporal gyrus | -0.091 | 0.078 | 0.245 | 0.736 | -0.244 | 0.061 | 228 | 595 |
| Insula | -0.075 | 0.078 | 0.345 | 0.869 | -0.228 | 0.079 | 224 | 592 |
| Full surface area | -0.115 | 0.078 | 0.143 | 0.574 | -0.268 | 0.038 | 228 | 598 |

Table S18. Differences in regional brain morphology between clinical controls and young people with a lifetime history of suicide attempt **in men only**. D: Cohen’s d effect size, SE: standard error; p: p-value, FDR-p: FDR corrected p-value, CI: confidence interval, HC: healthy controls, CC: clinical controls.

| **Region** | **D** | **SE** | **P** | **FDR-p** | **Lower CI** | **Upper CI** | **N attempt** | **N CC** |
| --- | --- | --- | --- | --- | --- | --- | --- | --- |
| **Subcortical volume** | | | | | | | | |
| Ventricle | -0.124 | 0.280 | 0.670 | 0.941 | -0.673 | 0.426 | 17 | 51 |
| Thalamus | -0.088 | 0.175 | 0.621 | 0.941 | -0.430 | 0.255 | 41 | 165 |
| Caudate | 0.409 | 0.176 | 0.022 | 0.941 | 0.065 | 0.754 | 41 | 164 |
| Putamen | -0.044 | 0.179 | 0.809 | 0.941 | -0.395 | 0.307 | 39 | 157 |
| Pallidum | -0.079 | 0.175 | 0.655 | 0.941 | -0.423 | 0.264 | 41 | 160 |
| Hippocampus | 0.067 | 0.173 | 0.700 | 0.941 | -0.272 | 0.406 | 42 | 164 |
| Amygdala | 0.131 | 0.173 | 0.455 | 0.941 | -0.208 | 0.470 | 42 | 165 |
| Accumbens | 0.076 | 0.177 | 0.670 | 0.941 | -0.270 | 0.423 | 40 | 162 |
| **Cortical thickness** | | | | | | | | |
| Banks superior temporal sulcus | 0.100 | 0.181 | 0.585 | 0.941 | -0.256 | 0.455 | 38 | 153 |
| Caudal anterior cingulate cortex | 0.076 | 0.173 | 0.663 | 0.941 | -0.262 | 0.414 | 42 | 167 |
| Caudal middle frontal gyrus | -0.200 | 0.173 | 0.251 | 0.941 | -0.539 | 0.139 | 42 | 166 |
| Cuneus | 0.021 | 0.175 | 0.905 | 0.941 | -0.321 | 0.363 | 41 | 164 |
| Entorhinal cortex | 0.132 | 0.175 | 0.456 | 0.941 | -0.212 | 0.476 | 41 | 158 |
| Fusiform gyrus | -0.042 | 0.171 | 0.810 | 0.941 | -0.378 | 0.294 | 43 | 163 |
| Inferior parietal cortex | 0.165 | 0.173 | 0.344 | 0.941 | -0.174 | 0.504 | 42 | 167 |
| Inferior temporal gyrus | -0.120 | 0.171 | 0.486 | 0.941 | -0.456 | 0.215 | 43 | 167 |
| Isthmus cingulate cortex | 0.102 | 0.171 | 0.554 | 0.941 | -0.233 | 0.438 | 43 | 165 |
| Lateral occipital cortex | 0.043 | 0.171 | 0.802 | 0.941 | -0.292 | 0.379 | 43 | 166 |
| Lateral orbitofrontal cortex | 0.045 | 0.176 | 0.800 | 0.941 | -0.300 | 0.391 | 40 | 165 |
| Lingual gyrus | -0.052 | 0.171 | 0.761 | 0.941 | -0.388 | 0.283 | 43 | 167 |
| Medial orbitofrontal gyrus | -0.082 | 0.175 | 0.643 | 0.941 | -0.424 | 0.261 | 41 | 163 |
| Middle temporal gyrus | -0.046 | 0.177 | 0.795 | 0.941 | -0.394 | 0.301 | 40 | 157 |
| Parahippocampal gyrus | 0.191 | 0.172 | 0.271 | 0.941 | -0.146 | 0.527 | 43 | 164 |
| Paracentral lobule | 0.101 | 0.171 | 0.560 | 0.941 | -0.235 | 0.436 | 43 | 166 |
| Pars opercularis | -0.011 | 0.171 | 0.949 | 0.974 | -0.346 | 0.324 | 43 | 166 |
| Pars orbitalis | 0.131 | 0.173 | 0.454 | 0.941 | -0.208 | 0.470 | 42 | 165 |
| Pars triangularis | 0.040 | 0.171 | 0.815 | 0.941 | -0.295 | 0.376 | 43 | 166 |
| Pericalcarine cortex | -0.202 | 0.173 | 0.246 | 0.941 | -0.541 | 0.136 | 42 | 167 |
| Postcentral gyrus | -0.047 | 0.171 | 0.787 | 0.941 | -0.382 | 0.289 | 43 | 165 |
| Posterior cingulate cortex | 0.130 | 0.171 | 0.450 | 0.941 | -0.205 | 0.466 | 43 | 166 |
| Precentral gyrus | 0.094 | 0.171 | 0.587 | 0.941 | -0.242 | 0.429 | 43 | 166 |
| Precuneus | 0.094 | 0.171 | 0.588 | 0.941 | -0.242 | 0.429 | 43 | 166 |
| Rostral anterior cingulate cortex | 0.103 | 0.171 | 0.552 | 0.941 | -0.233 | 0.438 | 43 | 167 |
| Rostral middle frontal gyrus | -0.132 | 0.171 | 0.444 | 0.941 | -0.468 | 0.204 | 43 | 164 |
| Superior frontal gyrus | 0.072 | 0.171 | 0.676 | 0.941 | -0.264 | 0.408 | 43 | 165 |
| Superior parietal gyrus | 0.026 | 0.171 | 0.882 | 0.941 | -0.310 | 0.361 | 43 | 167 |
| Superior temporal gyrus | 0.115 | 0.179 | 0.524 | 0.941 | -0.236 | 0.466 | 39 | 157 |
| Supramarginal gyrus | 0.109 | 0.173 | 0.533 | 0.941 | -0.230 | 0.447 | 42 | 166 |
| Frontal pole | 0.146 | 0.171 | 0.399 | 0.941 | -0.190 | 0.482 | 43 | 165 |
| Temporal pole | 0.088 | 0.171 | 0.611 | 0.941 | -0.247 | 0.423 | 43 | 167 |
| Transverse temporal gyrus | 0.071 | 0.171 | 0.679 | 0.941 | -0.264 | 0.407 | 43 | 166 |
| Insula | 0.023 | 0.171 | 0.895 | 0.941 | -0.313 | 0.358 | 43 | 166 |
| Mean Thickness | -0.001 | 0.171 | 0.994 | 0.994 | -0.337 | 0.334 | 43 | 166 |
| **Cortical surface area** | | | | | | | | |
| Banks superior temporal sulcus | -0.142 | 0.180 | 0.437 | 0.941 | -0.494 | 0.211 | 39 | 150 |
| Caudal anterior cingulate cortex | -0.044 | 0.173 | 0.803 | 0.941 | -0.382 | 0.295 | 42 | 165 |
| Caudal middle frontal gyrus | 0.071 | 0.173 | 0.685 | 0.941 | -0.268 | 0.410 | 42 | 165 |
| Cuneus | 0.117 | 0.175 | 0.509 | 0.941 | -0.226 | 0.459 | 41 | 164 |
| Entorhinal cortex | 0.200 | 0.176 | 0.260 | 0.941 | -0.144 | 0.544 | 41 | 157 |
| Fusiform gyrus | 0.100 | 0.171 | 0.563 | 0.941 | -0.235 | 0.436 | 43 | 166 |
| Inferior parietal cortex | -0.110 | 0.173 | 0.531 | 0.941 | -0.449 | 0.229 | 42 | 165 |
| Inferior temporal gyrus | 0.040 | 0.173 | 0.818 | 0.941 | -0.298 | 0.379 | 42 | 165 |
| Isthmus cingulate cortex | 0.090 | 0.172 | 0.604 | 0.941 | -0.246 | 0.427 | 43 | 161 |
| Lateral occipital cortex | 0.187 | 0.173 | 0.287 | 0.941 | -0.153 | 0.526 | 42 | 165 |
| Lateral orbitofrontal cortex | 0.022 | 0.174 | 0.901 | 0.941 | -0.320 | 0.364 | 41 | 166 |
| Lingual gyrus | 0.134 | 0.173 | 0.444 | 0.941 | -0.205 | 0.473 | 42 | 166 |
| Medial orbitofrontal gyrus | -0.116 | 0.175 | 0.512 | 0.941 | -0.459 | 0.227 | 41 | 162 |
| Middle temporal gyrus | 0.024 | 0.177 | 0.895 | 0.941 | -0.323 | 0.371 | 40 | 157 |
| Parahippocampal gyrus | 0.320 | 0.173 | 0.067 | 0.941 | -0.019 | 0.658 | 43 | 160 |
| Paracentral lobule | -0.178 | 0.172 | 0.306 | 0.941 | -0.514 | 0.159 | 43 | 163 |
| Pars opercularis | 0.149 | 0.172 | 0.391 | 0.941 | -0.187 | 0.486 | 43 | 162 |
| Pars orbitalis | 0.341 | 0.174 | 0.052 | 0.941 | 0.001 | 0.682 | 42 | 165 |
| Pars triangularis | 0.062 | 0.173 | 0.722 | 0.941 | -0.276 | 0.401 | 42 | 165 |
| Pericalcarine cortex | 0.071 | 0.174 | 0.686 | 0.941 | -0.271 | 0.413 | 41 | 166 |
| Postcentral gyrus | -0.262 | 0.172 | 0.132 | 0.941 | -0.599 | 0.075 | 43 | 164 |
| Posterior cingulate cortex | -0.252 | 0.172 | 0.146 | 0.941 | -0.589 | 0.084 | 43 | 166 |
| Precentral gyrus | 0.101 | 0.171 | 0.561 | 0.941 | -0.235 | 0.436 | 43 | 165 |
| Precuneus | -0.053 | 0.171 | 0.762 | 0.941 | -0.389 | 0.283 | 43 | 164 |
| Rostral anterior cingulate cortex | -0.138 | 0.171 | 0.425 | 0.941 | -0.474 | 0.197 | 43 | 166 |
| Rostral middle frontal gyrus | 0.038 | 0.171 | 0.829 | 0.941 | -0.298 | 0.373 | 43 | 165 |
| Superior frontal gyrus | -0.077 | 0.171 | 0.658 | 0.941 | -0.412 | 0.259 | 43 | 166 |
| Superior parietal gyrus | 0.028 | 0.173 | 0.872 | 0.941 | -0.310 | 0.367 | 42 | 167 |
| Superior temporal gyrus | -0.007 | 0.179 | 0.971 | 0.984 | -0.358 | 0.344 | 39 | 156 |
| Supramarginal gyrus | -0.257 | 0.173 | 0.144 | 0.941 | -0.597 | 0.083 | 42 | 164 |
| Frontal pole | 0.037 | 0.171 | 0.833 | 0.941 | -0.299 | 0.372 | 43 | 166 |
| Temporal pole | 0.354 | 0.172 | 0.042 | 0.941 | 0.017 | 0.691 | 43 | 167 |
| Transverse temporal gyrus | 0.180 | 0.171 | 0.300 | 0.941 | -0.156 | 0.516 | 43 | 166 |
| Insula | -0.110 | 0.173 | 0.532 | 0.941 | -0.448 | 0.229 | 42 | 165 |
| Full surface area | -0.027 | 0.171 | 0.876 | 0.941 | -0.362 | 0.308 | 43 | 166 |

Table 19. Differences in regional brain morphology between healthy controls and young people with a lifetime history of suicide attempt **in women only**. D: Cohen’s d effect size, SE: standard error; p: p-value, FDR-p: FDR corrected p-value, CI: confidence interval, HC: healthy controls, CC: clinical controls.

| **Region** | **D** | **SE** | **P** | **FDR-p** | **Lower CI** | **Upper CI** | **N attempt** | **N HC** |
| --- | --- | --- | --- | --- | --- | --- | --- | --- |
| **Subcortical volume** | | | | | | | | |
| Ventricle | 0.051 | 0.140 | 0.716 | 0.963 | -0.223 | 0.326 | 105 | 99 |
| Thalamus | -0.330 | 0.112 | 0.003 | 0.166 | -0.549 | -0.110 | 166 | 158 |
| Caudate | -0.067 | 0.110 | 0.541 | 0.934 | -0.282 | 0.147 | 169 | 164 |
| Putamen | -0.027 | 0.114 | 0.814 | 0.967 | -0.250 | 0.196 | 159 | 151 |
| Pallidum | -0.078 | 0.112 | 0.490 | 0.910 | -0.298 | 0.142 | 166 | 153 |
| Hippocampus | -0.302 | 0.110 | 0.006 | 0.166 | -0.518 | -0.086 | 171 | 163 |
| Amygdala | -0.190 | 0.111 | 0.089 | 0.348 | -0.408 | 0.028 | 166 | 159 |
| Accumbens | -0.049 | 0.109 | 0.654 | 0.963 | -0.263 | 0.165 | 167 | 168 |
| **Cortical thickness** | | | | | | | | |
| Banks superior temporal sulcus | -0.034 | 0.110 | 0.761 | 0.966 | -0.250 | 0.183 | 159 | 169 |
| Caudal anterior cingulate cortex | 0.013 | 0.108 | 0.902 | 0.967 | -0.198 | 0.225 | 171 | 172 |
| Caudal middle frontal gyrus | -0.236 | 0.108 | 0.030 | 0.194 | -0.448 | -0.024 | 172 | 172 |
| Cuneus | -0.065 | 0.109 | 0.551 | 0.934 | -0.279 | 0.148 | 170 | 167 |
| Entorhinal cortex | 0.045 | 0.111 | 0.687 | 0.963 | -0.172 | 0.261 | 157 | 171 |
| Fusiform gyrus | -0.270 | 0.108 | 0.013 | 0.194 | -0.483 | -0.057 | 170 | 173 |
| Inferior parietal cortex | -0.076 | 0.108 | 0.483 | 0.910 | -0.288 | 0.136 | 170 | 173 |
| Inferior temporal gyrus | -0.050 | 0.108 | 0.646 | 0.963 | -0.262 | 0.162 | 171 | 172 |
| Isthmus cingulate cortex | 0.022 | 0.108 | 0.842 | 0.967 | -0.190 | 0.233 | 171 | 173 |
| Lateral occipital cortex | -0.245 | 0.108 | 0.024 | 0.194 | -0.458 | -0.033 | 171 | 173 |
| Lateral orbitofrontal cortex | -0.157 | 0.108 | 0.151 | 0.489 | -0.369 | 0.056 | 171 | 170 |
| Lingual gyrus | -0.266 | 0.108 | 0.015 | 0.194 | -0.478 | -0.053 | 172 | 171 |
| Medial orbitofrontal gyrus | -0.087 | 0.108 | 0.423 | 0.910 | -0.300 | 0.125 | 170 | 171 |
| Middle temporal gyrus | -0.143 | 0.110 | 0.197 | 0.579 | -0.359 | 0.073 | 160 | 171 |
| Parahippocampal gyrus | 0.078 | 0.108 | 0.471 | 0.910 | -0.134 | 0.291 | 169 | 173 |
| Paracentral lobule | -0.077 | 0.108 | 0.475 | 0.910 | -0.289 | 0.134 | 172 | 172 |
| Pars opercularis | -0.222 | 0.108 | 0.041 | 0.215 | -0.435 | -0.010 | 170 | 172 |
| Pars orbitalis | 0.019 | 0.108 | 0.860 | 0.967 | -0.192 | 0.230 | 172 | 172 |
| Pars triangularis | -0.223 | 0.108 | 0.040 | 0.215 | -0.435 | -0.011 | 172 | 172 |
| Pericalcarine cortex | -0.032 | 0.108 | 0.768 | 0.966 | -0.245 | 0.180 | 172 | 168 |
| Postcentral gyrus | -0.154 | 0.108 | 0.157 | 0.489 | -0.366 | 0.058 | 171 | 172 |
| Posterior cingulate cortex | -0.077 | 0.108 | 0.476 | 0.910 | -0.289 | 0.134 | 172 | 171 |
| Precentral gyrus | -0.211 | 0.108 | 0.052 | 0.239 | -0.423 | 0.001 | 171 | 173 |
| Precuneus | -0.259 | 0.108 | 0.017 | 0.194 | -0.471 | -0.047 | 172 | 173 |
| Rostral anterior cingulate cortex | -0.189 | 0.108 | 0.083 | 0.340 | -0.401 | 0.023 | 169 | 173 |
| Rostral middle frontal gyrus | -0.139 | 0.108 | 0.200 | 0.579 | -0.352 | 0.073 | 172 | 170 |
| Superior frontal gyrus | -0.100 | 0.108 | 0.358 | 0.872 | -0.312 | 0.112 | 171 | 171 |
| Superior parietal gyrus | -0.234 | 0.108 | 0.032 | 0.194 | -0.446 | -0.022 | 171 | 173 |
| Superior temporal gyrus | -0.242 | 0.110 | 0.029 | 0.194 | -0.457 | -0.026 | 163 | 171 |
| Supramarginal gyrus | -0.119 | 0.108 | 0.274 | 0.738 | -0.331 | 0.093 | 170 | 173 |
| Frontal pole | 0.022 | 0.108 | 0.842 | 0.967 | -0.190 | 0.234 | 170 | 172 |
| Temporal pole | -0.026 | 0.108 | 0.807 | 0.967 | -0.238 | 0.185 | 172 | 173 |
| Transverse temporal gyrus | -0.170 | 0.108 | 0.118 | 0.417 | -0.382 | 0.042 | 171 | 173 |
| Insula | -0.076 | 0.108 | 0.486 | 0.910 | -0.287 | 0.136 | 171 | 172 |
| Mean Thickness | -0.197 | 0.108 | 0.070 | 0.302 | -0.408 | 0.015 | 172 | 173 |
| **Cortical surface area** | | | | | | | | |
| Banks superior temporal sulcus | -0.246 | 0.112 | 0.029 | 0.194 | -0.466 | -0.027 | 158 | 163 |
| Caudal anterior cingulate cortex | -0.126 | 0.108 | 0.247 | 0.688 | -0.339 | 0.086 | 170 | 172 |
| Caudal middle frontal gyrus | -0.052 | 0.108 | 0.631 | 0.963 | -0.264 | 0.159 | 172 | 172 |
| Cuneus | -0.034 | 0.109 | 0.758 | 0.966 | -0.248 | 0.180 | 170 | 166 |
| Entorhinal cortex | -0.082 | 0.110 | 0.463 | 0.910 | -0.298 | 0.135 | 157 | 173 |
| Fusiform gyrus | -0.176 | 0.108 | 0.107 | 0.397 | -0.388 | 0.036 | 170 | 173 |
| Inferior parietal cortex | -0.233 | 0.108 | 0.032 | 0.194 | -0.445 | -0.021 | 172 | 172 |
| Inferior temporal gyrus | -0.300 | 0.109 | 0.006 | 0.166 | -0.513 | -0.086 | 171 | 171 |
| Isthmus cingulate cortex | 0.102 | 0.108 | 0.349 | 0.872 | -0.109 | 0.313 | 172 | 173 |
| Lateral occipital cortex | -0.067 | 0.108 | 0.536 | 0.934 | -0.278 | 0.144 | 172 | 173 |
| Lateral orbitofrontal cortex | -0.002 | 0.108 | 0.984 | 0.990 | -0.214 | 0.210 | 171 | 171 |
| Lingual gyrus | -0.001 | 0.108 | 0.990 | 0.990 | -0.213 | 0.210 | 172 | 172 |
| Medial orbitofrontal gyrus | 0.020 | 0.108 | 0.853 | 0.967 | -0.192 | 0.233 | 170 | 170 |
| Middle temporal gyrus | -0.041 | 0.110 | 0.710 | 0.963 | -0.257 | 0.175 | 161 | 169 |
| Parahippocampal gyrus | -0.036 | 0.108 | 0.744 | 0.966 | -0.247 | 0.176 | 170 | 173 |
| Paracentral lobule | 0.254 | 0.108 | 0.020 | 0.194 | 0.042 | 0.465 | 172 | 173 |
| Pars opercularis | 0.092 | 0.108 | 0.400 | 0.910 | -0.120 | 0.304 | 171 | 172 |
| Pars orbitalis | 0.043 | 0.108 | 0.693 | 0.963 | -0.168 | 0.254 | 172 | 173 |
| Pars triangularis | 0.157 | 0.108 | 0.148 | 0.489 | -0.054 | 0.369 | 172 | 173 |
| Pericalcarine cortex | 0.044 | 0.108 | 0.686 | 0.963 | -0.168 | 0.257 | 172 | 168 |
| Postcentral gyrus | 0.115 | 0.108 | 0.289 | 0.751 | -0.096 | 0.327 | 172 | 172 |
| Posterior cingulate cortex | 0.017 | 0.108 | 0.878 | 0.967 | -0.195 | 0.228 | 172 | 172 |
| Precentral gyrus | 0.001 | 0.108 | 0.989 | 0.990 | -0.210 | 0.213 | 170 | 173 |
| Precuneus | 0.043 | 0.108 | 0.690 | 0.963 | -0.168 | 0.255 | 172 | 172 |
| Rostral anterior cingulate cortex | -0.009 | 0.108 | 0.932 | 0.983 | -0.221 | 0.202 | 170 | 173 |
| Rostral middle frontal gyrus | -0.070 | 0.108 | 0.520 | 0.934 | -0.282 | 0.142 | 172 | 170 |
| Superior frontal gyrus | -0.002 | 0.108 | 0.982 | 0.990 | -0.214 | 0.209 | 171 | 171 |
| Superior parietal gyrus | 0.013 | 0.108 | 0.903 | 0.967 | -0.198 | 0.225 | 172 | 172 |
| Superior temporal gyrus | -0.082 | 0.110 | 0.459 | 0.910 | -0.299 | 0.134 | 163 | 165 |
| Supramarginal gyrus | -0.051 | 0.108 | 0.637 | 0.963 | -0.263 | 0.161 | 171 | 171 |
| Frontal pole | -0.218 | 0.108 | 0.045 | 0.219 | -0.430 | -0.007 | 172 | 173 |
| Temporal pole | 0.013 | 0.108 | 0.905 | 0.967 | -0.198 | 0.224 | 172 | 173 |
| Transverse temporal gyrus | -0.062 | 0.108 | 0.567 | 0.941 | -0.273 | 0.149 | 172 | 173 |
| Insula | 0.025 | 0.108 | 0.820 | 0.967 | -0.187 | 0.236 | 171 | 172 |
| Full surface area | -0.045 | 0.108 | 0.681 | 0.963 | -0.256 | 0.166 | 172 | 173 |

Table S20. Differences in regional brain morphology between clinical controls and young people with a lifetime history of suicide attempt **in women only**. D: Cohen’s d effect size, SE: standard error; p: p-value, FDR-p: FDR corrected p-value, CI: confidence interval, HC: healthy controls, CC: clinical controls.

| **Region** | **D** | **SE** | **P** | **FDR-p** | **Lower CI** | **Upper CI** | **N attempt** | **N CC** |
| --- | --- | --- | --- | --- | --- | --- | --- | --- |
| **Subcortical volume** | | | | | | | | |
| Ventricle | 0.085 | 0.119 | 0.477 | 0.793 | -0.148 | 0.319 | 105 | 215 |
| Thalamus | -0.185 | 0.093 | 0.048 | 0.520 | -0.368 | -0.002 | 166 | 379 |
| Caudate | -0.096 | 0.093 | 0.301 | 0.793 | -0.278 | 0.085 | 169 | 377 |
| Putamen | 0.009 | 0.095 | 0.929 | 0.953 | -0.177 | 0.195 | 159 | 368 |
| Pallidum | -0.135 | 0.094 | 0.149 | 0.793 | -0.319 | 0.048 | 166 | 369 |
| Hippocampus | -0.138 | 0.092 | 0.135 | 0.793 | -0.319 | 0.043 | 171 | 378 |
| Amygdala | -0.086 | 0.093 | 0.354 | 0.793 | -0.268 | 0.096 | 166 | 386 |
| Accumbens | -0.027 | 0.093 | 0.774 | 0.907 | -0.209 | 0.155 | 167 | 383 |
| **Cortical thickness** | | | | | | | | |
| Banks superior temporal sulcus | -0.111 | 0.095 | 0.246 | 0.793 | -0.298 | 0.076 | 159 | 359 |
| Caudal anterior cingulate cortex | 0.078 | 0.092 | 0.394 | 0.793 | -0.101 | 0.258 | 171 | 389 |
| Caudal middle frontal gyrus | 0.079 | 0.092 | 0.391 | 0.793 | -0.101 | 0.258 | 172 | 389 |
| Cuneus | -0.064 | 0.092 | 0.488 | 0.793 | -0.244 | 0.116 | 170 | 388 |
| Entorhinal cortex | 0.012 | 0.095 | 0.901 | 0.953 | -0.175 | 0.199 | 157 | 366 |
| Fusiform gyrus | -0.159 | 0.092 | 0.085 | 0.735 | -0.339 | 0.021 | 170 | 391 |
| Inferior parietal cortex | 0.125 | 0.092 | 0.175 | 0.793 | -0.055 | 0.306 | 170 | 388 |
| Inferior temporal gyrus | -0.064 | 0.092 | 0.488 | 0.793 | -0.244 | 0.116 | 171 | 387 |
| Isthmus cingulate cortex | 0.079 | 0.092 | 0.388 | 0.793 | -0.100 | 0.259 | 171 | 389 |
| Lateral occipital cortex | -0.024 | 0.092 | 0.793 | 0.907 | -0.204 | 0.156 | 171 | 386 |
| Lateral orbitofrontal cortex | -0.123 | 0.092 | 0.182 | 0.793 | -0.304 | 0.057 | 171 | 383 |
| Lingual gyrus | -0.136 | 0.092 | 0.138 | 0.793 | -0.316 | 0.043 | 172 | 387 |
| Medial orbitofrontal gyrus | -0.018 | 0.092 | 0.842 | 0.925 | -0.199 | 0.162 | 170 | 387 |
| Middle temporal gyrus | -0.136 | 0.095 | 0.154 | 0.793 | -0.322 | 0.050 | 160 | 367 |
| Parahippocampal gyrus | -0.015 | 0.092 | 0.873 | 0.945 | -0.195 | 0.166 | 169 | 391 |
| Paracentral lobule | 0.086 | 0.092 | 0.349 | 0.793 | -0.094 | 0.266 | 172 | 389 |
| Pars opercularis | -0.072 | 0.092 | 0.436 | 0.793 | -0.252 | 0.108 | 170 | 390 |
| Pars orbitalis | 0.060 | 0.092 | 0.516 | 0.821 | -0.120 | 0.239 | 172 | 386 |
| Pars triangularis | -0.040 | 0.092 | 0.666 | 0.869 | -0.219 | 0.140 | 172 | 389 |
| Pericalcarine cortex | -0.069 | 0.092 | 0.451 | 0.793 | -0.249 | 0.110 | 172 | 388 |
| Postcentral gyrus | -0.023 | 0.092 | 0.803 | 0.907 | -0.203 | 0.157 | 171 | 391 |
| Posterior cingulate cortex | 0.041 | 0.092 | 0.656 | 0.869 | -0.139 | 0.221 | 172 | 388 |
| Precentral gyrus | -0.067 | 0.092 | 0.465 | 0.793 | -0.247 | 0.113 | 171 | 388 |
| Precuneus | -0.089 | 0.092 | 0.330 | 0.793 | -0.269 | 0.090 | 172 | 391 |
| Rostral anterior cingulate cortex | -0.054 | 0.092 | 0.558 | 0.837 | -0.235 | 0.126 | 169 | 391 |
| Rostral middle frontal gyrus | 0.051 | 0.092 | 0.582 | 0.841 | -0.129 | 0.230 | 172 | 387 |
| Superior frontal gyrus | 0.071 | 0.092 | 0.442 | 0.793 | -0.109 | 0.251 | 171 | 390 |
| Superior parietal gyrus | 0.009 | 0.092 | 0.920 | 0.953 | -0.171 | 0.189 | 171 | 390 |
| Superior temporal gyrus | -0.183 | 0.094 | 0.053 | 0.520 | -0.367 | 0.002 | 163 | 370 |
| Supramarginal gyrus | -0.073 | 0.092 | 0.429 | 0.793 | -0.253 | 0.107 | 170 | 389 |
| Frontal pole | -0.047 | 0.092 | 0.608 | 0.862 | -0.227 | 0.133 | 170 | 391 |
| Temporal pole | -0.143 | 0.092 | 0.120 | 0.793 | -0.323 | 0.037 | 172 | 388 |
| Transverse temporal gyrus | -0.186 | 0.092 | 0.044 | 0.520 | -0.366 | -0.006 | 171 | 390 |
| Insula | -0.094 | 0.092 | 0.307 | 0.793 | -0.274 | 0.086 | 171 | 390 |
| Mean Thickness | -0.028 | 0.092 | 0.762 | 0.907 | -0.207 | 0.152 | 172 | 390 |
| **Cortical surface area** | | | | | | | | |
| Banks superior temporal sulcus | -0.138 | 0.096 | 0.150 | 0.793 | -0.326 | 0.049 | 158 | 357 |
| Caudal anterior cingulate cortex | -0.281 | 0.092 | 0.002 | 0.188 | -0.462 | -0.100 | 170 | 390 |
| Caudal middle frontal gyrus | -0.055 | 0.092 | 0.551 | 0.837 | -0.234 | 0.125 | 172 | 391 |
| Cuneus | 0.029 | 0.092 | 0.752 | 0.907 | -0.151 | 0.210 | 170 | 386 |
| Entorhinal cortex | -0.100 | 0.095 | 0.296 | 0.793 | -0.287 | 0.087 | 157 | 366 |
| Fusiform gyrus | -0.209 | 0.092 | 0.024 | 0.467 | -0.389 | -0.028 | 170 | 391 |
| Inferior parietal cortex | -0.182 | 0.092 | 0.049 | 0.520 | -0.362 | -0.002 | 172 | 387 |
| Inferior temporal gyrus | -0.234 | 0.092 | 0.011 | 0.298 | -0.414 | -0.053 | 171 | 388 |
| Isthmus cingulate cortex | 0.099 | 0.092 | 0.282 | 0.793 | -0.081 | 0.278 | 172 | 390 |
| Lateral occipital cortex | 0.039 | 0.092 | 0.668 | 0.869 | -0.140 | 0.219 | 172 | 388 |
| Lateral orbitofrontal cortex | -0.081 | 0.092 | 0.383 | 0.793 | -0.261 | 0.100 | 171 | 384 |
| Lingual gyrus | 0.028 | 0.092 | 0.759 | 0.907 | -0.151 | 0.208 | 172 | 390 |
| Medial orbitofrontal gyrus | -0.067 | 0.092 | 0.467 | 0.793 | -0.248 | 0.113 | 170 | 387 |
| Middle temporal gyrus | -0.029 | 0.095 | 0.759 | 0.907 | -0.215 | 0.156 | 161 | 365 |
| Parahippocampal gyrus | 0.045 | 0.092 | 0.627 | 0.869 | -0.135 | 0.225 | 170 | 390 |
| Paracentral lobule | 0.002 | 0.091 | 0.984 | 0.984 | -0.177 | 0.181 | 172 | 391 |
| Pars opercularis | 0.010 | 0.092 | 0.917 | 0.953 | -0.170 | 0.189 | 171 | 389 |
| Pars orbitalis | -0.056 | 0.092 | 0.545 | 0.837 | -0.235 | 0.124 | 172 | 390 |
| Pars triangularis | 0.093 | 0.092 | 0.311 | 0.793 | -0.086 | 0.273 | 172 | 390 |
| Pericalcarine cortex | 0.078 | 0.092 | 0.397 | 0.793 | -0.102 | 0.257 | 172 | 389 |
| Postcentral gyrus | 0.088 | 0.092 | 0.341 | 0.793 | -0.092 | 0.267 | 172 | 390 |
| Posterior cingulate cortex | -0.068 | 0.092 | 0.461 | 0.793 | -0.247 | 0.112 | 172 | 389 |
| Precentral gyrus | -0.051 | 0.092 | 0.581 | 0.841 | -0.231 | 0.129 | 170 | 391 |
| Precuneus | 0.018 | 0.091 | 0.841 | 0.925 | -0.161 | 0.198 | 172 | 391 |
| Rostral anterior cingulate cortex | -0.102 | 0.092 | 0.271 | 0.793 | -0.282 | 0.079 | 170 | 391 |
| Rostral middle frontal gyrus | -0.109 | 0.092 | 0.235 | 0.793 | -0.289 | 0.070 | 172 | 389 |
| Superior frontal gyrus | -0.035 | 0.092 | 0.704 | 0.901 | -0.215 | 0.145 | 171 | 390 |
| Superior parietal gyrus | -0.077 | 0.092 | 0.403 | 0.793 | -0.256 | 0.102 | 172 | 391 |
| Superior temporal gyrus | -0.082 | 0.094 | 0.384 | 0.793 | -0.267 | 0.102 | 163 | 369 |
| Supramarginal gyrus | -0.024 | 0.092 | 0.795 | 0.907 | -0.204 | 0.156 | 171 | 386 |
| Frontal pole | -0.252 | 0.092 | 0.006 | 0.250 | -0.432 | -0.072 | 172 | 390 |
| Temporal pole | -0.102 | 0.092 | 0.266 | 0.793 | -0.282 | 0.077 | 172 | 390 |
| Transverse temporal gyrus | -0.040 | 0.092 | 0.663 | 0.869 | -0.219 | 0.139 | 172 | 391 |
| Insula | -0.005 | 0.092 | 0.960 | 0.972 | -0.184 | 0.175 | 171 | 389 |
| Full surface area | -0.090 | 0.092 | 0.330 | 0.793 | -0.269 | 0.090 | 172 | 391 |

**Supplemental note 1.**

In this study, we pooled data across 21 international research studies, which had used different instruments to assess suicidal thoughts and suicide attempt. While our previous work shows moderate to high correlations between different instruments of suicidal thoughts and suicide attempt (1) we have used a detailed approach to harmonise these measures as much as possible.

The following approach was used to harmonise the different instruments used to assess suicidal ideation (thoughts) and attempts. In analyses on suicide attempt, we examined lifetime suicidal attempt (coded yes/no), and in analyses on suicidal ideation (thoughts), we examined current suicidal ideation (in the past week, two weeks or month).

If sites had only one measure available on lifetime suicide attempt or suicidal ideation in the past week, two weeks or month, this information was used to code suicide attempt (yes/no) or current suicidal ideation (yes/no).

When sites had collected multiple measures on lifetime suicide attempt, we used the most detailed measure. For instance, when available, the lifetime non-interrupted suicide attempt (yes/no) measure from the Columbia Suicide Severity Rating Scale (C-SSRS) was used when available. When the C-SSRS was not available, information on lifetime information (coded yes/no) from clinical interviews were used. In addition, when multiple measures were available, we were mindful to select the measure that was most often used by other sites, and showed good correlations with other measures (1).

When sites had collected multiple measures on current suicidal ideation (in the past week, two weeks or month), we again used the most detailed measure. In this case, current suicidal ideation was determined by the C-SSRS or Beck Scale for Suicidal Ideation when this information was available. When there were multiple measures on suicidal ideation from depression severity rating scales (such as the Beck Depression Inventory, Montgomery-Asberg Depression Rating Scale or the Hamilton Depression Rating Scale), current suicidal ideation was defined as endorsement of current suicidal ideation on any of these rating scales. Again here, when multiple measures were available, we were mindful to select the measures that were most often used by other sites, and showed good correlations with other measures (1).
